# A Systematic Literature Review and Meta-Analysis of the Effects of Lockdowns on COVID-19 Mortality II

**DOI:** 10.1101/2023.08.30.23294845

**Authors:** Jonas Herby, Lars Jonung, Steve H. Hanke

## Abstract

The purpose of this systematic review and meta-analysis is to determine the effect of lockdowns on COVID-19 mortality based on available empirical evidence. Lockdowns are defined as the imposition of at least one compulsory, non-pharmaceutical intervention (NPI). We employ a systematic search and screening procedure in which 19,646 studies are identified that could potentially address the purpose of our study. After three levels of screening, 32 studies qualified. Of those, estimates from 22 studies could be converted to standardized measures for inclusion in the meta-analysis. They are separated into three groups: lockdown stringency index studies, shelter-in-place-order (SIPO) studies, and specific NPI studies. Stringency index studies find that the average lockdown in Europe and the United States in the spring of 2020 only reduced COVID-19 mortality by 3.2%. This translates into approximately 6,000 avoided deaths in Europe and 4,000 in the United States. SIPOs were also relatively ineffective in the spring of 2020, only reducing COVID-19 mortality by 2.0%. This translates into approximately 4,000 avoided deaths in Europe and 3,000 in the United States. Based on specific NPIs, we estimate that the average lockdown in Europe and the United States in the spring of 2020 reduced COVID-19 mortality by 10.7%. This translates into approximately 23,000 avoided deaths in Europe and 16,000 in the United States. In comparison, there are approximately 72,000 flu deaths in Europe and 38,000 flu deaths in the United States each year. When checked for potential biases, our results are robust. Our results are also supported by the natural experiments we have been able to identify. The results of our meta-analysis support the conclusion that lockdowns in the spring of 2020 had little to no effect on COVID-19 mortality. This result is consistent with the view that voluntary changes in behavior, such as social distancing, did play an important role in mitigating the pandemic.

## 1 Introduction

Social distancing works. If you keep distance from others, your risk of being infected with a communicable disease is reduced. However, the fact that social distancing works does not imply that compulsory non-pharmaceutical interventions (NPIs), commonly known as “lockdowns” – policies that restrict internal movement, close schools and businesses, ban international travel and/or other activities – work. If governments primarily close activities which hardly anyone wants to participate in during an ongoing pandemic, the effect of the lockdown will be modest. If there is too much non-compliance, the effect of the lockdown will be modest. If government only regulates a fraction of the activities where people can become infected, the effect of the lockdown will be modest. If people react strongly to lower infection rates following lockdowns by being much less careful, the effect of the lockdown will be modest. If, if, if…

Although many people perceive lockdowns as extremely effective in reducing Coronavirus Disease 2019 (COVID-19) infections and mortality, it is today – from a research perspective – unknown to what extent lockdowns did in fact reduce COVID-19 infections and COVID-19 mortality. The goal of this study is to answer the following research question: “Were lockdowns effective in reducing COVID-19 mortality?” We also examine if some NPIs were more effective than others.

### Definition of “lockdown” and “NPI”

We use “NPI” to describe *any government mandate which directly restrict peoples’ possibilities*. Our definition does *not* include governmental recommendations, governmental information campaigns, access to mass testing, voluntary social distancing, etc., but *do* include mandated interventions such as closing schools or businesses, mandated face masks, etc. During the COVID-19 pandemic, lockdowns have mainly been used to describe two different things. Some use “lockdown” under the definition of “a period of time in which people are not allowed to leave their homes or travel freely”. Others use “lockdown” more broadly to describe governments’ responses to the pandemic in terms of less or more strict interventions.^1^ We follow the latter use and define *lockdown* as any policy consisting of at least one NPI as described above.^2^ We use shelter-in-place order (SIPO) to describe the former use of the term “lockdown”.

Our focus is on the effect of compulsory NPIs, policies that, for example, restrict internal movement, close schools, and businesses, ban international travel, etc. We do not look at the effect of voluntary behavioral changes (e.g., voluntary mask wearing), the effect of recommendations (e.g., recommended mask wearing), or governmental services (e.g. voluntary mass testing).

The first NPIs were implemented in China. From there, the pandemic and NPIs spread first to Italy and later to virtually all other countries (see Figure 1).Of the 186 countries covered by the Oxford COVID-19 Government Response Tracker (OxCGRT),^3^ only Comoros, an island country in the Indian Ocean with a population below 1 million, did not impose at least one NPI (as defined by OxCGRT) before the end of March 2020. Since virtually all countries have implemented some sort of restrictions, we are essentially studying how the degree of lockdowns affects mortality rates.

**Figure 1:**
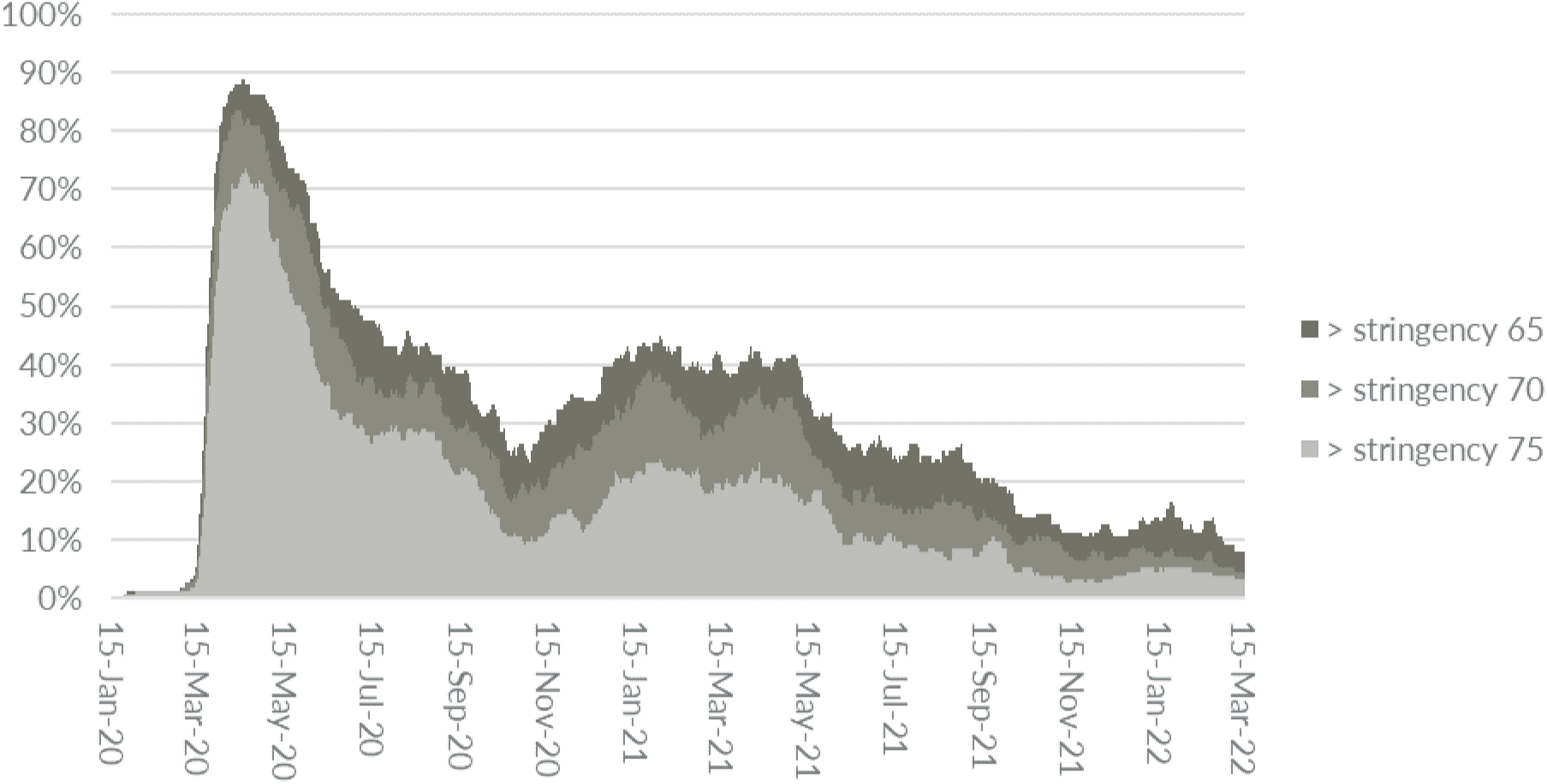
Percentage of countries with Oxford COVID-19 Government Response Tracker (OxCGRT) stringency index readings above thresholds 65, 70, and 75, respectively. Source: Our World in Data (2022). Comment: The OxCGRT Stringency Index measures the stringency of lockdowns on a scale from 0 to 100, where a higher value corresponds to stricter lockdowns. The figure shows the share of countries, where the OxCGRT stringency index on a given date surpassed index 65, 70 and 75 respectively. Only countries with more than one million citizens are included (153 countries in total). The OxCGRT stringency index records the strictness of NPI policies that restrict people’s behavior. It is calculated using all ordinal containment and closure policy indicators (i.e., the degree of school and business closures, etc.), plus an indicator recording public information campaigns.

### Do lockdowns work?

One could question the necessity to examine the effectiveness of certain NPIs that have been used for centuries. However, although NPIs such as school and workplace closures were recommended by the World Health Organization (WHO) before the COVID-19 pandemic in the event of an extraordinarily severe pandemic influenza, the evidence of the effectiveness of such measures were, in general, very low (see Table 1).

**Table 1:**
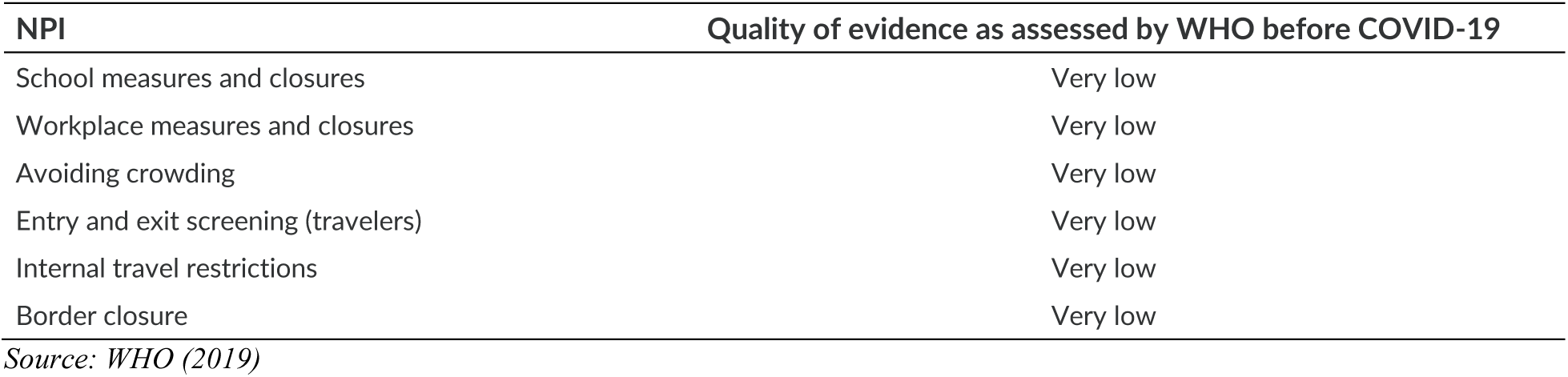
Quality of evidence for selected NPIs as assessed by WHO before the COVID-19 pandemic.

Despite the very low quality of evidence (see Table 1), early epidemiological studies predicted that NPIs would have large effects. An often cited model simulation study by researchers at Imperial College London, Ferguson et al. (2020), predicted that a suppression strategy would reduce COVID-19 mortality by up to 99%.^4^ Ferguson et al. (2020) state that “it is highly likely that there would be significant spontaneous changes in population behavior even in the absence of government-mandated interventions” but also that “any one intervention in isolation is likely to be limited, requiring multiple interventions to be combined to have a substantial impact on transmission” and “we predict that transmission will quickly rebound if interventions are relaxed” causing many to perceive their projections as a forecast in the case of no lockdown.^5^ And many hold the results from Ferguson et al. (2020) responsible for the subsequent lockdown in United Kingdom.^6,7^

Already early in the pandemic, there was reason to question whether lockdowns were as effective as promised.^8^ First, there was no clear negative correlation between the degree of lockdown and actual outcomes on fatalities in the spring of 2020 (see Figure 2). Given the large perceived effect of lockdowns that studies such as Ferguson et al. (2020) had led to, one would expect to at least observe a simple negative correlation between COVID-19 mortality and the degree to which lockdowns were imposed.

**Figure 2:**
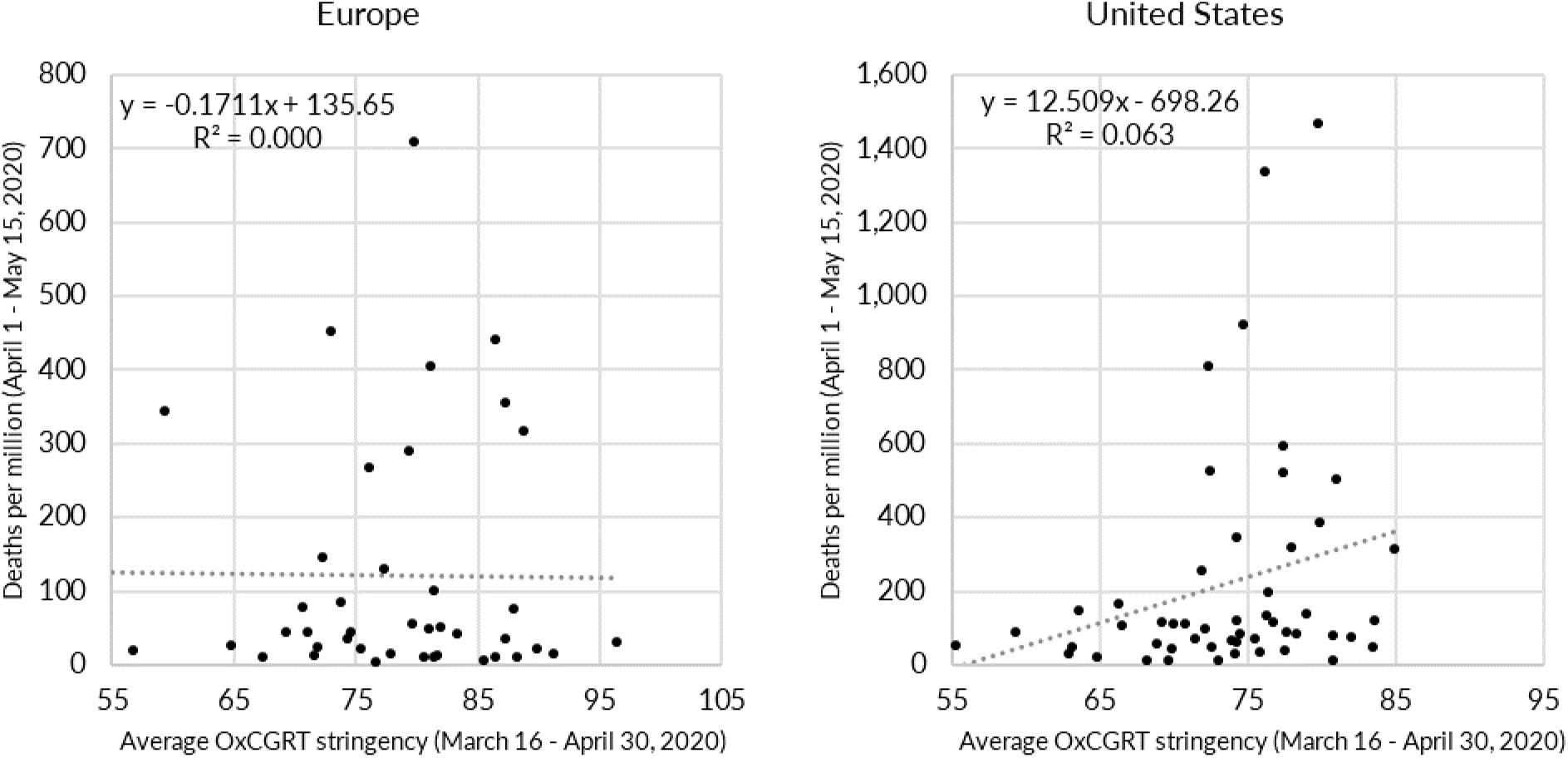
The positive correlation between the OxCGRT stringency index and COVID-19 mortality in 44 European countries and the 50 U.S. states (and Washington, D.C.) during the first wave in 2020. Source: Our World in Data (2022). Note: The OxCGRT stringency Index measures the stringency of lockdowns on a scale from 0 to 100, where a higher value corresponds to stricter lockdowns. Isle of Man and North Macedonia are not included, because there is no stringency index data for these two countries before April 30, 2020.

Second, several studies pointed to facts questioning the effect of lockdowns. For example, Atkeson et al. (2020) showed in August 2020 that “across all countries and US states that we study, the growth rates of daily deaths from COVID-19 fell from a wide range of initially high levels to levels close to zero within 20-30 days after each region experienced 25 cumulative deaths”. Goolsbee and Syverson (2021) found in June 2020 that “legal shutdown orders account for only a modest share of the decline of economic activity […]. While overall consumer traffic fell by 60 percentage points, legal restrictions explain only 7 of that. Individual choices were far more important and seem tied to fears of infection. Traffic started dropping before the legal orders were in place; was highly tied to the number of COVID deaths in the county; and showed a clear shift by consumers away from larger/busier stores toward smaller/less busy ones in the same industry.”

Although the externalities associated with a communicable disease like COVID-19 are evident, it is less clear how these externalities can be regulated effectively. ^9^ Specifically, it remains an open question as to whether lockdowns have had a large, significant effect on COVID-19 mortality.

### Our contribution

We address the question “Were lockdowns effective in reducing COVID-19 mortality?” by evaluating the current academic literature on the relationship between lockdowns and COVID-19 mortality rates.^10^ Our analysis is based on the evidence found in studies published between January 1, 2020, and February 21, 2022.

We are still in the early phase of the scientific and quantitative evaluation of the effects of lockdowns, and future research will continue to improve our understanding of lockdowns. Still, we find it valuable to summarize in a consistent way the evidence available from the first two years of the pandemic.

Compared to other reviews such as Herby (2021a) and Allen (2021), the main difference in our approach is that we carry out a systematic and comprehensive search strategy to identify all papers potentially relevant, and carry out a meta-analysis combining evidence from several existing studies to answer the question we pose. Results need repeated replication to be believable, but replication is unfortunately rare. Mueller-Langer et al. (2019) find that only 0.1% of publications in the top 50 economics journals were replication studies. However, as described by Paldam (2022), the same question is often analyzed in many studies, which use different data samples, estimation models, control variables, etc. The theories of the studies may differ, but the estimation models are often similar. Thus, instead of strict replication, there are often several partial replications. A meta-analysis is a technique developed to analyze if the aggregation of the evidence from studies analyzing the same question leads to a general result. Thus, in our meta-analysis, we aim at replacing replication by presenting results in such a way that they can be systematically assessed and used to derive overall conclusions.

In Figure 3 below, we compare the measured results from our meta-analysis to the forecasts derived from the models used in Ferguson et al. (2020). Overall, the meta-analysis does not support the notion that lockdowns in the spring of 2020 had a large effect on COVID-19 mortality, as many modelling studies had concluded they would.

**Figure 3:**
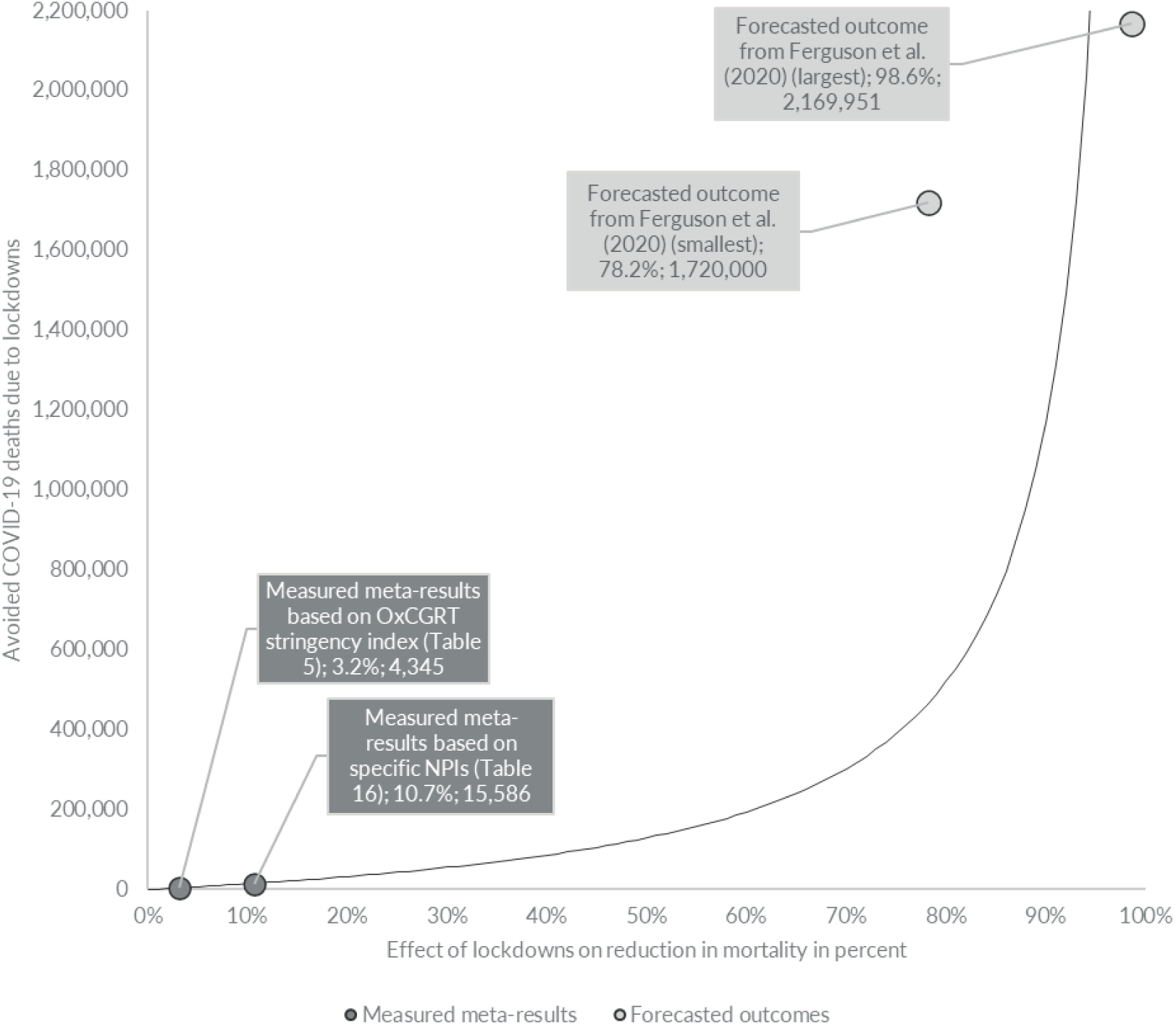
Divergence between avoided number of deaths in the United States as measured by our meta-results and the forecasted outcome from Imperial College London. Note: The effect of lockdowns on total mortality based on the meta-study’s precision-weighted averages (PWA) is calculated as total COVID-19 deaths by July 1, 2020 (128,063 COVID-19 deaths) × (1/(1-PWA)-1). The relative effect of lockdowns on total mortality based on Ferguson et al. (2020) is calculated as the largest and smallest predicted relative effect multiplied with their mortality estimate of 2.2 million deaths in a “do nothing”-scenario in the United States. For more details see Figure 10 p. 59.

### Updates in this version

The first version of this literature review and meta-analysis, Herby et al. (2022), generated countless comments ranging from ad-hominem, wrong, and irrelevant – to of use and of constructive value. For more on the discussion of the first version of this literature review and meta-analysis, see Appendix II.

In this updated version, we have incorporated all constructive comments. Paragraphs have been rewritten for clarity. For example, it was not clear to some commentators that we distinguished between the effect of social distancing and the effect of lockdowns. We have tried to make this distinction more transparent, and the first sentence in the introduction is now “Social distancing works”. Our definition of lockdown also created confusion among some critics, so we have elaborated our definition to make it clearer. We have also added more examples to support the understanding of our results and conclusions. For example, we have elaborated section 5.2.3 where we discuss why the effect of lockdowns – as our measured meta-results show – was limited.

Our results have also changed for three reasons. First, we have excluded some studies which we now believe to be ineligible. Second, we have updated our literature search, so more studies are now included. Finally, we have changed some calculations. It is worth noting that we, not the commentators, did identify and correct one computational error that was contained in the first version of this study. Its correction had no significant effect on our overall conclusion.

We have also expanded the section on specific NPIs to give a more in-depth analysis of the results. These updates have changed our estimates, but not the overall conclusion. We believe that one major mistake in our first version was our failure to explain that the overall conclusions do not depend on whether the impact of lockdowns on COVID-19 mortality was 0.2%, 3.0% or 15%. As Figure 3 illustrates, all cases based on actual measurements of saved lives due to lockdowns are much smaller and far removed from the promises made by many epidemiologists, politicians, and the media.

Our literature review and meta-analysis is organized in the following way. In section 2, we describe our identification process for selecting relevant studies. That is, we explain our search strategy and eligibility criteria. In section 3, we present an account of the empirical evidence. Section 4 contains our meta-analysis of the impact of lockdowns on COVID-19 mortality. Section 5 contains our concluding observations and discussion.

## 2 Identification process: Search strategy and eligibility criteria

Figure 4 presents an overview of our identification process. It uses a flow diagram designed according to Preferred Reporting Items for Systematic Reviews and Meta-Analyses (PRISMA) guidelines by Moher et al. (2009). Of 19,646 studies identified during our database searches, 1,220 remained after a title-based screening. Then 1,074 studies were excluded because they either did not measure the effect of lockdowns on mortality or did not use an empirical approach. This left 146 studies that were read and inspected carefully. After a more thorough assessment, 114 of the 146 were excluded, leaving 32 eligible studies. The 114 studies excluded in the final step are listed in Table 18 in Appendix I.

**Figure 4:**
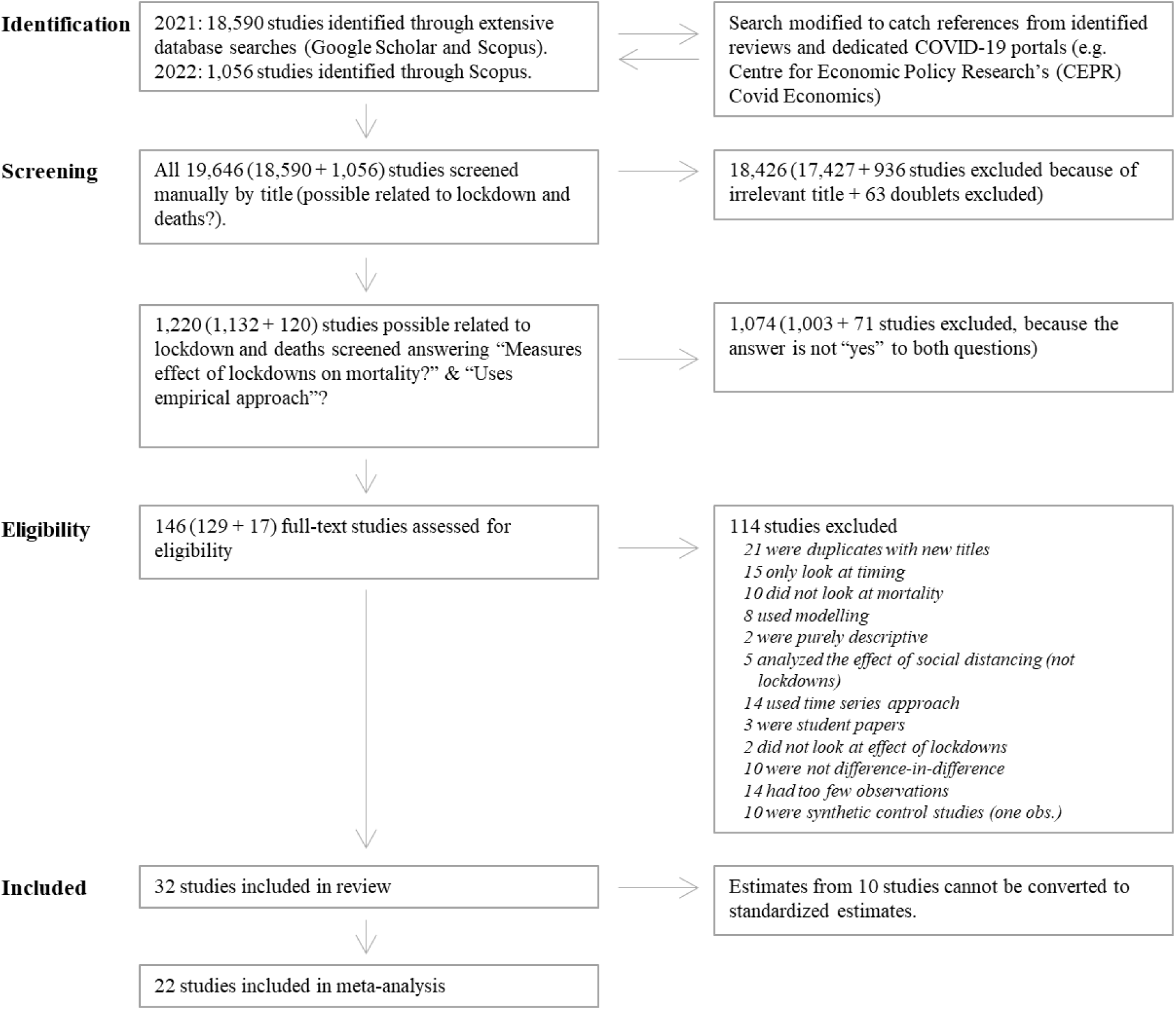
The PRISMA flow diagram for the selection of studies.

The inclusion rate of 32 eligible studies out of 19,646 identified studies (0.2%) is in the range of other systematic literature reviews on the topic of COVID-19 lockdowns. Our inclusion rate is similar to Talic et al. (2021), who include 72 of 36,729 identified studies (0.2%) and Iezadi et al. (2021) (35 of 12,523, 0.3%), while is relatively low compared to e.g. Rezapour et al. (2021) (26 of 2,176 studies, 1.2%), Zhang et al. (2021) (47 of 1,649 studies, 2.9%), and Johanna et al. (2020) (14 of 623 studies, 2.2%). A major reason for the difference in the inclusion rate is the choice of search strategy. Rezapour et al. (2021), Zhang et al. (2021) and Johanna et al. (2020) identify studies by searching publication databases such as PubMed, Scopus, Web of Science, SAGE, etc., while our search in Google Scholar is more broad. For example, our search also includes presentations and books. Performing our search only with Scopus, instead of both Google Scholar and Scopus, results in an inclusion rate of 0.7%.

Below, we present our search strategy and eligibility criteria, which follow the PRISMA guidelines and are specified in detail in our protocol that we posted in 2021, Herby et al. (2021).^11^

### 2.1 Search strategy

The studies we reviewed were identified by scanning Google Scholar and Scopus for English-language studies. We used a wide range of search terms which are combinations of three search strings: a disease search string (“covid”, “corona”, “coronavirus”, “sars-cov-2”), a government response search string^12^, and a methodology search string.^13^ We identified papers based on 1,360 search terms. We also required mentions of “deaths”, “death”, and/or “mortality”. The search terms were continuously updated (by adding relevant terms) to fit our criteria.^14^

We also included all papers published in *Covid Economics*. Our first search was performed between July 1 and July 5, 2021 and resulted in 18,590 unique studies. All studies identified using Scopus and *Covid Economics* were also found using Google Scholar. This made us comfortable that including other sources such as VOXeu and SSRN would not materially change the result. Indeed, many papers found using Google Scholar were from these sources. On February 21, 2022, we repeated our search on Scopus resulting in another 1,056 studies.

All 19,646 (18,590 from July 2021, and 1,056 from February 2022) studies were first screened based on the title. Studies clearly not related to our research question were deemed irrelevant.^15^

After screening based on the title, 1,088 papers remained. These papers were manually screened by answering two questions:

1. Does the study measure the effect of lockdowns on mortality?
2. Does the study use an empirical *ex post* difference-in-difference approach (see eligibility criteria below)?

Studies to which we could not answer “yes” to both questions were excluded. When in doubt, we made the assessment based on reading the full paper, and in some cases, we consulted colleagues.^16^

After the manual screening, 146 studies were retrieved for a full, detailed inspection. These studies were carefully examined, and metadata and empirical results were stored in an Excel spreadsheet. All studies were assessed by at least two researchers. During this process, another 114 papers were excluded because they did not meet our eligibility criteria. A table with all 114 studies excluded in the final step can be found in Appendix I, Table 18. Below we explain the most important of our eligibility criteria. A full list can be found in our protocol Herby et al. (2021).

### 2.2 Eligibility criteria

#### Focus on mortality and lockdowns

We only include studies that attempt to establish a relationship (or lack thereof) between lockdown policies and COVID-19 mortality or excess mortality. Following our protocol Herby et al. (2021), we exclude studies that use cases, hospitalizations, or other measures.

While we regard analysis based on cases problematic because of large data problems,^17^ one could argue that including studies examining the effect of lockdowns on hospitalization could improve the quality of our review and meta-analysis because it would allow us to include more studies. Using the same search strings at Scopus, but replacing (“deaths”, “death”, and/or “mortality”) with (“hospitalization”, “intensive care”, and/or “ICU”), indicates that including hospitalizations would yield another 1-2 eligible studies.^18^

Although including studies examining the effect of lockdowns on hospitalization would potentially strengthen our results by adding more studies to the review and meta-analysis, we see little reason to believe that doing so would change our results significantly. It is true that a key argument for locking down countries around the world was to protect the healthcare sector and keep hospitalizations down. But one of the arguments for protecting the healthcare sector was that if hospitalizations were high and hospitals were overcrowded, there would be an unusually high excess mortality rate because COVID-19 patients would not be able to receive treatment.^19^ Given this relationship between hospitalizations and deaths, we should see the effect of hospitalizations in our analysis of mortality.

#### Assessment of actual outcomes (in contrast to modelled outcomes)

There are two different approaches to examine the relationship between mortality rates and lockdown policies. The first approach uses actual, measured mortality data. These are *ex post* studies based on actual mortality outcomes. The second approach uses simulated data on mortality and infection rates generated from models.^20^ These are *ex ante* studies based on modelled outcomes.

In this review and meta-analysis, we include all studies from the former group but exclude all *ex ante* studies, as the results from these studies are determined by model assumptions and calibrations and cannot be the basis for solid empirical evidence for policy purposes, before these models have been validated empirically which is exactly the point of our study. This means that we exclude e.g. the much-cited Ferguson et al. (2020) from Imperial College.

#### Counterfactual difference-in-difference approach

We exclude studies which do not use a counterfactual difference-in-difference approach (called ‘controlled before-and-after studies’ in some social sciences).^21^

Difference-in-difference is a quasi-experimental design that makes use of longitudinal data from treatment and control groups to obtain an appropriate counterfactual to estimate a causal effect. Difference-in-difference is typically used to estimate the effect of a specific intervention (for example, a non-pharmaceutical intervention) by comparing the changes in outcomes over time between a treatment group (where a specific NPI was in place) and a control group (where the NPI was not in place).

Difference-in-difference is used in observational settings where exchangeability cannot be assumed between the treatment and control groups, as is the case for COVID-19 mortality, where mortality rates differ between countries and over time. The difference-in-difference method relies on a somewhat weaker – although not negligible – exchangeability assumption known as the parallel, or common, trends assumption as illustrated in Figure 5, Panel A.^22^

**Figure 5:**
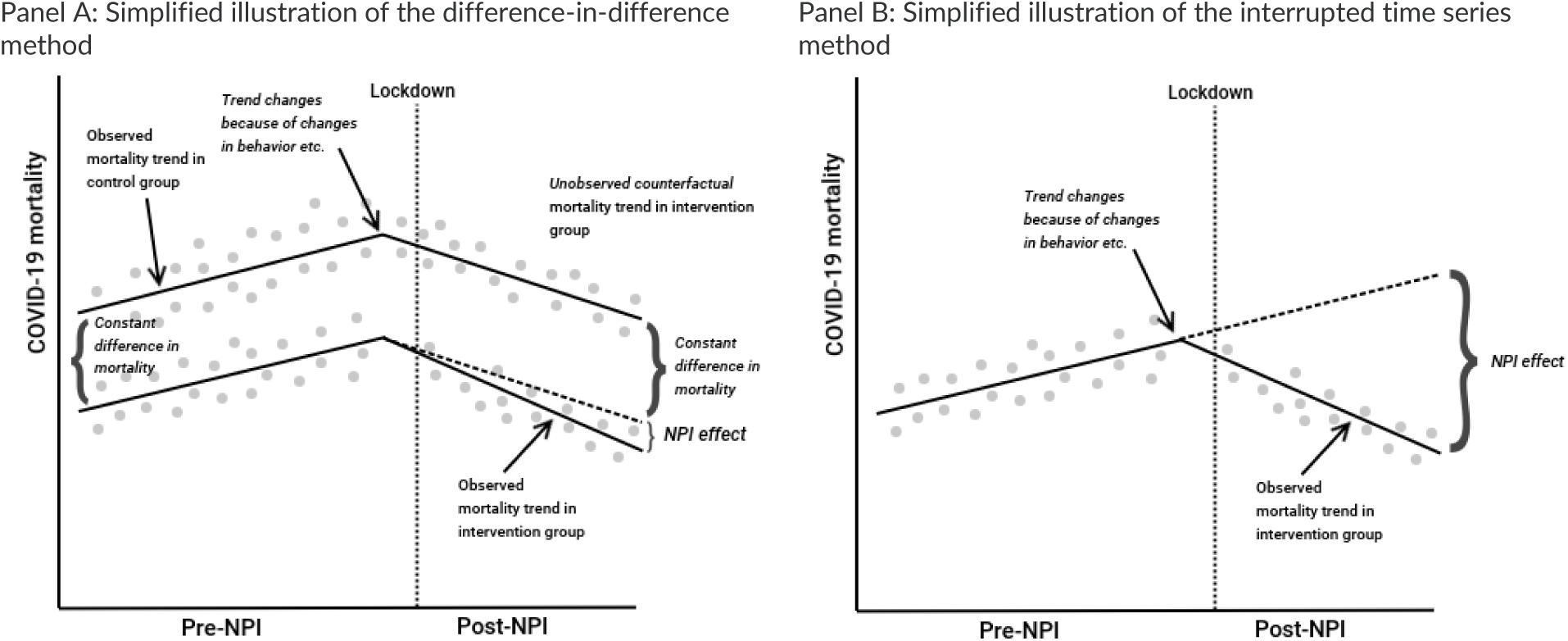
Simplified illustration of the difference-in-difference approach compared to interrupted time series approach when trends changes.

Difference-in-difference is a useful technique to employ when examining the effect of lockdowns where randomization is not possible. The approach removes biases in post-intervention period comparisons between the treatment and control group that could be the result from permanent differences between those groups (e.g. caused by coincidences early in the pandemic^23^), as well as biases from comparisons over time in the treatment group that could be the result of trends due to other causes of the outcome (e.g. changes in behavior or seasonality).

The exclusion of studies that do not use a counterfactual difference-in-difference approach means that we exclude all studies where the counterfactual is based on forecasting (for example, using a SIR-model calibrated on mortality data). This means that we excludes studies like Duchemin et al. (2020) and Matzinger and Skinner (2020). We also exclude all studies based on interrupted time series designs. Interrupted time series designs are useful when there is a stable long-term period before and after the time of the intervention examined (lockdowns), and where other things are relatively constant and/or can be controlled for. This is not the case with COVID-19 and lockdowns, where the period before (and often after) the intervention is short, where things are far from constant, and where changes in behavior cannot easily be controlled for. As illustrated in Figure 5, Panel B, interrupted time series risk overestimating the effect of lockdowns, if, for example, voluntary behavioral changes are important. Excluding interrupted time series studies rule out studies like Bakolis et al. (2021) and Siedner et al. (2020).

Given our criteria, we also exclude the much-cited paper by Flaxman et al. (2020), which claimed that lockdowns saved three million lives in Europe. Flaxman et al. (2020) assume that the pandemic would follow an epidemiological curve unless countries locked down. However, this assumption means that the only interpretation possible for the empirical results is that lockdowns are the only factor that matters, even if other factors like changes in voluntary behavior, seasonality, etc. caused the observed change in the reproduction rate, R_t_. Figure 6 illustrates how problematic Flaxman’s assumption is. The figure shows Flaxman et al. (2020)’s estimate of the effect of various NPIs on the effective reproduction number, R ^24^ in Denmark and Sweden. According to the results, banning public events had close-to-zero effect in Denmark (Panel A) but huge effects in Sweden (Panel B).

**Figure 6.**
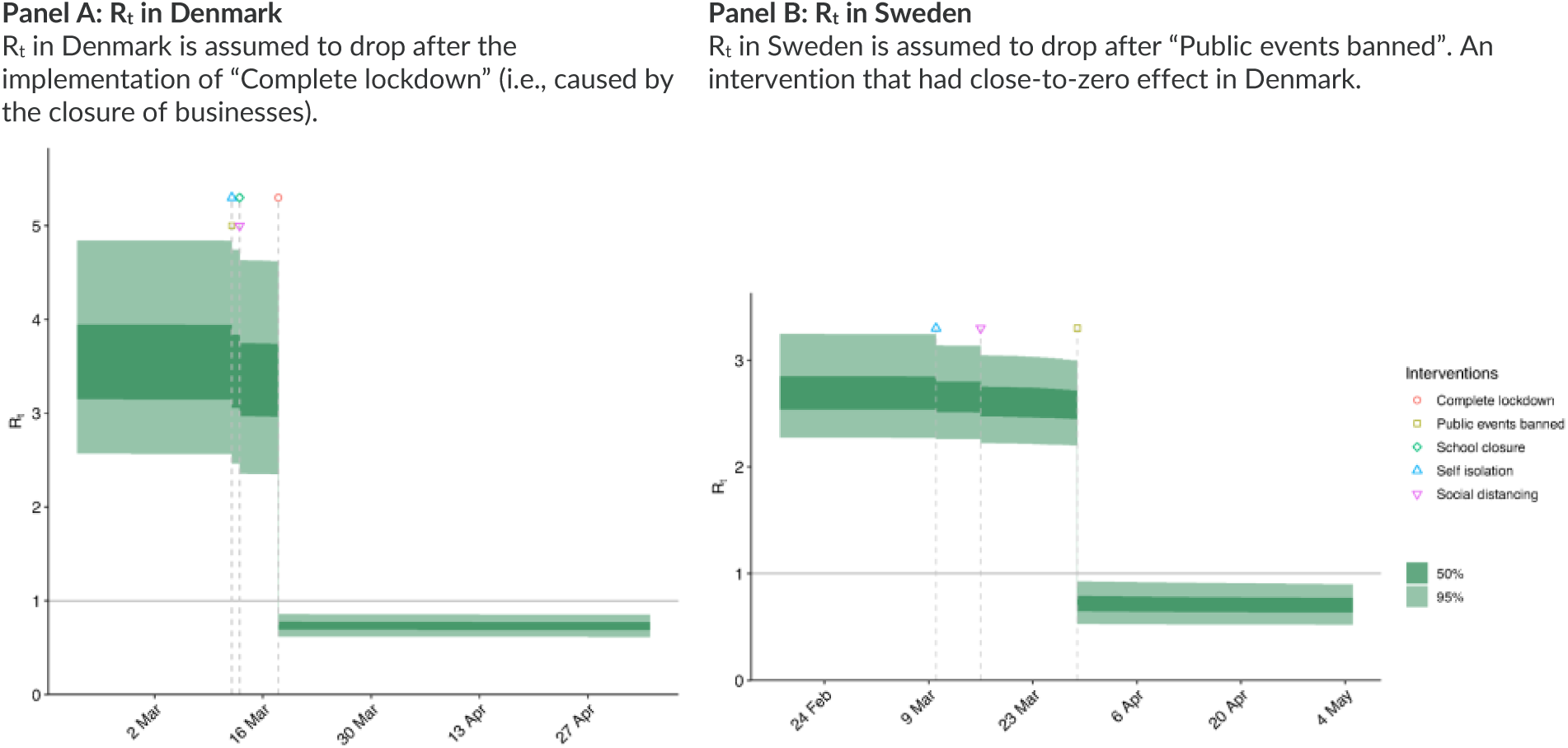
The assumptions used by Flaxman, et al. lead to two contradictory conclusions: That banning public events had no effect in Denmark but were extremely effective in Sweden in March 2020. Source: Flaxman et al. (2020), Extended Data Fig. 1 & Fig. 2.

Flaxman et al. (2020) are aware of this problem and state that “our parametric form of R_t_ assumes that changes in R_t_ are an immediate response to interventions rather than gradual changes in behavior”. Despite stating that the results cannot be interpreted as an effect of lockdowns, media around the globe – supported by statements by the study’s authors^25^ – reported these findings as “proof” that lockdowns had saved millions of lives.^26^ Similar interpretation problems are typical and are found in Brauner et al. (2021)^27^ and Hsiang et al. (2020) for example.

In our view, Flaxman et al. (2020), Brauner et al. (2021), Hsiang et al. (2020), etc. illustrate how problematic it is to force data to fit a certain model and certain assumptions if you want to infer the effect of lockdowns on COVID-19 mortality, and how these assumptions – while not being academically incorrect, as they are readily available in their paper – can lead to misguided perceptions in the media.^28^ Including the estimates from these studies and interpreting them as the effect of lockdowns would without any doubt greatly overstate the effectiveness of lockdowns.

#### Jurisdictional variance - few observations

We also exclude studies with little jurisdictional variance.^29^ For example, we exclude Conyon et al. (2020) who “exploit policy variation between Denmark and Norway on the one hand and Sweden on the other” and, thus, only have one jurisdictional area in the control group. Although this is a difference-in-difference approach, there is a non-negligible risk that differences are caused by much more than just differences in lockdowns (As of matter of fact, research has shown that Sweden was particularly unlucky in the spring of 2020. Arnarson (2021) and Björk et al. (2021) show that areas in Europe – like Sweden – where the winter holiday was relatively late (in week 9 or 10 rather than week 6, 7 or 8) were hit especially hard by COVID-19 during the first wave because the virus outbreak in the Alps could spread to those areas with ski tourists). Another example is Wu and Wu (2020), who use all U.S. states, but pool groups of states so they end up with basically three observations.^30^ None of the excluded studies cover more than 10 jurisdictional areas.

#### Synthetic control studies

The synthetic control method is a special case of the difference-in-difference method used to evaluate the effect of an intervention in comparative case studies. It involves the construction of a synthetic control which functions as the counterfactual and is constructed as an (optimal) weighted combination of a pool of donors. For example, Born et al. (2021) create a synthetic control for Sweden which consists of 30.0% Denmark, 25.3% Finland, 25.8% Netherlands, 15.0% Norway, and 3.9% Sweden. The effect of the intervention is derived by comparing the actual developments to those derived through the synthetic control. We exclude synthetic control studies because of too little jurisdictional variance, as these studies examine the effects of lockdowns based on one country/state compared to a synthetic counterfactual.

But – as discussed by Bjørnskov (2021b) – synthetic control studies also have empirical problems in relation to studying the effect of lockdowns. Bjørnskov (2021b) finds that the synthetic control version of Sweden in Born et al. (2021) deviates substantially from “actual Sweden”, when looking at the period before mid-March 2020, when Sweden decided not to lock down. Bjørnskov (2021b) estimates that *actual Sweden* experienced approximately 500 fewer deaths the first 11 weeks of 2020 and 4,500 fewer deaths in 2019 compared to *synthetic Sweden*. Such empirical problems are inherent to all synthetic control studies of COVID-19 because the synthetic control should be fitted based on a long period of time before the intervention or the event one is studying the consequences of – i.e., the lockdown.^31^ This is not possible for the coronavirus pandemic, as there clearly is no long period with coronavirus *before* the lockdown. Hence, the synthetic control study approach is *by design* not well suited for studying the effects of lockdowns.

#### The role of optimal timing

One important thesis on the effect of lockdowns is that timing is important for a lockdown to be effective. The rationale behind this thesis is straightforward (assuming lockdowns are effective): If an epidemic is growing exponentially, the benefit of intervening sooner rather than later is disproportionately large. For example, if the doubling-time is one week, then locking down one week earlier will more than halve the total number of deaths, assuming that the pre-lockdown reproduction number is larger than two and *if* the lockdown brings the reproduction number below one. On the other hand, locking down too strongly and too early can result in a resurgence when restrictions are lifted if there is a failure to completely eliminate the virus, with potentially higher deaths than if it was permitted to spread to a small extent prior to the lockdown. Hence, the argument goes that there is an optimal timing of lockdowns.^32^

We, however, evaluate the general effect of lockdowns, i.e., whether lockdowns on average have been effective in reducing COVID-19 mortality. We therefore exclude studies which solely analyze the effect of optimally timed lockdowns in contrast to less well-timed lockdowns. There are several reasons for this exclusion.^33^

First, studies searching for the optimal timing of lockdowns will by design find inflated effects of the average lockdown, because they – if optimal timing is important – will neglect all the less well-timed lockdowns implemented around the world. And hence, these studies will not result in an unbiased estimate of the average effect of lockdowns.

Secondly, it is inherently difficult to differentiate between the effect of public awareness and the effect of lockdowns when looking at timing because people and politicians are likely to react to the same information. In fact, it is difficult for a democratic country’s political leaders to impose and enforce a lockdown, unless there is a widespread belief that a danger is imminent.

Björk et al. (2021) illustrate the difficulties in analyzing the effect of timing in Europe. They find that a 10-stringency-points-stricter lockdown would reduce COVID-19 mortality by a total of 200 deaths per million^34^ if done in week 11, 2020, but would only have approximately 1/3 of the effect if implemented one week earlier or later and close to no effect if implemented three weeks earlier or later. One interpretation of this result is that lockdowns do not work if people either find them unnecessary and fail to obey the mandatory restrictions or if people voluntarily lock themselves down. This is the argument Allen (2021) uses for the ineffectiveness of the lockdowns he identifies. If this interpretation is correct, what Björk et al. (2021) find is that information and signaling are far more important than the strictness of the lockdown. There may be other interpretations, but our point is that studies focusing on timing cannot differentiate between these two conflicting interpretations.

This view is also supported by Figure 7, which illustrates that all European countries and US states which were hit hard and early by COVID-19 in the spring of 2020 experienced high overall mortality rates, whereas all countries hit relatively late experienced low mortality rates. The figure shows that there is no doubt that being prepared for a pandemic and knowing when it arrives at your doorstep is vital. But to what degree this can be attributed to well-timed lockdowns or simply to alert citizens is a question, which is not easily answered and may previously have been misunderstood or neglected in prior research on e.g. the 1918 Spanish Flu pandemic (we will get back to this issue in section 5.2.4 on p. 71).

**Figure 7:**
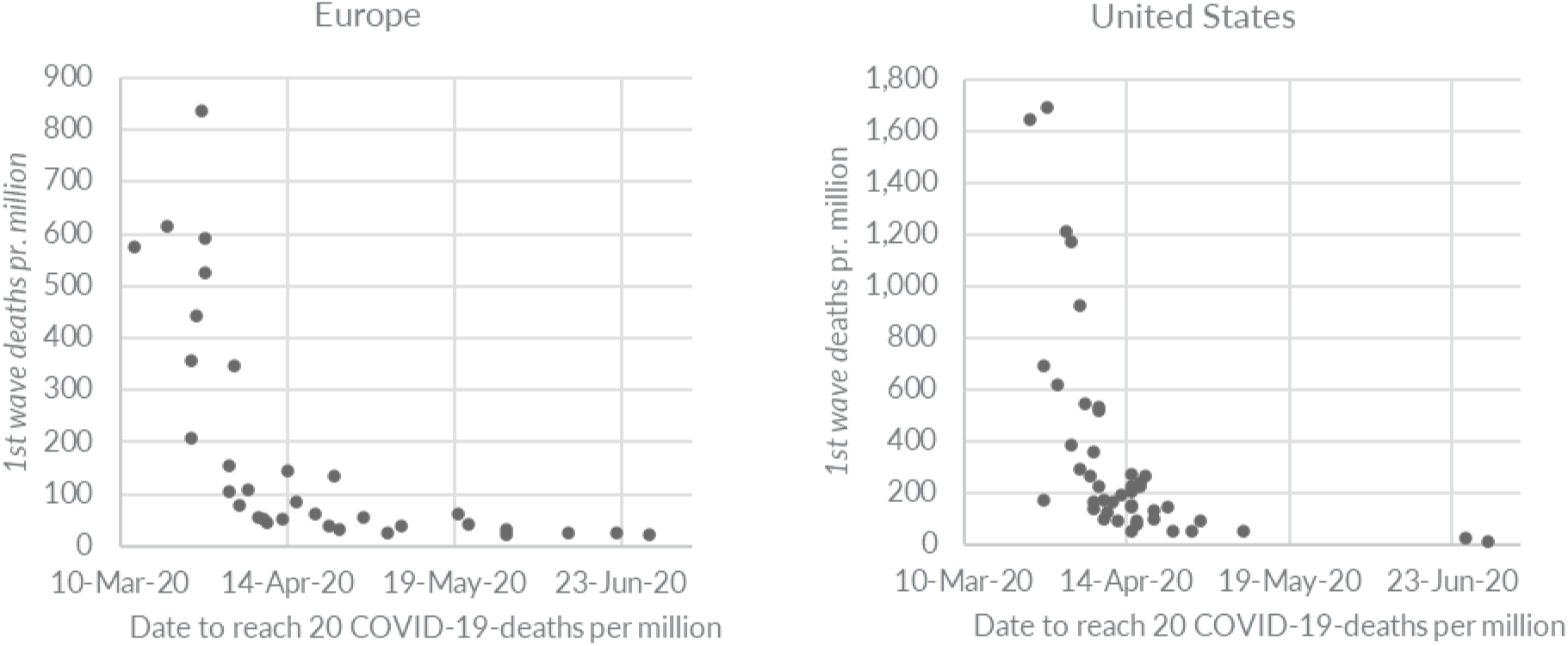
All countries and states that were hit late by the pandemic experienced lower COVID-19 mortality rates. Comment: The figure shows the relationship between early pandemic strength and total 1st wave of COVID-19 mortality. On the X-axis is “Days to reach 20 COVID-19-deaths per million (measured from February 15, 2020).” The Y-axis shows mortality (deaths per million) by June 30, 2020. Source: Reported COVID-19 deaths and OxCGRT stringency for European countries and US states with more than one million citizens. Data from Our World in Data (2022).

We are aware of three reviews (Iezadi et al. (2021), Perra (2020), and Stephens et al. (2020)), which opine on the importance of timing. Stephens et al. (2020) find 22 studies that look at policy and timing with respect to mortality rates, however, only four were multi-country, multi-policy studies, which could possibly account for the problems described above. Stephens et al. (2020) conclude that “the timing of policy interventions across countries relative to the first Wuhan case, first national disease case, or first national death, is not found to be correlated with mortality.” Iezadi et al. (2021) write that “it is very important to contain the spread of the infection at the very early stage of the outbreak. At later stages, no NPIs, even if implemented harshly, might be very effective”, while Perra (2020) writes that “countries that acted early, with respect to the local spread, were most successful in controlling the spread and reported markedly lower death tolls”. But these three reviews do not distinguish between the effect of information (which is the effect of being hit late by the pandemic) and the effect of lockdowns.

Although the verdict on optimal timing is still out, we would like to stress the importance of alternative interpretations here. As Figure 7 illustrates, one can easily interpret the lower mortality rates as an effect of early lockdowns even when it is caused by changes in voluntary behavior or – not unlikely – by a combination of both. One should be careful concluding that early lockdown is important when alternative conclusions – such as changing information and voluntary behavior – may explain outcomes equally well.

We further note that even if future research finds that the timing of lockdowns *is* crucial, such knowledge may not be useful for future policymakers. First, it is not easy to know when the right timing is. When COVID-19 hit Europe and the United States, it was virtually impossible to determine the right timing. The World Health Organization declared the COVID-19 pandemic a Public Health Emergency of International Concern (PHEIC) on January 30, 2020, WHO (2020a). However, this was the sixth PHEIC in just 11 years, and could not reasonably justify a lockdown.^35^ March 11, 2020, was the first time the WHO characterized COVID-19 as a pandemic, WHO (2020b). But at that date, Italy had already registered 13.7 COVID-19 deaths per million. On March 29, 2020, 18 days after the WHO declared the outbreak a pandemic and the earliest date that a lockdown response to the WHO’s announcement could potentially have a large effect due to the lag between infection and death, the mortality rate in Italy was a staggering 178 COVID-19 deaths per million, with an additional 13 per million dying each day.^36^

Second, we already pointed to the fact that policymakers (at least in democratic countries) need support from the electorate to impose and enforce lockdowns. So even if a few people know the right time to impose a lockdown, this information is only useful if citizens and politicians agree that there is a dangerous and threatening infectious disease, and act upon that threat.^37^ But under these conditions, they are also likely to respond significantly (and voluntarily) to recommendations, making a lockdown less necessary. In fact, data from the influenza surveillance program in Denmark from Statens Serum Institut (2020) show that the influenza vanished *before* lockdowns were implemented but possibly *coinciding with* the announcement of coming lockdowns which spurred significant voluntary behavioral changes. However, the influenza vanished at the exact same time in Norway and Sweden, as illustrated in Figure 8, suggesting that *if* lockdowns spurred significant voluntary behavioral changes, the Swedish 500-person limit on public gatherings effective by March 12, 2020, may have been sufficient to spur these changes.

**Figure 8:**
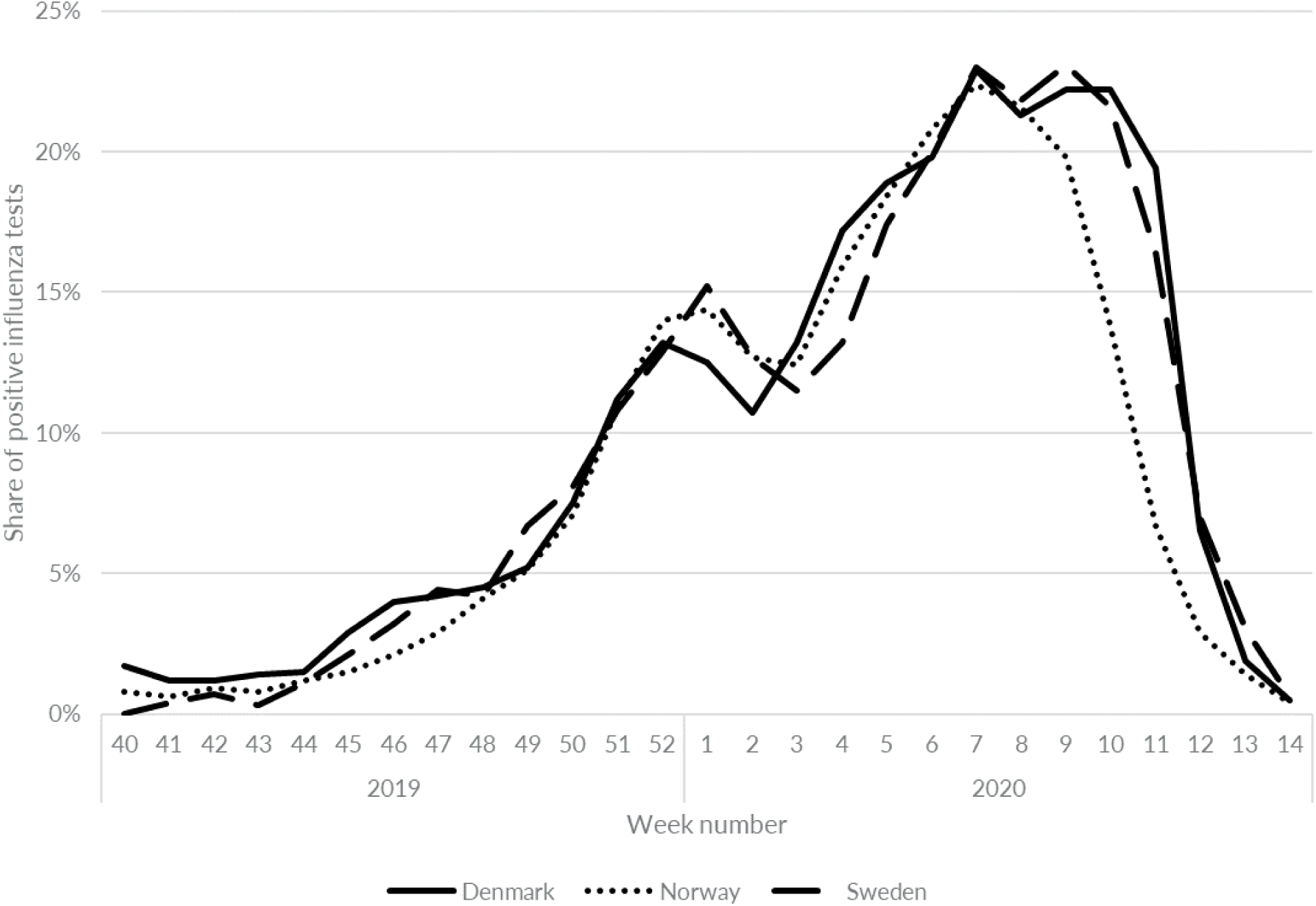
Influenza disappeared at the same time in Denmark, Norway, and Sweden in March 2020 despite radical differences in lockdown policies. Source: Data from Emborg, et al. (2021) Note: In Sweden, gatherings were limited to 500 persons from Thursday, March 12, 2020 (week 11), while high schools and higher education was closed from Wednesday, March 18, 2020 (week 12).

To conclude: We believe that most – if not all – studies focusing on timing fails to distinguish between the effects of lockdowns and the effect of voluntary behavioral changes.

## 3 The empirical evidence

In this section we present the empirical evidence found through our identification process. We describe the studies and their results. In addition, we comment on the methodology and possible identification problems and biases.

### 3.1 Preliminary considerations

Before we turn to the eligible studies, we present some considerations that we adopted when interpreting the empirical evidence.

#### The empirical interpretation is based on results – not the studies’ authors’ conclusions

While the policy conclusions contained in some studies are based on statistically significant results, many of these conclusions are ill-founded due to the tiny impact associated with the statistically significant results. For example, Ashraf (2020) states that “social distancing measures has proved effective in controlling the spread of [a] highly contagious virus.” However, their estimates show that the average lockdown in Europe and the U.S only reduced COVID-19 mortality by 2.4%.^38^

Another example is Chisadza et al. (2021). The authors argue that “less stringent interventions increase the number of deaths, whereas more severe responses to the pandemic can lower fatalities.” Their conclusion is based on a negative estimate for the squared term of *stringency* which results in a total negative effect on mortality rates (i.e., fewer deaths) for stringency values larger than 124. This means that for lockdowns with a stringency of at least 124, the lockdown theoretically reduces mortality. However, the stringency index is limited to values between 0 and 100 by design, so for all possible values of the stringency index, the total effect of lockdowns on mortality is positive (more deaths), so Chisadza et al.’s conclusion is infeasible.

This is illustrated in Figure 9 below. The figure describes the total policy effect based on Chisadza et al. (2021) estimates for their squared specification. Starting from a lockdown with a stringency of 0 (no lockdown) and increasing stringency from there, a stricter lockdown increases mortality. But, at stringency 62.4, a stricter lockdown reduces mortality at the margin. However, the total effect is still an increase in mortality for stringency values below 124. And because stringency values are capped at 100, there are no such lockdowns that decrease mortality overall.

**Figure 9:**
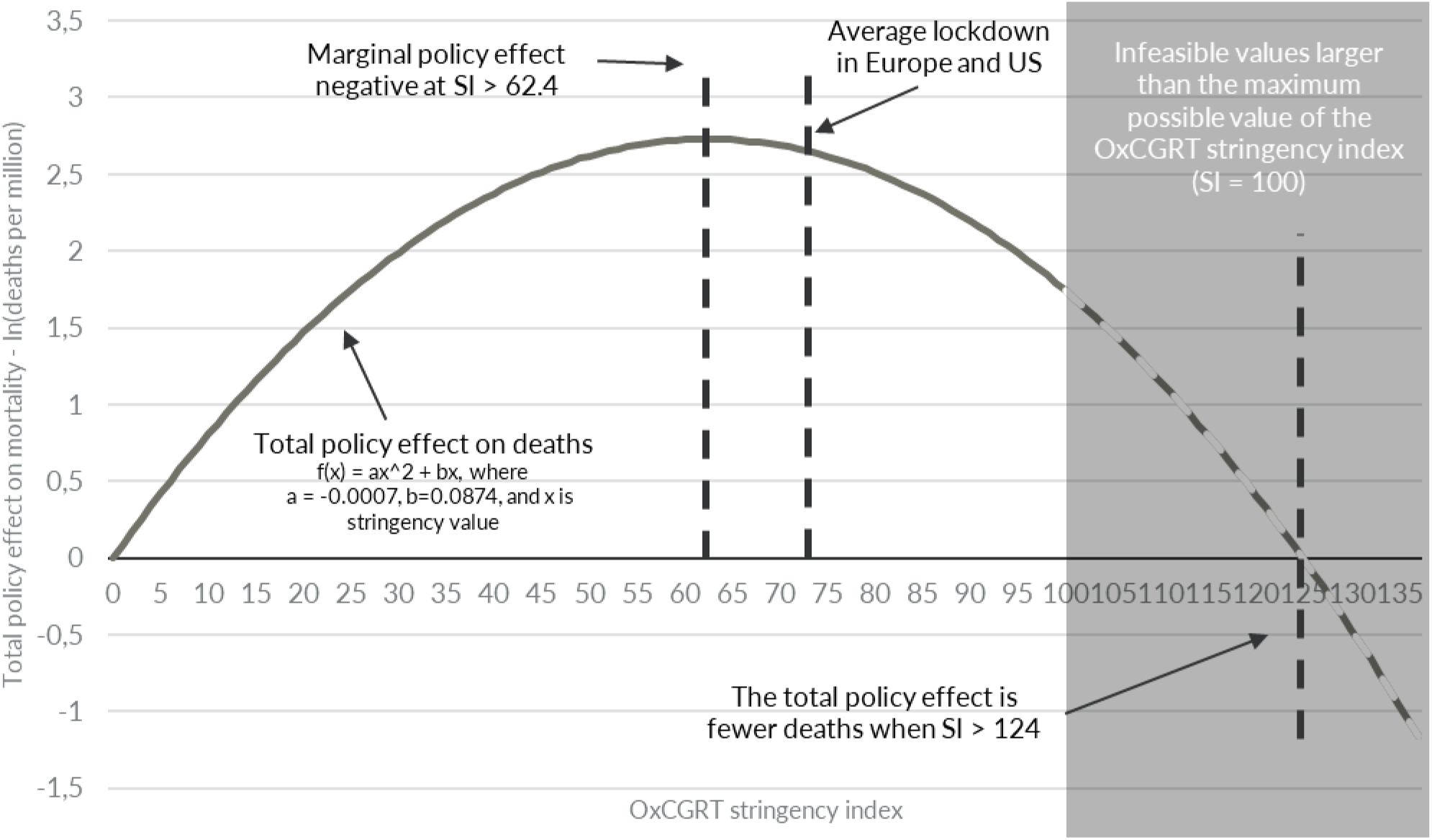
The total policy effect, including infeasible values outside the range of the OxCGRT stringency index, as estimated by Chisadza, et al. (2021) Source: Chisadza et al. (2021) Note: SI = OxCGRT stringency Index. The OxCGRT stringency Index measures the stringency of lockdowns on a scale from 0 to 100, where a higher value means stricter lockdowns.

Again, to avoid any such biases, we base our interpretations solely on the empirical estimates and not on the authors’ own interpretation of their results.

#### Handling multiple models, specifications, and uncertainties

Several studies adopt a number of models to understand the effect of lockdowns. For example, Bjørnskov (2021a) estimates the effect after one, two, three, and four weeks of lockdowns. For these studies, we select the longest time horizon analyzed to obtain the estimate closest to the long-term effect of lockdowns.

Several studies also use multiple specifications, including and excluding potentially relevant variables. For these studies, we choose the model which the authors regard as their main specification.

Finally, some studies have multiple models which the authors regard as equally important. One interesting example is Chernozhukov et al. (2021), who estimate two models with and without national case numbers as a variable. They show that including this variable in their model substantially reduces the efficacy of lockdowns on mortality. The explanation could be that people responded to information about national conditions. For these studies, we present both estimates in Table 2, but – following Doucouliagos and Paldam (2008) – we use an average of the estimates in our meta-analysis to avoid giving more weight to a study with multiple models relative to studies with just one principal model (note that for Chernozhukov et al. (2021), we can only include the model estimating large effects of lockdowns, because the report the counterfactual effect for this model only – see Table 19).

**Table 2:** Summary of eligible studies.

| 1. Study (Author & title) | 2. Measure | 3. Description | 4. Results | 5. Comments |
| --- | --- | --- | --- | --- |
| Alderman and Harjoto (2020); "COVID-19: U.S. shelter-in-place orders and demographic characteristics linked to | COVID-19 mortality | The study uses state-level data from all U.S. states published by the COVID-19 Tracking Project, and a multivariate regression analysis to empirically investigate the impacts of the duration of shelter-in-place orders on mortality. | The study finds that shelter-in-place orders are - for the average duration - associated with 1% (insignificant) fewer deaths per capita. | The study does not use a stringent difference-in-difference approach; but this approach is included as the study uses panel data with a time dimension. |

<sup>40</sup> The following numbers are based on data presented in Table 4.
<sup>41</sup> Yue Li et al. (2021) use "data on the stringency of state social distancing measures from wallethub.com".

| 1. Study (Author & title) | 2. Measure | 3. Description | 4. Results | 5. Comments |
| --- | --- | --- | --- | --- |
| cases, mortality, and recovery rates" |  |  |  |  |
| An et al. (2021); "Policy Design for COVID-19: Worldwide Evidence on the Efficacies of Early Mask Mandates and Other Policy Interventions" | COVID-19 deaths | The authors use 164 countries to estimate the long run efficacy of early mandate adoption. They use both a country fixed-effects model and a country random effects model. They also use several other methods in order to estimate the specific NPI's effect on infection rates and mortality, and they look specifically at timing for each NPI. | With different modeling approaches and alternative measurements (for both the focal independent and dependent variables), the analysis shows that domestic lockdowns and restaurant closures do not display any consistent associations with new infection and mortality rates in the short term. Mass gathering bans and school closures need more time to manifest their short-term efficacies. Both cross-sectional and longitudinal analyses provide consistent evidence that only mask mandates demonstrate persistent long-run efficacy from early adoption. | The study uses several different methods to estimate how effective timing and specific NPIs are and finds that the only NPI that shows consistent reduction of infection rates and mortality both in the short and long term is mask mandates. |
| Ashraf (2020); "Socioeconomic conditions, government interventions and health outcomes during COVID-19" | COVID-19 mortality | The study's focus is on the effectiveness of policies targeted to diminish the effect of socioeconomic inequalities (economic support) on COVID-19 deaths. The study uses data from 80 countries worldwide and includes the OxCGRT stringency as a control variable in its models. The paper finds a significant negative (fewer deaths) effect of stricter lockdowns. The effect of lockdowns becomes insignificant when the author includes an interaction term between the socioeconomic conditions index and the economic support index in the study's model. | For each 1-unit increase in OxCGRT stringency index, the cumulative mortality changes by -0.326 deaths per million (fewer deaths). The estimate is -0.073 deaths per million but becomes insignificant when including an interaction term between the socioeconomic conditions index and the economic support index. |  |
| Berry et al. (2021); "Evaluating the effects of shelter-in-place policies during the COVID-19 pandemic" | COVID-19 mortality | The authors use U.S. county data on COVID-19 deaths from Johns Hopkins University and SIPO data from the University of Washington to estimate the effect of SIPOs. They find no detectable effects of SIPOs on deaths. The authors stress that their findings should not be interpreted as evidence that social distancing behaviors are not effective. Many people had already changed their behaviors before the introduction of shelter-in-place orders, and shelter-in-place orders appear to have been ineffective precisely because they did not meaningfully alter social distancing behavior. | A SIPO increases the number of deaths by 0,654 per million after 14 days (see Fig. 2) | The authors conclude that "We do not find detectable effects of these policies [SIPO] on disease spread or deaths." However, this statement does not correspond to their results. In figure 2 they show that the effect on deaths is significant after 14 days. The paper looks at the effect 14 days after SIPOs are implemented, which is a short lag given that the time between infection and deaths is at least 2-3 weeks. |
| Bjørnskov (2021a); "Did Lockdown Work? An Economist's Cross-Country Comparison" | Excess mortality | The study uses excess mortality and OxCGRT stringency data from 24 European countries to estimate the effect of lockdowns on the number of deaths in the subsequent one, two, three and four weeks. | A stricter lockdown (OxCGRT stringency) does not have a significant effect on excess mortality. Specifications using instrument variables yields similar results. | The study finds a positive (more deaths) effect after one and two weeks, which could indicate that other factors (omitted variables) affect the results. |
| Blanco et al. (2020); "Do Coronavirus Containment Measures Work? Worldwide Evidence" | COVID-19 mortality | The study uses data for deaths and NPIs from Hale et al. (2020) covering 158 countries between January and August 2020 to evaluate the effect of eight different NPIs: stay at home orders, bans on gatherings, bans on public events, school closures, lockdowns of workplaces, interruption of public transportation services, and international border closures. They address the possible endogeneity of the NPIs by using instrumental variables. | When using the naïve dummy variable approach, all parameters are statistically insignificant. On the contrary, estimates using the instrumental variable approach indicate that NPIs are effective in reducing the growth rate in the daily number of deaths 14 days later. | The study runs the same model four times for each of the different NPIs (stay at home orders, bans on meetings, bans on public events, and mobility restrictions). These NPIs were often introduced almost simultaneously so there is a high risk of multicollinearity with each run capturing the same underlying effect. Indeed, the size and standard errors of the estimates are worryingly similar. The study looks at the effect 14 days after NPIs are implemented, which is a fairly short lag given that the time between infection and deaths is 2-3 weeks; see e.g., Flaxman et al. (2020), which according to Bjørnskov (2020) appears to be the minimum typical time from infection to death. |
| Bonardi et al. (2020); "Fast and local: How did lockdown policies affect the spread and severity of the covid-19" | Growth rates | The study uses NPI data collected from news headlines from LexisNexis and death data from Johns Hopkins University up to April 1st, 2020 in a panel structure with 184 countries. The study controls for country fixed-effects, day fixed-effects, and within-country evolution of the disease. | The study finds that certain interventions (SIPO, regional lockdown and partial lockdown) work (in developed countries), but that stricter interventions (SIPOs) do not have a larger effect than less strict interventions (e.g., restrictions on gatherings). It finds no effect of border closures. | The study finds a positive (more deaths) effect on day 1 after lockdown which may indicate that their results are driven by other factors (omitted variables). We rely on their publicly available version submitted to CEPR Covid Economics, but estimates on the effect of deaths can be found in Supplementary material, which is available in an updated version hosted on the Danish Broadcasting Corporation's webpage: https://www.dr.dk/static/documents/2021/03/04/managing_pandemics_e3911c11.pdf |
| Chernozhukov et al. (2021); "Causal impact of masks, policies, behavior on early covid-19 pandemic in the U.S." | Growth rates | The study uses COVID-deaths from the New York Times, Johns Hopkins University, and data on U.S. states' policies from Raifman et al. (2020) to estimate the effect of SIPOs, closed nonessential businesses, closed K-12 schools, closed restaurants except takeout, closed movie theaters, and face mask mandates for employees in public facing businesses. | The study finds that mandatory masks for employees and closing K-12 schools reduces deaths. SIPO and closing business (average of closed businesses, restaurants and movie theaters) have no statistically significant effect. The effect of school closures is highly sensitive to the inclusion of national case and death data. | The authors state that "our regression specification for case and death growths is explicitly guided by an SIR model although our causal approach does not hinge on the validity of an SIR model." We are uncertain if this means that data are managed to fit an SIR-model (and thus should fail our eligibility criteria). |
| Chisadza et al. (2021); "Government Effectiveness and the COVID-19 Pandemic" | COVID-19 mortality | The study uses COVID-19-deaths and OxCGRT stringency from 144 countries to estimate the effect of lockdowns on the number of COVID-19-deaths. It finds a significant positive (more deaths) non-linear association between government response indices and the number of deaths. | In the authors' linear model, an increase by 1 on "stringency index" increases the number of log deaths by 0.0130 per million (corresponding to 1.3%). In their non-linear model, the sign of the squared term is negative, but the combined non-linear estimate is positive (increases deaths) and larger than the linear estimate for all values of the OxCGRT stringency index. | The authors state that "less stringent interventions increase the number of deaths, whereas more severe responses to the pandemic can lower fatalities." However, according to their estimates this is not correct, as the combined non-linear estimate cannot be negative for relevant values of the OxCGRT stringency index (0 to 100). |
| Clyde et al. (2021); "A Study of the Effectiveness of Governmental Strategies" | COVID-19 mortality | The study uses data for NPIs in 37 OECD countries from the Oxford COVID-19 Government Response Tracker (OxCGRT) to examine the impact of policy variable | Results indicate that only school closings and public transportation closings have a persistently significant |  |
| for Managing Mortality from COVID-19" |  | changes between March 15 and October 31, 2020. | impact. Stay-at-home policies only show a significant impact after 70 days. Workplace closings, restrictions on the size of gatherings, and restrictions on internal travel show no significant impact on mortality rates. Moreover, stricter measures are not significantly associated with lower growth rates in mortality. |  |
| Dave et al. (2021); "When Do Shelter-in-Place Orders Fight Covid-19 Best? Policy Heterogeneity Across States and Adoption Time" | COVID-19 mortality | The study uses smartphone location tracking and state data on COVID-19 deaths and SIPO data (supplemented by their own searches) collected by the New York Times to estimate the effect of SIPOs. The authors find that a SIPO was associated with a 9%-10% increase in the rate at which state residents remained in their homes full-time, but overall, they do not find a significant effect on mortality after 20+ days (see Figure 4). They indicate that the lacking significance may be due to long term estimates being identified of a few early adopting states. | The study finds no overall significant effect of a SIPO on deaths but does find a negative effect (fewer deaths) in early adopting states. | The study finds large effects of a SIPO on deaths after 6-14 days in early adopting states (see Table 8), which is before a SIPO-related effect would be seen. This could indicate that other factors rather than SIPOs drive the results. |
| Dergiades et al. (2020); "Effectiveness of government policies in response to the COVID-19 outbreak" | COVID-19 mortality | The study uses daily deaths from the European Centre for Disease Prevention and Control and OxCGRT stringency from 32 countries worldwide (including U.S.) to estimate the effect of lockdowns on the number of deaths. | The study finds that the greater the strength of government interventions at an early stage, the more effective these are in slowing down or reversing the growth rate of deaths. | The focus is on the effect of early stage NPIs and thus does not absolutely live up to our eligibility criteria. However, we include the study as it differentiates between lockdown strength at an early stage. |
| Ertem et al. (2021); "The impact of school opening model on SARS-CoV-2 community incidence and mortality" | COVID-19 deaths | The study uses 459 U.S. counties to estimate the impact of school opening on mortality. It uses multivariate Poisson regressions with robust standard errors. The authors define 3 school modes, as they focus on the difference between traditional and virtual. | "There was no impact of school opening mode on subsequent COVID-19-related deaths during the entire 12-week period after school opening in any region (Fig. 4 and Extended Data Fig. 2)." The authors also conclude that there are major limitations because the underlying reason for regional differences could not be delineated. |  |
| Fakir and Bharati (2021); "Pandemic catch-22: The role of mobility restrictions and institutional inequalities in halting the spread of COVID-19" | COVID-19 mortality | The study uses data from 127 countries, combining high-frequency measures of mobility data from Google's daily mobility reports, country-date-level information on the stringency of restrictions in response to the pandemic from Oxford's Coronavirus Government Response Tracker (OxCGRT), and daily data on deaths attributed to COVID-19 from Our World in Data and Johns Hopkins University. The study instruments stringency using day-to-day changes in the stringency of the restrictions in the rest of the world. | The study finds large causal effects of stricter restrictions on the weekly growth rate of recorded deaths attributed to COVID-19. It shows that more stringent interventions help more in richer, more educated, more democratic, and less corrupt countries with older, healthier populations and more effective governments. | The authors find a larger effect on deaths after 0 days than after 14 and 21 days (Table 3). This is surprising given that it takes 2-3 weeks from infection to death, and it may indicate that their results are driven by other factors. |
| Fowler et al. (2021); "Stay-at-home orders associate with subsequent decreases in COVID-19 cases and fatalities in the United States" | COVID-19 mortality | The study uses U.S. county data on COVID-19 deaths and SIPO data collected by the New York Times to estimate the effect of SIPO's using a two-way fixed-effects difference-in-differences model. It finds a large and early (after few days) effect of SIPOs on COVID-19 related deaths. | Stay-at-home orders are also associated with a 59.8 percent (18.3 to 80.2) average reduction in weekly fatalities after three weeks. These results suggest that stay-at-home orders might have reduced confirmed cases by 390,000 (the 95% confidence interval spans from 170,000 to 680,000) and fatalities by 41,000 (from 27,000 to 59,000) within the first three weeks in localities that implemented stay-at-home orders. | The study finds the largest effect of SIPOs on deaths after 10 days (see Figure 4), before a SIPO-related effect could possibly be seen as it takes 2-3 weeks from infection to death. This could indicate that other factors drive their results. |
| Fuller et al. (2021); "Mitigation Policies and COVID-19-Associated Mortality – 37 European Countries, January 23–June 30, 2020" | COVID-19 mortality | The study uses COVID 19-deaths and OxCGRT stringency in 37 European countries to estimate the effect of lockdowns on the number of COVID-19 deaths. It finds a significant negative (fewer deaths) effect of stricter lockdowns after the mortality threshold is reached. The threshold is a daily rate of 0.02 new COVID-19 deaths per 100,000 population, based on a 7-day moving average. | For each 1-unit increase in OxCGRT stringency index, the cumulative mortality decreases by 0.55 deaths per 100,000. |  |
| Gibson (2020); "Government mandated lockdowns do not reduce Covid-19 deaths: implications for evaluating the stringent New Zealand response" | COVID-19 mortality | The study uses data for every county in the United States from March through June 1, 2020, to estimate the effect of SIPOs (called "lockdowns") on COVID-19 mortality. Policy data are acquired from American Red Cross reporting on emergency regulations. The author's control variables include county population and density, the elder share, the share in nursing homes, nine other demographic and economic characteristics and a set of regional fixed-effects. The author handles causality problems using instrument variables (IV). | The author finds no statistically significant effect of SIPOs. | The author uses the word "lockdown" as synonym for SIPO (writes "technically, government-ordered community quarantine") |
| Goldstein et al. (2021); "Lockdown Fatigue: The Diminishing Effects of Quarantines on the Spread of COVID-19 " | COVID-19 mortality | The study uses panel data from 152 countries with data from the onset of the pandemic until December 31, 2020. It finds that lockdowns tend to reduce the number of COVID-19 related deaths, but also that this benign impact declines over time: after four months of strict lockdown, NPIs have a significantly weaker contribution in terms of their effect in reducing COVID-19 related fatalities. | Stricter lockdowns reduce deaths for the first 60 days, whereafter the cumulative effect begins to decrease. If reintroduced after 120 days, the effect of lockdowns is smaller in the short run, but after 90 days the effect is almost the same as during the first lockdown (only approximately 10% lower). | There is little documentation in the study (e.g., no tables with estimates). |
| Guo et al. (2021); "Mitigation Interventions in the United States: An Exploratory Investigation of Determinants and Impacts" | COVID-19 mortality | The study uses policy data of 1,470 executive orders from the state-government websites for all 50 U.S. states and Washington DC and COVID-19 deaths from Johns Hopkins University in a random-effect spatial error panel model to estimate the effect on COVID-19 deaths of nine NPIs: SIPO, strengthened | Two mitigation strategies (all school closure and mandatory self-quarantine of travelers) showed a positive (more deaths) impact on COVID-19-deaths per 10,000. Six mitigation strategies (SIPO, | The study only concludes on NPIs which reduce mortality. However, the conclusion is based on one-tailed tests, which means that all positive estimates (more deaths) are deemed insignificant. Thus, in their mortality-specification (Table 3, Proportion of Cumulative Deaths Over the Population), the estimate of all school closures (.204) and |
| | | SIPO, public school closure, all school closure, large-gathering ban of more than 10 people, any gathering ban, restaurant/bar limit to dining out only, nonessential business closure, and mandatory self-quarantine of travelers. | public school closure, large gathering bans [$>10$], any gathering ban, restaurant/bar limit to dining out only, and nonessential business closure) did not show any impact (Table 3, "Proportion of Cumulative Deaths Over the Population). | mandatory self-quarantine of travelers (0.363) is deemed insignificant based on schools CI [.029, .379] and quarantine CI [.193, .532]. We believe these results should be interpreted as a significant increase in mortality and that these results should have been part of their conclusion. |
| Hale et al. (2020); "Global assessment of the relationship between government response measures and COVID-19 deaths" | COVID-19 mortality | The study uses the OxCGRT stringency and COVID-19-deaths from the European Centre for Disease Prevention and Control for 170 countries. It estimates both cross-sectional models in which countries are the unit of analysis, as well as longitudinal models on time-series panel data with country-day as the unit of analysis (including models that use both time and country fixed-effects). | The study finds that higher stringency in the past leads to a lower growth rate in the present, with each additional point of stringency corresponding to a 0.039%-point reduction in daily deaths growth rates six weeks later. |  |
| Hale, Angrist, Hale, et al. (2021); "Government responses and COVID-19 deaths: Global evidence across multiple pandemic waves" | COVID-19 mortality | The study uses the OxCGRT stringency to analyze the effect of stricter lockdowns in 113 countries through 1-3 waves. | The study finds that stricter lockdowns reduce mortality and that the effect was particularly large – even during the first wave – in countries which experienced three waves. | The authors' results on three wave countries are based on just 10 countries. However, since there is no obvious reason why lockdowns in countries, which later experienced a third wave, should be particularly effective, this could indicate that some model misspecification is driving the results. |
| Leffler et al. (2020); "Association of country-wide coronavirus mortality with demographics, testing, lockdowns, and public wearing of masks" | COVID-19 mortality | The study uses COVID-19 deaths from Worldometer and info about NPIs (mask/mask recommendations, international travel restrictions and lockdowns [defined as any closure of schools or workplaces, limits on public gatherings or internal movement, or stay-at-home orders]) from Hale et al. (2020) for 200 countries to estimate the effect of the duration of NPIs on the number of deaths. | The study finds that masking (mask recommendations) reduces mortality. For each week that masks were recommended the increase in per-capita mortality was 8.1% (compared to 55.7% increase when masks were not recommended). It finds no significant effect of the number of weeks with internal lockdowns and international travel restrictions (Table 2). | The authors' "mask recommendation" category includes some countries where masks were mandated (see Supplemental Table A1) and may (partially) capture the effect of mask mandates. The study looks at duration, which may cause a causality problem, because politicians may be less likely to ease restrictions when there are many cases or deaths. |
| Yue Li et al. (2021); "Association of state social distancing restrictions with nursing home COVID-19 and non-COVID-19 outcomes" | COVID-19 mortality | The study uses data on the stringency of state social distancing measures from wallethub.com (based on 17 state COVID-19 policy metrics) to categorize US states into either "high" or "low" stringency. The study then uses linear regression to estimate cumulative numbers of deaths among residents in long-term care in low and high stringency states based on mortality data from 14,046 nursing homes. | The study finds that stricter lockdowns reduced COVID-19 mortality and across-facility disparities in COVID-19 outcomes, but also caused more deaths due to non-COVID reasons among nursing home residents. |  |
| Mccafferty and Ashley (2021); "Covid-19 Social Distancing Interventions by Statutory Mandate and Their Observational Correlation to Mortality in the United States and Europe" | Other | The study uses data from 27 U.S. states and 12 European countries to analyze the effect of NPIs on peak mortality rate using general linear mixed-effects modelling. | The study finds that no mandate (school closures, prohibition on mass gatherings, business closures, stay at home orders, severe travel restrictions, and closure of non-essential businesses) |  |
|  |  |  | was effective in reducing the peak COVID-19 mortality rate. |  |
| Pan et al. (2020); "Covid-19: Effectiveness of non-pharmaceutical interventions in the United States before phased removal of social distancing protections varies by region" | COVID-19 mortality | The study uses county-level data for all U.S. states. Mortality data are obtained from Johns Hopkins University, while policy data are obtained from official governmental websites. It categorizes 12 policies into 4 levels of disease control; Level 1 (low) - State of Emergency; Level 2 (moderate) - school closures, restricting access (visits) to nursing homes, or closing restaurants and bars; Level 3 (high) - non-essential business closures, suspending non-violent arrests, suspending elective medical procedures, suspending evictions, or restricting mass gatherings of at least 10 people; and Level 4 (aggressive) - sheltering in place or stay-at-home, public mask requirements, or travel restrictions. It uses a stepped-wedge cluster randomized trial (SW-CRT) for clustering and negative binomial mixed model regression. | The study concludes that only (duration of, see comment in next column) level 4 restrictions are associated with reduced risk of death, with an average 15% decline in the COVID-19 death rate per day. Implementation of level 3 and level 2 restrictions increased death rates in 6 of 6 regions, while longer duration increased death rates in 5 of 6 regions. | The authors focus on the negative estimate of duration of Level 4. However, their implementation estimate is large and positive, and the combined effect of implementation and duration is unclear. |
| Pincombe et al. (2021); "The effectiveness of national-level containment and closure policies across income levels during the COVID-19 pandemic: an analysis of 113 countries" | COVID-19 mortality | The study uses daily data for 113 countries on cumulative COVID-19 death counts over 130 days between February 15, 2020, and June 23, 2020, to examine changes in mortality growth rates across the World Bank's income group classifications following shelter-in-place recommendations or orders (the authors use one variable covering both recommendations and orders). | The study finds that shelter-in-place recommendations or orders reduces mortality growth rates in high income countries (although insignificant) but increases growth rates in countries in other income groups. |  |
| Sears et al. (2020); "Are we #stayinghome to Flatten the Curve?" | COVID-19 mortality | The study uses cellular location data from all 50 states and the District of Columbia to investigate mobility patterns during the pandemic across states and time. The authors estimate the effect of stay-at-home policies on COVID-19 mortality by adding COVID-19 death tolls and the timing of SIPOs for each state.<br>"We obtain travel activity and social distancing data from the analytics company Unacast."<br>"To denote periods before or after a state implemented a 'stay at home order,' we obtain the date each statewide policy was issued [45] for all fifty states and the District of Columbia" – from the New York Times. | The study finds that SIPOs lower deaths by 0.13- 0.17 per 100,000 residents, equivalent to death rates 29-35% lower than in the absence of policies. However, these estimates are insignificant at a 95% confidence interval (see Table 4). The study also finds reductions in activity levels prior to mandates. The human encounter rate fell by 63 percentage points and nonessential visits by 39 percentage points relative to pre-COVID-19 levels, prior to any state implementing a statewide mandate. | In the abstract the authors state that death rates would be 42-54% lower than in the absence of policies. However, this includes averted deaths due to pre-mandate social distancing behavior (p. 6). The effect of a SIPO is a reduction in deaths by 29%-35% compared to a situation without a SIPO but with pre-mandate social distancing. These estimates are insignificant at a 95% confidence interval. |
| Shiva and Molana (2021); "The Luxury of Lockdown" | COVID-19 mortality | The study uses COVID-19-deaths and OxCGRT stringency from 169 countries to estimate the effect of a lockdown on the number of deaths 1-8 weeks later. It finds that stricter lockdowns reduce COVID-19-deaths 4 weeks later (but insignificantly after 8 weeks) and have the greatest effect in high income countries. It finds no effect of workplace closures in low-income countries. | A stricter lockdown (1 stringency point) reduces deaths by 0.1% after 4 weeks. After 8 weeks the effect is insignificant. |  |
| Spiegel and Tookes (2021a); "All or Nothing? Partial Business Shutdowns and COVID-19 Fatality Growth" | Growth rates | The study uses hand-collected policy data for all US counties from March through December 2020 to estimate the effect of capacity limits on spas, bars, restaurants, and gyms. | The study finds mixed results, where partial capacity restrictions on restaurants and bars are more effective than full shutdowns, and full shutdowns of gyms reduce fatality growth rate, while partial capacity restrictions are counterproductive. |  |
| Spiegel and Tookes (2021b); "Business restrictions and Covid-19 fatalities" | COVID-19 mortality | The study uses data for every county in the United States from March through December 2020 to estimate the effect of various NPIs on the COVID-19 deaths growth rate. Derives causality by 1) assuming that state regulators primarily focus on the state's most populous counties, so state regulation in smaller counties can be viewed as a quasi-randomized experiment, and 2) conducting county pair analysis, where similar counties in different states (and subject to different state policies) are compared. | The study finds that some interventions (e.g., mask mandates, restaurant and bar closures, gym closures, and high-risk business closures) reduces mortality growth, while other interventions (closures of low- to medium-risk businesses and personal care/spa services) did not have an effect and may even have increased the number of deaths. | In total the authors analyze the lockdown effect of 21 variables. 14 of 21 estimates are significant, and of these 6 are negative (reduces deaths) while 8 are positive (increases deaths). Some results are not readily intuitive; e.g., mask recommendations increase deaths by 48% while mask mandates reduce deaths by 12%, and closing restaurants and bars reduces deaths by 50%, while closing bars but not restaurants only reduces deaths by 5%, although the latter could potentially be explained by more crowding in open venues. They handle early effect (within 1-2 weeks) in Table 5. |
| Stokes et al. (2020); "The relative effects of non-pharmaceutical interventions on early Covid-19 mortality: natural experiment in 130 countries" | COVID-19 mortality | The study uses daily Covid-19 deaths for 130 countries from the European Centre for Disease Prevention and Control (ECDC) and daily policy data from the Oxford COVID-19 Government Response Tracker (OxCGRT). It looks at all levels of restrictions for each of the nine sub-categories of the OxCGRT stringency index: school, work, events, gatherings, transport, SIPO, internal movement, and travel. | Of the nine sub-categories in the OxCGRT stringency index, only travel restrictions are consistently significant (with level 2 "Quarantine arrivals from high-risk regions" having the largest effect, and the strictest level 4 "Total border closure" having the smallest effect). Restrictions on very large gatherings (>1,000) have a large significant negative (fewer deaths) effect, while the effect of stricter restrictions on gatherings are insignificant. The authors recommend that the closing of schools (level 1) has a very large (in absolute terms it is twice the effect of border quarantines) positive effect (more deaths) while stricter interventions on schools have no significant effect. Required cancelling of public events also has a significant positive (more deaths) effect. We focus on their 14-38 days results, as they catch the longest time frame (the authors' 0-24- | The authors' results are counter intuitive and somewhat inconclusive. Why does limiting very large gatherings (>1,000) work, while stricter limits do not? Why does recommending school closures cause more deaths? Why is the effect of border closures before the first death insignificant, while the effect of closing borders after the first death is significant (and large)? Why does quarantining arrivals from high-risk regions work better than total border closures? With 23 estimated parameters in total, these counter intuitive and inconclusive results could be caused by multiple test bias (we correct for this in the meta-analysis) but may also be caused by other factors such as omitted variable bias. |
|  |  |  | day model returns mostly insignificant results). |  |
| Yang et al. (2021); "What is the relationship between government response and COVID-19 pandemics? Global evidence of 118 countries" | COVID-19 deaths | The study uses OxCGRT stringency covering 118 countries from January 1 to April 13, 2021 to estimate the effect of lockdowns on mortality. | The study finds a statistical negative (fewer deaths) effect of stricter lockdowns on COVID-19 mortality rates 21 days later. | The study finds a (relatively large) effect after just seven days before any policy could be effective, which could indicate that other factors (omitted variables) affect the results. |

*Note: All comments on the significance of estimates are based on a 5% significance level unless otherwise stated.*

For studies which look at different classes of countries (e.g., rich and poor), we report both estimates in Table 2 but use the estimate for rich Western countries in our meta-analysis, where we derive standardized estimates for Europe and the United States.

#### Effects are measured relative to “doing the least in the spring of 2020”

Virtually all countries in the world implemented some kind of lockdown in response to the COVID-19 pandemic. Hence, most estimates are relative to “doing the least,” which in many Western countries means relative to doing as Sweden did during the first wave, when Sweden, due to constitutional constraints,^39^ implemented very few restrictions compared to other western countries. However, some studies state that they *do* compare the effect of doing something to the effect of doing absolutely nothing (e.g. Bonardi et al. (2020)).

The consequence is that some estimates are relative to “doing the least” while others are relative to “doing nothing.” This may lead to biases if “doing the least” works as a signal (or warning) which alters the behavior of the public. For example, Gupta et al. (2020) find a large effect of emergency declarations, which they argue “are best viewed as an information instrument that signals to the population that the public health situation is serious and they act accordingly”, on social distancing but not of other policies such as SIPOs. Thus, if we compare a country issuing a SIPO to a country doing nothing, we may overestimate the effect of a SIPO, because it is the sum of the signal *and* the SIPO. Instead, we should compare the country issuing the SIPO to a country “doing the least” to estimate the *marginal* effect of the SIPO.

To take an example, Bonardi et al. (2020) find relatively large effects of doing *something* but no effect of doing *more*. They find no extra effect of stricter lockdowns relative to less strict lockdowns and state that “our results point to the fact that people might adjust their behaviors quite significantly as partial measures are implemented, which might be enough to stop the spread of the virus”. Hence, whether the baseline is “doing the least”, or the baseline is “doing nothing” can affect the magnitude of the estimated impacts. There is no obvious right way to resolve this issue, but since estimates in most studies are relative to doing less, we report results as compared to “doing the least” when available. Hence, for Bonardi et al. (2020), we state that the effect of lockdowns is zero (compared to “doing the least”).

Note that this also means that our results cannot say much about the importance of *signaling*. One could imagine that lockdown may serve as a signal to citizens that now is the time to be careful. If signaling is thought to be important, future research should focus on finding the least costly signals.

### 3.2 Overview of the findings of eligible studies

Table 2 covers the 32 studies eligible for our review.^40^ Out of these 32 studies, 19 were peer-reviewed and 13 were working papers. The studies analyze lockdowns during the first wave. Most of the studies (25) use data collected before September 1^st^, 2020, and 6 use data collected before May 1^st^, 2020. Only two studies use data collected after January 1^st^, 2021. All studies are cross-sectional, ranging across jurisdictions. Geographically, 15 studies cover countries worldwide, two cover European countries, 13 cover the United States, one covers Europe and the United States, and one covers the OECD member countries. Seven studies analyze the effect of SIPOs, 11 studies analyze the effect of stricter lockdowns (measured by the OxCGRT stringency index), 13 studies analyze specific NPIs independently, and one study analyzes other measures.^41^

Several studies find no statistically significant effect of lockdowns on mortality. This includes Bjørnskov (2021a) and Goldstein et al. (2021), who find no significant effect of stricter lockdowns (higher OxCGRT stringency index), Sears et al. (2020) and Dave et al. (2021), who find no significant effect of SIPOs, and An et al. (2021) and Guo et al. (2021) who find no significant negative (fewer deaths) effect of any of the analyzed NPIs, including business closures, school closures, and border closures.

Other studies find a significant *negative* relationship between lockdowns and mortality. Fowler et al. (2021) conclude that SIPOs reduce COVID-19 mortality by 35%, while Chernozhukov et al. (2021) state that employee mask mandates reduces mortality by 34% and closing businesses and bars reduces mortality by 29%.

A few studies find a significant *positive* relationship between lockdowns and mortality. This includes Chisadza et al. (2021), who find that stricter lockdowns (higher OxCGRT stringency index) increases COVID-19 mortality and Berry et al. (2021), who find that SIPOs increase COVID-19 mortality by 1% after 14 days.

Most studies use the number of official COVID-19 deaths as the dependent variable. Only one study, Bjørnskov (2021a), looks at total excess mortality which we believe to be the best (albeit still imperfect) measure, as it overcomes the measurement problems related to proper reporting of COVID-19 deaths.

It is difficult to draw any firm conclusions based on the overview presented in Table 2. For example, is −0.073 to −0.326 deaths/million per stringency point, as estimated by Ashraf (2020), a large or a small effect relative to the 98% reduction in mortality predicted by the study published by Imperial College London (Ferguson et al. (2020))? This question is the subject for our meta-analysis in the next section. Here, it turns out that Ashraf’s (2020) −0.073 to −0.326 deaths/million per stringency point represents a relatively modest effect—one that corresponds to only a 2.4% reduction in COVID-19 mortality on average in the U.S. and Europe.

## 4 Meta-analysis: The impact of lockdowns on COVID-19 mortality

We now turn to the meta-analysis, where we focus on the impact of lockdowns on COVID-19 mortality.

In the meta-analysis, we include 22 studies in which we can derive a *standardized estimate,* which is the relative effect of lockdowns on COVID-19 mortality. That is, we obtain an estimate of how many percent fewer deaths there were due to lockdowns. For some studies, the authors state the relative effect, and our standardized estimate is thus readily available. For other studies, their estimate must be converted to our standardized estimate. Doing so is fairly straightforward for most studies and our calculations are explained in Table 19. However, the estimates from 10 studies cannot be converted to our standardized estimates. This include studies estimating the effect of lockdowns on mortality growth rates, unless the authors, like e.g. Chernozhukov et al. (2021), calculate a counterfactual scenario.^42^ Also, Mccafferty and Ashley (2021) estimate the effect of lockdowns on peak mortality, but an estimate of peak mortality cannot meaningfully be converted to our standardized estimate, which measures the relative impact on COVID-19 mortality.

The conclusions in the excluded studies are, overall, in correspondence with the conclusions in the included studies, see Table 3. Five excluded studies find lockdowns reduce mortality, two find mixed effects of NPIs with some NPIs working and others not working, and three find no or positive (more deaths) effects of NPIs. In comparison, eight of the included studies find lockdowns reduce mortality, 11 find mixed or insignificant effects of NPIs, and three studies find no or positive (more deaths) effects of lockdowns. Our reasons for not including a study are described in Table 19 in Appendix I.

**Table 3:** Conclusions from included and excluded studies in the meta-analysis are similar.

| Conclusion | Number (share) of studies included in meta-analysis | Number (share) of studies not included in meta-analysis |
| --- | --- | --- |
| Find that lockdowns reduce mortality | 8 (36%) | 5 (50%) |
| Find mixed effects of NPIs | 11 (50%) | 3 (30%) |
| Find that lockdowns have no effect (or increases mortality) | 3 (14%) | 2 (20%) |
| Total | 22 | 10 |

*Note: Mixed effects means that some of the examined NPIs reduce mortality while others increase mortality. The category “Find that lockdowns reduce mortality” includes studies which find at least one significant negative (fewer deaths) estimate ( $p < 0.05$ ) and no significant positive estimates (more deaths). The category “Find mixed effects of NPIs” includes studies which find both significantly negative and significantly positive estimates. The category “Find that lockdowns have no effect (or increases mortality)” includes studies which find no significant estimates or find at least one significant positive (more deaths) estimate ( $p < 0.05$ ) and no significant negative (fewer deaths) estimates.*

The studies we examine are placed in three categories. Eight studies analyze the effect of stricter lockdowns based on the OxCGRT stringency indices, 12 studies analyze the effect of SIPOs (six studies only analyze SIPOs and six analyze SIPOs together with other interventions), and eight studies analyze the effect of specific NPIs independently.^43^ Each of these categories is handled so that comparable estimates can be made across categories.

### Bias dimensions

Not all eligible studies are of the same quality. One way to handle this problem is to evaluate the quality of each study and use this evaluation to weigh or group the studies. However, there is currently no consensus as to best practices and/or an established scientific framework for evaluating the effectiveness of NPIs and lockdowns.^44^ As a result, such evaluation risk being subjective. Instead, we investigate whether there are any biases in the reviewed studies that can affect the studies’ conclusions. We do this by dividing them into different “bias dimensions”. Below, we describe the dimensions, as well as our reasons to believe why they could describe important biases. We also describe which group we perceive as “better” (meaning “probably less biased”). However, it should be noted that the primary objective of these dimensions is to identify and understand any biases in the studies, which can affect our overall results.

- *Peer-reviewed vs. working papers*: We distinguish between peer-reviewed studies and working papers. All else being equal, we perceive peer-reviewed studies as better than working papers.^45^
- *Long vs. short data periods*: We distinguish between studies based on long time periods (with data series ending *after* May 31, 2020) and short time periods (data series ending at or before May 31, 2020), because the first wave did not fully end before late June in the U.S. and Europe. Thus, studies relying on short data periods omit the last part of the first wave and may yield biased results if lockdowns only “flatten the curve” and do not prevent deaths. All else being equal, we perceive studies based on long periods as better than studies based on short periods.
- *No early effect vs. early effect on mortality*: On average, it takes approximately three to four weeks from infection to death.^46^ However, several studies find effects of lockdown on mortality almost immediately, which is clearly inconsistent with the standard view of COVID-19 transmission. Fowler et al. (2021) find a significant effect of SIPOs on mortality after just four days and the largest effect after 10 days. An early effect may indicate that other factors (omitted variables) drive the results. Thus, we distinguish between studies which find an effect on mortality sooner than 14 days after lockdown and those that do not.^47^ Note that many studies do not look at the short term and thus fall into the latter category by default. All else being equal, we perceive no early effect as better than an early effect (or no info).
- *Lagged vs. no lag of policy measures*: On average, it takes approximately three to four weeks from COVID-19 infection to death. Hence, the effect of a lockdown policy on mortality should be seen about three weeks after the policy measure is implemented, and a specification with a 2-4 weeks lagged policy variable is expected to be better than a specification with no lag, because the latter also captures the development around the policy decision which is not influenced by the policy.^48^ All else being equal, we perceive a lagged effect to be better than no lag.
- *Panel vs. no panel estimation:* The development of a pandemic in a country is affected by several factors and therefore may be inherently different from country to country. One way to handle these intrinsic differences is to exploit the panel data structure using fixed or random effects regression. Thus, we distinguish between studies which use fixed or random effects and those that do not. All else being equal, we perceive fixed/random effects regression as better than those that do not use fixed/random effects.
- *Verified vs. unverified data:* We distinguish between studies using verified data (e.g., from OxCGRT or New York Times) and unverified data (e.g., data collected by researchers but not readily publicly available and not updated on a continuous basis). All else being equal, we perceive studies using verified data as better than studies using unverified data.^49^
- *Address vs. do not address causality*: Not all studies address the causality/endogeneity question. We distinguish between studies addressing the causality question and studies which do not address the question. We consider the question as addressed if the authors handle causality technically (e.g., using instrument variable or lagged dependents) or argue for the causality of their results. All else being equal, we perceive studies that address causality as better than studies which do not address causality.
- *Social sciences vs. other sciences*: While it is true that epidemiologists and researchers in natural sciences should, in principle, know much more about COVID-19 and how it spreads than social scientists, social scientists are, in principle, experts in evaluating the effect of various policy interventions. Thus, we distinguish between studies published by scholars in social sciences and by scholars from other fields of research. For each study, we have registered the research field for the corresponding author’s associated institute (e.g., for a scholar from “Institute of Economics”, the research field is registered as “Economics”). Where no corresponding author was available, the affiliation of the first author has been used. Afterwards, all research fields have been classified as either from the “Social Science” or “Other.”^50^ All else being equal, we perceive social sciences as better suited for examining the effect of policies than other sciences.

We also considered including a bias dimension to distinguish between studies based on excess mortality and studies based on COVID-19 mortality, as we believe that excess mortality is potentially a better measure for two reasons. First, data on total deaths in a country is far more accurate than data on COVID-19 related deaths, which may be both underreported (due to lack of tests) or overreported (because some people die *with* – but not *because of* – COVID-19).

Second, a major goal with lockdowns was to save lives. To the extent lockdowns shift deaths *from* COVID-19 *to* other causes (e.g., suicide), estimates based on COVID-19 mortality will overestimate the effect of lockdowns. Likewise, if lockdowns save lives in other ways (e.g., fewer traffic accidents) lockdowns’ effect on mortality will be underestimated. However, as only one of the 32 studies, Bjørnskov (2021a),is based on excess mortality, we have to disregard this bias dimension.

Meta-data used for our bias dimensions as well as other relevant information are shown in Table 4.

**Table 4:** Bias dimension data for the studies included in the meta-analysis.

| 1. Study (Author & title) | 2. Publication status | 3. End of data period | 4. Earliest effect | 5. Policy/death lag | 6. Fixed/random effects | 7. Verified data | 8. Handling of causality | 9. Field of research | 10. Quality index |
| --- | --- | --- | --- | --- | --- | --- | --- | --- | --- |
| Alderman and Harjoto (2020); "COVID-19: US shelter-in-place orders and demographic characteristics linked to cases, mortality, and recovery rates" | Peer-review | 11-Jun-20 | n/a | No | No | Other official source (Verified) | Does not handle causality | Economics (Social science) | 0.25 |
| An et al. (2021); "Policy Design for COVID-19: Worldwide Evidence on the Efficacies of Early Mask Mandates and Other Policy Interventions" | Peer-review | 15-Jul-20 | n/a | Yes | Yes | Other official source (Verified) | Does not handle causality | Public policy (Social science) | 0.14 |
| Ashraf (2020); "Socioeconomic conditions, government interventions and health outcomes during COVID-19" | WP | 20-May-20 | n/a | No | Yes | OxCGRT stringency index (Verified) | Other model handling | Economics (Social science) | 0.25 |
| Berry et al. (2020); "Evaluating the effects of shelter-in-place policies during the COVID-19 pandemic" | Peer-review | 30-May-20 | 8-14 days | No | Yes | Other research data (Unverified) | Lag dependent (>2 weeks) | Public policy (Social science) | 0.00 |
| Bjørnskov (2021); "Did Lockdown Work? An Economist's Cross-Country Comparison" | Peer-review | 30-Jun-20 | <8 days | Yes | Yes | OxCGRT stringency index (Verified) | Instrument variable | Economics (Social science) | 0.77 |
| Bonardi et al. (2020); "Fast and local: How did lockdown policies affect the spread and severity of the covid-19" | WP | 13-Apr-20 | <8 days | Yes | Yes | Own data (Unverified) | Argumentation | Economics (Social science) | 0.39 |
| Chernozhukov et al. (2021); "Causal impact of masks, policies, behavior on early covid-19 pandemic in the U.S." | Peer-review | 03-Jun-20 | n/a | Yes | Yes | Other research data (Unverified) | Other model handling | Economics (Social science) | 0.14 |
| Chisadza et al. (2021); "Government Effectiveness and the COVID-19 Pandemic" | Peer-review | 01-Sep-20 | n/a | No | no | OxCGRT stringency index (Verified) | Does not handle causality | Economics (Social science) | 0.25 |
| Dave et al. (2020); "When Do Shelter-in-Place Orders Fight Covid-19 Best? Policy Heterogeneity Across States and Adoption Time" | Peer-review | 20-Apr-20 | Finds no effect | Yes | Yes | New York Times (Verified) | Argumentation | Economics (Social science) | 0.77 |
| Ertem et al. (2022); "The impact of school opening model on SARS-CoV-2 community incidence and mortality" | Peer-review | 21-Dec-20 | >21 days | Yes | Yes | Other data (Unverified) | Does not handle causality | Engineering (Other) | 0.77 |
| Fowler et al. (2021); "Stay-at-home orders associate with subsequent decreases in COVID-19 cases and fatalities in the United States" | Peer-review | 07-May-20 | <8 days | No | Yes | New York Times (Verified) | Other model handling | Public Health (Social science) | 0.39 |
| Fuller et al. (2021); "Mitigation Policies and COVID-19–Associated Mortality – 37 European Countries, January 23–June 30, 2020" | WP | 30-Jun-20 | n/a | Yes | No | OxCGRT stringency index (Verified) | Does not handle causality | Epidemiology (Other) | 0.14 |
| Gibson (2020); "Government mandated lockdowns do not reduce Covid-19 deaths: implications for evaluating the stringent New Zealand response" | Peer-review | 01-Jun-20 | Finds no effect | Yes | Yes | Other data (Unverified) | Instrument variable | Economics (Social science) | 0.77 |
| Goldstein et al. (2021); "Lockdown Fatigue: The Diminishing Effects of Quarantines on the Spread of COVID-19 " | WP | 31-Dec-20 | <8 days | Yes | Yes | OxCGRT stringency index (Verified) | Lag dependent (>2 weeks) | International Development (Social science) | 0.56 |
| Guo et al. (2021); "Mitigation Interventions in the United States: An Exploratory Investigation of Determinants and Impacts" | Peer-review | 07-Apr-20 | n/a | No | Yes | Own data (Unverified) | Does not handle causality | Social work (Social science) | 0.06 |
| Hale et al. (2021); "Government responses and COVID-19 deaths: Global evidence across multiple pandemic waves" | Peer-review | 11-Mar-21 | n/a | Yes | Yes | OxCGRT stringency index (Verified) | Other model handling | Government (Social science) | 0.77 |
| Leffler et al. (2020); "Association of country-wide coronavirus mortality with demographics, testing, lockdowns, and public wearing of masks" | Peer-review | 09-May-20 | n/a | No | No | OxCGRT data (Verified) | Does not handle causality | Ophthalmology (Other) | 0.39 |
| Sears et al. (2020); "Are we #stayinghome to Flatten the Curve?" | WP | 29-Apr-20 | Finds no effect | No | Yes | New York Times (Verified) | Argumentation | Economics (Social science) | 0.39 |
| Shiva and Molana (2021); "The Luxury of Lockdown" | Peer-review | 08-Jun-20 | 15-21 days | Yes | Yes | OxCGRT stringency index (Verified) | Does not handle causality | Government (Social science) | 0.77 |
| Spiegel and Tookes (2021); "Business restrictions and Covid-19 fatalities" | Peer-review | 31-Dec-20 | 15-21 days | Yes | No | Own data (Unverified) | Other model handling | Management (Social science) | 0.25 |
| Stokes et al. (2020); "The relative effects of non-pharmaceutical interventions on early Covid-19 mortality: natural experiment in 130 countries" | WP | 01-Jun-20 | n/a | Yes | Yes | OxCGRT data (Verified) | Other model handling | Economics (Social science) | 0.06 |
| Yang et al. (2021); "What is the relationship between government response and COVID-19 pandemics? Global evidence of 118 countries" | Peer-review | 03-Dec-20 | <8 days | Yes | No | OxCGRT stringency index (Verified) | Does not handle causality | Economics (Social science) | 0.39 |

*Note: Research fields classified as social sciences were economics, public health, health science, management, political science, government, international development, and public policy, while research fields not classified as social sciences were ophthalmology, environment, medicine, evolutionary biology and environment, human toxicology, epidemiology, and anesthesiology.*

### Interpreting and weighting estimates

The estimates used in the meta-analysis are not always readily available in the studies shown in Table 4. In Table 19 in Appendix I, we describe for each paper how we interpret the estimates and how they are converted to a standardized estimate (the relative effect of lockdowns on COVID-19 mortality), which is comparable across all studies.

Following Paldam (2015) and Stanley and Doucouliagos (2010), we also convert standard errors (SE)^51^ and use the precision of each estimate (defined as 1/SE) to calculate the precision-weighted average of all estimates. The precision-weighted average (PWA) is our primary indicator of the efficacy of lockdowns, but we also report arithmetic averages and medians in the meta-analysis.

### Sensitivity analyses

Given the relatively low number of studies in the meta-analysis, a study with an outlier estimate or outlier weight may influence our primary indicator of the efficacy of lockdowns. One way to deal with this uncertainty and illustrate the robustness of our estimates is to cap the estimates and weights at the end of the tails.

We therefore carry out four sensitivity analyses, where we replace the outlier (min/max) estimate/weight with the nearest estimate/weight and recalculate the PWA. For instance, the conclusion of Chisadza et al. (2021) is an outlier which finds that the average lockdown *increases* COVID-19 mortality. In one sensitivity analysis, we replace the estimate from Chisadza et al. (2021) with the nearest estimate from Bjørnskov (2021a) and recalculate the PWA. We report the result of our four sensitivity analyses as a span (from/to) at the bottom of each table.

### Quality-adjusted precision-weighted average

As a supplement to the PWA and the sensitivity analyses, we also calculate a quality-adjusted PWA based on our bias dimensions displayed in Table 4. The quality-adjusted PWA is calculated as the PWA weighted by a quality index, where the score on the quality index for each study is the squared number of bias dimensions, where the study is of “better” quality. Hence, each study can score between 0 and 64 on the index (because it includes eight bias dimensions). Finally, the index is normalized to 0-1 by dividing by 64.

In the following sections, we present the meta-analysis for each of the three groups of studies: stringency index-studies, SIPO-studies, and studies analyzing specific NPIs.

### 4.1 Stringency index studies

Eight eligible studies examine the link between lockdown stringency and COVID-19 mortality.^52^ The results from these studies, converted to standardized estimates, are presented in Table 5 below. All studies are based on the COVID-19 Government Response Tracker’s (OxCGRT) stringency index of Oxford University’s Blavatnik School of Government, Hale et al. (2020).

**Table 5:** Estimates of the effect on COVID-19 mortality of the average lockdown in Europe and in the United States from studies based on the OxCGRT stringency index.

| *Effect on COVID-19 mortality* | **Standardized estimate**<br>**(Estimated Averted Deaths /**<br>**Total Deaths)** | **Standard error** | **Weight (1/SE)** |
| --- | --- | --- | --- |
| Bjørnskov (2021) | -0.3% | 0.822% | 122 |
| Shiva and Molana (2021) | -4.0%** | 0.395% | 253 |
| Chisadza et al. (2021) | 11.7%** | 1.442% | 69 |
| Goldstein et al. (2021) | -7.5% | 1.964% | 51 |
| Fuller et al. (2021) | -35.3%** | 9.085% | 11 |
| Ashraf (2020) | -2.4%** | 0.391% | 256 |
| Yang et al. (2021) | -16.3%** | 4.523% | 22 |
| Hale et al. (2021) | -16.9%** | 2.812% | 36 |
| **Precision-weighted average (arithmetic average / median)** | **-3.2% (-8.9%/-5.8%)** |  |  |
| **Sensitivity analysis (quality-adjusted PWA)** | **-4.4% to -3.0% (-3.8%)** |  |  |

*Note: \*\* (\*) denote significance at $p < 0.01$ ( $p < 0.05$ ). The table shows the estimates for each study converted to a standardized estimate, i.e., the implied effect on COVID-19 mortality in Europe and United States. A negative number corresponds to fewer deaths, so -5% means 5% lower COVID-19 mortality. For details on how the estimates are converted to standardized estimates see Table 19 in Appendix I. The quality-adjusted PWA is calculated as the PWA weighted by a quality index, where the score on the quality index for each study is the number of bias dimensions squared (except “peer-reviewed” and “social sciences”), where the study is of “better” quality, see Table 4.*

The OxCGRT stringency index neither measures the expected effectiveness of the lockdowns nor the expected costs. Instead, it describes the stringency based on nine equally weighted parameters.^53^ Many countries followed similar patterns and almost all countries closed schools, while only a few countries issued SIPOs without closing businesses. Hence, it is reasonable to perceive the stringency index as continuous, although not necessarily linear. The index includes recommendations (e.g. “workplace closing” is 1 if the government recommends closing (or work from home), see Hale, Angrist, Goldszmidt, et al. (2021b), but the effect of including recommendations in the index is primarily to shift the index parallelly upward and should not alter the results relative to our focus on mandated NPIs.

It is important to note that the index is far from perfect. As pointed out by Book (2020), it is certainly possibly to identify errors and omissions in the index. However, the index is objective and unbiased and as such, useful for cross-sectional analysis with several observations, even if not suitable for comparing the overall strictness of lockdowns across two countries. In any case, there is no better available measure to adopt for cross-country comparisons.

Since the studies examined use different units of estimates, we have created standardized estimates for Europe and United States to make them comparable. The standardized estimates show the effect of the average lockdown in Europe and United States (with average stringencies of 76 and 74, respectively, between March 16^th^ and April 15^th^, 2020)^54^ compared to the most lenient lockdown, which we define as a COVID-19 policy based solely on recommendations (stringency 44).^55^

For example, Ashraf (2020) estimates that the effect of stricter lockdowns is −0.073 to −0.326 deaths/million per stringency point. We use the average of these two estimates (−0.200) in the meta-analysis. The average lockdown in Europe between March 16^th^ and April 15^th^, 2020, was 32 points stricter than a policy solely based on recommendations (76 vs. 44). In the United States, it was 30 points. Hence, the total effect of the lockdowns compared to the recommendation policy was (using rounded numbers) −6.37 deaths/million in Europe (32 × - 0.200) and −5.91 (30 × −0.200) deaths/million in United States. With populations of 748 million and 333 million, respectively the total effect as estimated by Ashraf (2020) is 4,766 averted COVID-19 deaths in Europe and 1,969 averted COVID-19 deaths in United States. By the end of the study period in Ashraf (2020), which is May 20, 2020, 164,600 people in Europe and 97,081 people in the United States had died of COVID-19. Hence, the 4,766 averted COVID-19 deaths in Europe and the 1,969 averted COVID-19 deaths in the United States corresponds to 2.8% and 2.0% of all COVID-19 deaths, respectively, with an arithmetic average of 2.4%.

Our standardized estimate is thus −2.4%, see Table 5. Our approach is not unproblematic. First of all, the level of stringency varies over time for all countries. Secondly, OxCGRT has changed the index over time, and a 10-point difference today may not be the exact same as a 10-point difference when the studies were finalized. However, we believe these problems are small and unlikely to significantly alter our results.

Table 5 demonstrates that the studies find that lockdowns, on average, have reduced COVID-19 mortality rates by 3.2% (precision-weighted average) and the sensitivity analysis shows a span from 4.4% to 3.0%. The results yield an arithmetic average of 8.9% and a median of 5.8%. To put the estimate in perspective, there were 188,542 registered COVID-19 deaths in Europe and 128,063 COVID-19 deaths in United States by June 30, 2020. Thus, the 3.2% PWA (8.9% arithmetic average, 5.8% median) corresponds to 6,000 (18,000, 12,000) avoided deaths in Europe and 4,000 (13,000, 8,000) avoided deaths in the United States.^56^ In comparison, there are approximately 72,000 flu deaths in Europe and 38,000 flu deaths in the United States each year.^57^

Hence, based on the stringency index studies, we find that mandated lockdowns in Europe and the United States had little to no effect on COVID-19 mortality rates.

We now turn to the bias dimensions. Table 6 presents the results differentiated by the bias dimensions. We find no evidence of significant biases. Although the effect is generally of a larger magnitude (more negative, i.e., fewer deaths) for studies we – all else being equal – perceive as better, the difference is marginal. Only one bias dimension, social sciences vs. other sciences, shows a large difference. This is because Fuller et al. (2021) find that lockdowns reduced COVID-19 mortality rates by 35.3%, and is the only study from the non-social sciences. Fuller et al. (2021) do not exploit the panel structure of their data by using fixed or random effects, nor do they address the causality question. There are three studies which both lag the policy implementation, exploit panel estimation, and address the causality question. These three studies find that lockdowns reduced COVID-19 mortality rates by 4.9% (compared to 2.6% for the other studies).

**Table 6:** Estimates of the effect on COVID-19 mortality of the average lockdown in Europe and in the United States from studies based on the OxCGRT stringency index classified according to the bias dimensions.

| *Values show effect on COVID-19 mortality* | Precision-weighted average | Arithmetic average | Median |
| --- | --- | --- | --- |
| **Peer-reviewed vs. working papers** |  |  |  |
| Peer-reviewed [5] | -2.4% | -5.2% | -4.0% |
| Working paper [3] | -5.0% | -15.1% | -7.5% |
| **Long vs. short data period** |  |  |  |
| Data period ends after 31 May 2020 [7] | -3.5% | -9.8% | -7.5% |
| Data period ends before 31 May 2020 [1] | -2.4% | -2.4% | -2.4% |
| **No early effect on mortality** |  |  |  |
| Does not find an effect within the first 14 days [1] | -4.0% | -4.0% | -4.0% |
| Finds effect within the first 14 days (including n/a) [7] | -2.8% | -9.6% | -7.5% |
| **Lag vs. no lag of policy measures** |  |  |  |
| Lag policy implementation [6] | -5.6% | -13.4% | -11.9% |
| No lag of policy implementation [2] | 1.5% | 4.7% | 4.7% |
| **Panel vs. no panel estimation** |  |  |  |
| Panel estimation [5] | -3.8% | -6.2% | -4.0% |
| No panel estimation [3] | 0.6% | -13.3% | -16.3% |
| **Verified vs. unverified data** |  |  |  |
| Verified data [8] | -3.2% | -8.9% | -5.8% |
| Unverified data [0] | n/a | n/a | n/a |
| **Address vs. do not address causality** |  |  |  |
| Address causality [4] | -3.7% | -6.8% | -5.0% |
| Do not address causality [4] | -2.7% | -11.0% | -10.2% |
| **Social sciences vs. other sciences** |  |  |  |
| Social sciences [7] | -2.8% | -5.1% | -4.0% |
| Other sciences [1] | -35.3% | -35.3% | -35.3% |

*Note: The table shows the standardized estimate as described in Table 5 for each bias dimension. The number of studies in each category is in square brackets. A negative number corresponds to fewer deaths, so -5% means 5% lower COVID-19 mortality.*

#### Overall conclusion on stringency index studies

Compared to a policy based solely on recommendations, we find little evidence that *stricter* lockdowns had a noticeable impact on COVID-19 mortality. Only one study, Fuller et al. (2021), finds a substantial effect of stricter lockdowns compared to the most lenient lockdown, while the remaining studies find little to no effect. Indeed, according to the stringency index studies, the average lockdown in Europe and the United States only reduced COVID-19 mortality by 3.2% using the precision-weighted average. The sensitivity analysis ranges from 4.4% and 3.0%, and overall, our bias dimensions do not suggest that biases are important.

We stress that this result does not imply that lockdowns do not work. It simply indicates that the most lenient lockdowns had virtually the same effect on mortality as stricter lockdowns. Since no country did nothing, we cannot reject the thesis that some NPI would be required, e.g., to spur voluntary behavioral changes.^58^

It should also be noted that the eight stringency studies are all based on the same index (OxCGRT stringency index). Although OxCGRT is widely recognized as the best index recording the strictness of ‘lockdown style’ policies that restrict people’s behavior and tracks and compares policy responses around the world, rigorously and consistently, we cannot rule out the possibility that the lack of evidence of the efficacy of lockdowns is caused by the limitations of the index.^59^ In the following section, we will look at the effect of one of the strictest NPIs used during the COVID-19 pandemic, SIPOs, following the same structure as the current section.

### 4.2 Shelter-in-place order (SIPO) studies

We have identified 12 eligible studies that estimate the effect of Shelter-In-Place Orders (SIPOs) on COVID-19 mortality, see Table 7.^60^ Six of these studies look at multiple NPIs of which a SIPO is just one, while six studies estimate the effect of a SIPO vs. no SIPO in the United States. According to the containment and closure policy indicators from OxCGRT, 41 states in the United States issued SIPOs in the spring of 2020. Usually, these were introduced after implementing other NPIs such as school closures or workplace closures.

**Table 7:** Estimates of the effect on COVID-19 mortality of shelter-in-place orders (SIPO)

| *Values show effect on COVID-19 mortality* | Standardized estimate<br>(Estimated Averted Deaths /<br>Total Deaths) | Standard<br>error | Weight (1/SE) |
| --- | --- | --- | --- |
| **Studies where SIPO is one of several examined interventions and not (as) likely to capture the effect of other interventions** |  |  |  |
| Chernozhukov et al. (2021) | -17.7% | 14.3% | 7 |
| Stokes et al. (2020) | 4.9% | 2.8% | 36 |
| Spiegel and Tookes (2021b) | 13.1%* | 6.6% | 15 |
| Bonardi et al. (2020)* | 0.0% | n/a | n/a |
| Guo et al. (2021) | 4.6% | 14.8% | 7 |
| An et al. (2021) | 15.6% | 7.8% | 13 |
| Precision-weighted average (arithmetic average / median) where<br>SIPO is one of several variables | 6.0% (3.4%/4.7%) |  |  |
| **Studies where SIPO is the only examined intervention and may capture the effect of other interventions** |  |  |  |
| Sears et al. (2020) | -32.2% | 17.6% | 6 |
| Alderman and Harjoto (2020) | -1.0% | 0.6% | 169 |
| Berry et al. (2020)* | 1.1% | n/a | n/a |
| Fowler et al. (2021) | -35.0%** | 7.0% | 14 |
| Gibson (2020) | -6.0% | 24.3% | 4 |
| Dave et al. (2020) | -40.8% | 36.1% | 3 |
| Precision-weighted average (arithmetic average / median) where<br>SIPO is the only variable | -5.1% (-19.0%/-19.1%) |  |  |
| **Precision-weighted average (arithmetic average / median) for all**<br>**studies** | **-2.0% (-7.8%/-0.5%)** |  |  |
| **Sensitivity analysis (quality-adjusted PWA)** | **-4.1% to -1.4% (-1.8%)** |  |  |

Note: \*\* (\*) denote significance at $p < 0.01$ ( $p < 0.05$ ). A negative number corresponds to fewer deaths, so -5% means 5% lower COVID-19 mortality. The quality-adjusted PWA is calculated as the PWA weighted by a quality index, where the score on the quality index for each study is the number of bias dimensions squared (except “peer-reviewed” and “social sciences”), where the study is of “better” quality, see Table 4.
\* Bonardi et al. (2020) and Berry et al. (2021) do not affect the precision-weighted average, as we do not know the standard error.
\*\* An et al. (2021) report estimates for SIPO introduced both early and late.<sup>62</sup> For simplicity we only report the average estimate and standard error (detailed estimates are +13% and +18%). Stokes et al. (2020) report estimates for SIPO implemented before first death and within 14 days after first death. Again, for simplicity we only report the average estimate and standard error (detailed estimates are +1.8% and +8.0%%, average SE is calculated as $\sqrt{(SE_1^2 + SE_2^2 + \dots + SE_n^2)/k}$ , where $k$ is the number of estimates).
<sup>61</sup> The average estimated flu deaths in United States the five years prior to COVID-19 was 38,400 according to CDC (2022), and WHO (2022) writes that there are 72,000 flu deaths in Europe each year.
<sup>62</sup> An et al. (2021) define early mandate adoption as being taken within 14 days (the median in their dataset) after the first reported infection in each country.

On average, SIPOs were issued 7½ days after *both* schools and workplaces closed, and 12 days after the first of the two closed. Only one state, Tennessee, issued a SIPO before schools and workplaces closed. The 10 states that did not issue SIPOs all closed schools. Moreover, of those 10 states, three closed some non-essential businesses, while the remaining 7 closed all non-essential businesses. Because of this, we perceive estimates for SIPOs based on US-data as the marginal effect of SIPOs on top of other restrictions, although we cannot rule out that the estimates may capture the effects of other NPI measures as well.

The results of eligible studies based on SIPOs are presented in Table 7. This table demonstrates that the studies generally find that SIPOs have reduced COVID-19 mortality by 2.0% (precision-weighted average) and the sensitivity analysis shows a span from 4.1% to 1.4%. The arithmetic average estimate is 7.8% and the median is 0.5%. To put these numbers into perspective, there were 188,542 registered COVID-19 deaths in Europe and 128,063 COVID-19 deaths in the United States by June 30, 2020. Thus, the reduction of 2.0% PWA (7.8% arithmetic average, 0.5% median) corresponds to approximately 4,000 (16,000, 1,000) avoided deaths in Europe and 3,000 (11,000, 1,000) avoided deaths in the United States had all countries and states implemented SIPOs. In comparison, there are approximately 72,000 flu deaths in Europe and 38,000 flu deaths in the United States each year.^61^

There is an apparent difference between studies in which a SIPO is one of multiple NPIs, and studies in which a SIPO is the only examined intervention. The former group generally finds that SIPOs *increase* COVID-19 mortality by 6.0%, whereas the latter finds that SIPOs *decrease* COVID-19 mortality by 5.1%. As we will see below, this difference may – at least partly – be explained by the data period covered by each study.

Table 8 presents the results differentiated by bias dimensions. One bias dimension, long vs. short data period, shows a large potential bias driven by relatively short data periods. The four studies with relatively short data periods find a very large effect of SIPOs (25.9% reduction in mortality rates), while studies based on longer data periods find a modest increase in mortality rates of 1.0%. The last data points for the three studies which find the – by far – largest effects of SIPOs (Sears et al. (2020), Fowler et al. (2021), and Dave et al. (2021)) are April 29, May 7, and April 20 in 2020, respectively. These findings may indicate that SIPOs can delay deaths but not eliminate them completely. However, these studies were done very early in the pandemic and could not – as do the other studies – “stand on the shoulder of giants”. The bias dimensions “Lag vs. no lag of policy implementation”, “Panel vs. no panel estimation”, and “Address vs. do not address causality” also find some potential bias, although of less magnitude.

**Table 8:** Estimates of the effect on COVID-19 mortality of shelter-in-place orders (SIPO) classified according to the bias dimensions.

| *Values show effect on COVID-19 mortality* | Precision-weighted average | Arithmetic average | Median |
| --- | --- | --- | --- |
| **Peer-reviewed vs. working papers** |  |  |  |
| Peer-review [8] | -2.3% | -8.4% | -3.5% |
| Working paper [2] | -0.2% | -13.6% | 0.0% |
| **Long vs. short data period** |  |  |  |
| Data period ends after 31 May 2020 [6] | 1.0% | 1.5% | 1.9% |
| Data period ends before 31 May 2020 [4] | -25.9% | -25.8% | 0.0% |
| **No early effect on mortality** |  |  |  |
| Finds effect within the first 14 days [4] | -4.3% | -16.5% | -19.1% |
| Does not find an effect within the first 14 days (including n/a) [6] | -1.7% | -4.8% | 0.0% |
| **Lag vs. no lag of policy measures** |  |  |  |
| Lag policy implementation [6] | 3.8% | -5.2% | -0.6% |
| No lag of policy implementation [4] | -4.2% | -15.9% | 0.0% |
| **Panel vs. no panel estimation** |  |  |  |
| Panel estimation [8] | -6.4% | -13.3% | -11.9% |
| No panel estimation [2] | 0.2% | 6.0% | 0.0% |
| **Verified vs. unverified data** |  |  |  |
| Verified data [6] | -2.6% | -14.7% | -16.6% |
| Unverified data [4] | 2.5% | -1.5% | 0.0% |
| **Address vs. do not address causality** |  |  |  |
| Address causality [7] | -6.7% | -16.2% | -17.7% |
| Do not address causality [3] | 0.2% | 6.4% | 0.6% |
| **Social sciences vs. other sciences** |  |  |  |
| Social sciences [10] | -2.0% | -9.4% | -3.5% |
| Other sciences [0] | n/a | n/a | n/a |

*Note: The table shows the standardized estimate as described in Table 7 for each bias dimension. The number of studies in each category is in square brackets (note that the numbers do not include Bonardi et al. (2020) and Berry et al. (2021), because they do not affect the precision-weighted average, as we do not know the standard error). A negative number corresponds to fewer deaths, so -5% means 5% lower COVID-19 mortality.*

There are three studies that both use long data series, lag the policy implementation, exploit panel estimation, and address the causality question. These studies find that SIPOs *increased* COVID-19 mortality rates by 0.6% (compared to an average reduction in mortality by 2.5% in the other studies).

#### Overall conclusion on SIPO studies

We find that SIPOs had little to no effect on COVID-19 mortality. On average, countries in Europe and states in the United States which used SIPOs only reduced COVID-19 mortality by 2.0% (precision-weighted average). The sensitivity analysis ranges from 1.4% to 4.1%, and our bias dimensions suggest that using long data series is important and that this will further reduce the estimated effect of SIPOs, possibly making the effect of a SIPO on COVID-19 mortality positive (more deaths).

Multiple studies find a small *positive* effect of SIPOs on COVID-19 mortality. Although such a result might appear to be counterintuitive, it could, for, example be the result of an (asymptomatic) infected person being isolated at home under a SIPO who can infect family members with a higher viral load causing more severe illness.^63,64^ Our result is in line with Nuzzo et al. (2019),who state that “in the context of a high-impact respiratory pathogen, quarantine may be the least likely NPI to be effective in controlling the spread due to high transmissibility” and World Health Organization Writing Group (2006),which concludes that “forced isolation and quarantine are ineffective and impractical.”^65^

In the following section, we will look at the effects found in studies analyzing other specific NPIs.

### 4.3 Studies of other specific NPIs

A total of eight eligible studies examine the effect of specific NPIs.^66^ The definition of these specific NPIs varies from study to study which makes comparison difficult. The variety of definitions can be seen in the analysis of non-essential business closures and bar/restaurant closures. Chernozhukov et al. (2021) focus on a combined parameter (the average of business closure and bar/restaurant closure in each state), Spiegel and Tookes (2021b) examine bar and/or restaurant closure but not business closure, and Guo et al. (2021) look at both business closures and bar/restaurant closures independently.

Some studies include several NPIs (e.g. Stokes et al. (2020) and Spiegel and Tookes (2021b)), while others cover very few. For example, Leffler et al. (2020) look at internal lockdowns of any type, mask recommendations, and international travel restrictions). Too few NPIs in a model are potentially a problem because they can capture the effect of excluded NPIs.^67^ On the other hand, several NPIs in a model increase the risk of multiple test bias. Also, looking at one NPI at a time may be problematic, as behavioral spillover effects may not be fully captured. For example, if we show that closing bars work because people who went to bars were more likely to be infected than people not going to bars, then this finding does not automatically imply that closing bars will have a significant impact on the overall number of infections, if people adjust their behavior according to official case numbers and are more careful when case numbers rise.

The differences in the choice of NPIs and in the number of NPIs generally make it challenging to create an overview of the results. In the following, we go through the evidence for the effectiveness of specific NPIs. First, we cover business closures, then school closures, limiting gatherings, border closures and face masks, as these NPIs are all covered by at least four studies. Last, we cover NPIs covered by one or two studies (cancellation of public events, closing public transport, and restrictions on internal movement).

#### Business closures

Five studies examine the effect of business closure on COVID-19 mortality.^68^ Table 9 presents an overview of the estimates in these studies. Closing businesses reduced COVID-19 mortality rates by 7.5% (precision-weighted average), and an arithmetic average of 10.5% and a median of 5.5%. The sensitivity analysis shows a span from 6.6% to 9.3% reduction in mortality rates.

**Table 9:** Estimates of the effect on COVID-19 mortality of business closures.

| Effect on COVID-19 mortality | Description | Standardized estimate<br>(Estimated Averted Deaths /<br>Total Deaths) | Standard<br>error | Weight<br>(1/SE) |
| --- | --- | --- | --- | --- |
| Chernozhukov et al. (2021)* | Measure is average of business and bar/restaurant closure | -28.6% | 20.842% | 5 |
| Spiegel and Tookes (2021b) | Business closure (average) | -13.3%* | 5.812% | 17 |
|  | Bars & restaurants | -50.2%** | 8.735% | 11 |
|  | Bars but not restaurants | -4.9% | 4.208% | 24 |
|  | Gyms closed | -13.8%** | 4.272% | 23 |
|  | Spas closed | 15.9%** | 4.782% | 21 |
| Stokes et al. (2020)* | Workplace closing | -4.9%** | 1.822% | 55 |
|  | Implemented before 1st death | -4.8%* | 1.955% | 51 |
|  | Implemented 14 days after 1st death | -5.0%** | 1.678% | 60 |
| Guo et al. (2021) | Business closure (average) | -0.4% | 15.616% | 6 |
|  | Business closure | 4.7% | 13.600% | 7 |
|  | Bars & restaurants | -5.5% | 17.400% | 6 |
| An et al. (2021) | Business closure (average) | -5.5% | 13.062% | 8 |
|  | Early business closure | -7.5% | 5.760% | 17 |
|  | Late business closure | -3.6% | 17.551% | 6 |
| Precision-weighted average (arithmetic average / median) |  | -7.5% (-10.5%/-5.5%) |  |  |
| Sensitivity analysis (quality-adjusted PWA) |  | -9.3% to -6.6% (-11.9%) |  |  |

Note: \*\* (\*) denote significance at $p < 0.01$ ( $p < 0.05$ ). For studies with several estimates related to the measure, we report all submeasures in grey but focus on the average of these measures and use the average when calculating the PWA. The average standard error is calculated as $\sqrt{(SE_1^2 + SE_2^2 + \dots + SE_n^2)/k}$ , where $k$ is the number of estimates. The quality-adjusted PWA is calculated as the PWA weighted by a quality index, where the score on the quality index for each study is the number of bias dimensions squared (except “peer-reviewed” and “social sciences”), where the study is of “better” quality, see Table 4. Values in grey are not included in the calculations of the precision-weighted average, the arithmetic average, and the median. A negative number corresponds to fewer deaths, so -5% means 5% lower COVID-19 mortality.
\* The estimate from Chernozhukov et al. (2021) is based on their model without the national deaths-variable (see their Table 7). They also run a model with national deaths, where they find that business closures increase mortality slightly (see their Table 9). However, since they do not calculate a counterfactual for this model, this estimate is not included in the Table 9.
\*\* Stokes et al. (2020) consider two distinct time periods of 24 days to account for the changing magnitude of effects of an exponentially transmitted virus: i) 0-24 days and ii) 14-38 after the first COVID-19 death. We report both of them and use the average in the precision-weighted average.

Three studies find little to no effect, one study (Spiegel and Tookes (2021b)) finds some effect, and another study (Chernozhukov et al. (2021)) finds a relatively large effect. It should be noted that the estimate from Chernozhukov et al. (2021) is based on their model without the national death-variable (see their Table 7). They also run a model with national deaths, where they find that business closures *increase* mortality (see their Table 9). However, since they do not calculate a counterfactual for this model, this estimate is not included in Table 9. If there is a large effect, it seems related to closing bars and restaurants. Indeed, the “close business” category in Chernozhukov et al. (2021) is an average of closed businesses, restaurants, and movie theaters. And, the “closing bars and restaurants”-submeasure (in grey in Table 9) in Spiegel and Tookes (2021b) delivers largest relative effect. Note that overall estimate of business closures for Spiegel and Tookes (2021b) is much smaller than the estimate for just its “closing bars and restaurants”-submeasure.

#### School closures

Four studies examine the effect of school closure on COVID-19 mortality.^69^ Table 10 presents an overview of the estimates in the studies. Closing schools reduced COVID-19 mortality rates by 5.9% (precision-weighted average) with an arithmetic average of 0.2% and a median of 0.0%. The sensitivity analysis shows a span from 2.5% to 6.2%.

**Table 10:** Estimates of the effect on COVID-19 mortality of school closures.

| Effect on COVID-19 mortality | Description | Standardized estimate<br>(Estimated Averted Deaths /<br>Total Deaths) | Standard error | Weight<br>(1/SE) |
| --- | --- | --- | --- | --- |
| Stokes et al. (2020)* | School closures (average) | -10.9%** | 1.567% | 64 |
|  | *School closing implemented before 1<sup>st</sup> death* | -2.2% | 1.681% | 59 |
|  | *School closing implemented after 1<sup>st</sup> death* | -19.7%** | 1.443% | 69 |
| Guo et al. (2021)** | School closures (average) | 10.1% | 20.662% | 5 |
|  | *Public school closure* | -0.2% | 23.400% | 4 |
|  | *All school closure* | 20.4% | 17.500% | 6 |
| An et al. (2021) | School closures (average) | 3.8% | 7.374% | 14 |
|  | *Early school closure* | 2.8% | 7.590% | 13 |
|  | *Late school closure* | 4.7% | 7.152% | 14 |
| Ertem et al. (2022)*** | Opening schools (virtual vs. traditional) | -3.7%** | 1.982% | 50 |
| Precision-weighted average (arithmetic average / median) |  | -5.9% (-0.2%/0.0%) |  |  |
| Sensitivity analysis (quality-adjusted PWA) |  | -6.2% to -2.5% (-1.2%) |  |  |

Note: \*\* (\*) denote significance at $p < 0.01$ ( $p < 0.05$ ). For studies with several estimates related to the measure, we report all submeasures in grey but focus on the average of these measures and use the average when calculating the PWA. The average standard error is calculated as $\sqrt{(SE_1^2 + SE_2^2 + \dots + SE_n^2) / k}$ , where $k$ is the number of estimates. Chernozhukov et al. (2021) also examine the effect of school closures but does not report a counterfactual estimate. Based on a back-of-the envelope sensitivity analysis it is unlikely that including Chernozhukov et al. (2021) would have a considerable effect on the result. The quality-adjusted PWA is calculated as the PWA weighted by a quality index, where the score on the quality index for each study is the number of bias dimensions squared (except “peer-reviewed” and “social sciences”), where the study is of “better” quality, see Table 4. Values in grey are not included in the calculations of the precision-weighted average, the arithmetic average, and the median. A negative number corresponds to fewer deaths, so -5% means 5% lower COVID-19 mortality.
\* Stokes et al. (2020) consider two distinct time periods of 24 days to account for the changing magnitude of effects of an exponentially transmitted virus: i) 0-24 days and ii) 14-38 after the first COVID-19 death. We report both of them and use the average in the precision-weighted average.
\*\* The estimate from Guo et al. (2021) is based on school closures in United States where all 50 states closed schools between March 13, 2020, and March 23, 2020, and 44 states closed schools in just four school days (March 15, 2020 (Sunday) to March 19, 2020 (Friday)), see Table 1 in Auger et al. (2020). Hence, the difference-in-difference effect is based on a maximum of seven school days. As noted by Chernozhukov et al. (2021), this lack of cross-sectional variation can lead to considerable uncertainty over the effects of school closures, and the results from Guo et al. (2021) should be treated with this in mind.
\*\*\* The standard error for Ertem et al. (2021) is underestimated and, thus, the weight, $1/SE$ , is overestimated. See the description for Ertem et al. (2021) in Table 19 in Appendix I for further details.

Since schools in United States were closed almost simultaneously, the estimate from Guo et al. (2021) suffers from lack of variation in school closures, but has little impact on the precision-weighted average due to the low precision/weight.^70^ Ertem et al. (2021) look at school re-openings and find a (small) increase in mortality rates following school reopenings. Assuming that the effect of closing and reopening is identical, this corresponds to a (small) decrease in mortality rates following school closures.

The absence of a notable effect of school closures is in line with Irfan et al. (2021), who – based on a systematic review and meta-analysis of 90 published or preprint studies of transmission in children – concluded that “risks of infection among children in educational-settings was lower than in communities. Evidence from school-based studies demonstrate it is largely safe for young children (<10 years of age) to be at schools; however, older children (between 10 and 19 years of age) might facilitate transmission.” UNICEF (2020) and ECDC (2020) reach similar conclusions. UNICEF (2020) concludes, “The preliminary findings thus far suggest that in-person schooling – especially when coupled with preventive and control measures – had lower secondary COVID-19 transmission rates compared to other settings and do not seem to have significantly contributed to the overall community transmission risks.” Whereas, ECDC (2020) concludes, “School closures can contribute to a reduction in SARS-CoV-2 transmission, but by themselves are insufficient to prevent community transmission of COVID-19 in the absence of other nonpharmaceutical interventions (NPIs) such as restrictions on mass gathering,” and concludes “There is a general consensus that the decision to close schools to control the COVID-19 pandemic should be used as a last resort. The negative physical, mental health and educational impact of proactive school closures on children, as well as the economic impact on society more broadly, would likely outweigh the benefits.” ^71^

Even though UNICEF (2020) and ECDC (2020) published their reviews in December 2020, there were still at least 160 countries who closed schools during 2021, according to Oxford COVID-19 Government Response Tracker.^72^

#### Limiting gatherings

Four studies examine the effect of limiting gatherings. Table 11 presents an overview of the estimates in the studies. Limiting gatherings *increased* COVID-19 mortality rates by 5.9% (precision-weighted average) and the sensitivity analysis shows a span from a 4.9% to an 8.9% *increase* in mortality rates. The arithmetic average is 8.5% and the median is 7.0%, while the quality-adjusted PWA is 9.8%.

**Table 11:** Estimates of the effect on COVID-19 mortality of limiting gatherings.

| Effect on COVID-19 mortality | Description | Standardized estimate<br>(Estimated Averted<br>Deaths /<br>Total Deaths) | Standard<br>error | Weight<br>(1/SE) |
| --- | --- | --- | --- | --- |
| Stokes et al. (2020) | Limiting gatherings (average) | 3.8%** | 1.307% | 77 |
|  | *Restrictions implemented before 1<sup>st</sup> death* | 2.4% | 1.403% | 71 |
|  | *Restrictions implemented after 1<sup>st</sup> death* | 5.2%** | 1.204% | 83 |
| Spiegel and Tookes (2021b) | Limiting gatherings (average) | 16.0%* | 7.014% | 14 |
|  | *Gatherings <=10* | 4.5% | 8.034% | 12 |
|  | *Gatherings 11-100* | 19.1%** | 6.249% | 16 |
|  | *Gatherings (limit > 100)* | 24.4%** | 6.631% | 15 |
| Guo et al. (2021) | Limiting gatherings (average) | 5.7% | 13.100% | 8 |
|  | *Gatherings <=10* | 8.7% | 13.050% | 8 |
|  | *Gatherings 11-100* | 2.7% | 13.150% | 8 |
| An et al. (2021) | Limiting gatherings (average) | 8.4% | 12.442% | 8 |
|  | *Early gathering limits* | 1.1% | 8.149% | 12 |
|  | *Late gathering limits* | 15.7% | 15.595% | 6 |
| Precision-weighted average (arithmetic average / median) |  | 5.9% (8.5%/7.0%) |  |  |
| Sensitivity analysis (quality-adjusted PWA) |  | 4.9% to 8.9% (9.8%) |  |  |

Note: \*\* (\*) denote significance at $p < 0.01$ ( $p < 0.05$ ). For studies with several estimates related to the measure, we report all submeasures in grey but focus on the average of these measures and use the average when calculating the PWA. The average standard error is calculated as $\sqrt{(SE_1^2 + SE_2^2 + \dots + SE_n^2)/k}$ , where $k$ is the number of estimates. A negative number corresponds to fewer deaths, so -5% means 5% lower COVID-19 mortality. The quality-adjusted PWA is calculated as the PWA weighted by a quality index, where the score on the quality index for each study is the number of bias dimensions squared (except “peer-reviewed” and “social sciences”), where the study is of “better” quality, see Table 4. Values in grey are not included in the calculations of the precision-weighted average, the arithmetic average, and the median. Stokes et al. (2020) consider two distinct time periods of 24 days to account for the changing magnitude of an exponentially transmitted virus: i) 0-24 days and ii) 14-38 after the first COVID-19 death. We report both of them and use the average in the precision-weighted average.

It is worth noting that no studies have estimates showing that limiting gatherings reduced COVID-19 mortality. Indeed, all four studies find positive – and sometimes rather large – effects of limiting gatherings on mortality.

#### Travel restrictions

Five studies examine the effect of travel restrictions.^73^ Table 12 presents an overview of the estimates in these studies. Travel restrictions reduced COVID-19 mortality rates by 3.4% (precision-weighted average) and the sensitivity analysis shows a span from 0.4% to 4.7%. The arithmetic average is an 5.3% *increase* in mortality and the median is 0.0%, while the quality-adjusted PWA shows an *increase* of 2.1%. Note that the description of the NPI varies greatly between studies and may not be comparable. This may partly explain the large span of estimates (from a *reduction* of 15.6% to an *increase* of 36.3%).

#### Mask mandates

The three studies examining the effect of mask mandates – an intervention that was not widely used in the spring of 2020, and in many countries was even discouraged – on average find that masks mandates reduced COVID-19 mortality by 18.7% (precision-weighted average), see Table 13. The sensitivity analysis shows a span from 12.5% to 19.9%, and the arithmetic average and median are 18.7% and 13.5%, respectively.

**Table 12:** Estimates of the effect on COVID-19 mortality of travel restrictions.

| Effect on COVID-19 mortality | Description | Standardized estimate<br>(Estimated Averted Deaths / Total Deaths) | Standard error | Weight (1/SE) |
| --- | --- | --- | --- | --- |
| Leffler et al. (2020) | International travel restrictions | -15.6% | 5.737% | 17 |
| Stokes et al. (2020) | Travel restrictions (average) | -6.1%** | 1.320% | 76 |
|  | Travel restrictions implemented before 1<sup>st</sup> death | -2.1% | 1.417% | 71 |
|  | Travel restrictions implemented after 1<sup>st</sup> death | -10.0%** | 1.216% | 82 |
| Bonardi et al. (2020) | Border closures | 0.0% | n/a | n/a |
| Guo et al. (2021) | Quarantine of travelers | 36.3% | 16.950% | 6 |
| An et al. (2021) | Travel restrictions (average) | 12.1% | 8.440% | 12 |
|  | Early travel restrictions | 5.5% | 5.366% | 19 |
|  | Late travel restrictions | 18.7% | 10.662% | 9 |
| Precision-weighted average (arithmetic average / median) |  | -3.4% (5.3%/0.0%) |  |  |
| Sensitivity analysis (quality-adjusted PWA) |  | -4.7% to -0.4% (1.1%) |  |  |

Note: \*\* (\*) denote significance at $p < 0.01$ ( $p < 0.05$ ). For studies with several estimates related to the measure, we report all submeasures in grey but focus on the average of these measures and use the average when calculating the PWA. The average standard error is calculated as $\sqrt{(SE_1^2 + SE_2^2 + \dots + SE_n^2)/k}$ , where $k$ is the number of estimates. A negative number corresponds to fewer deaths, so -5% means 5% lower COVID-19 mortality. The quality-adjusted PWA is calculated as the PWA weighted by a quality index, where the score on the quality index for each study is the number of bias dimensions squared (except “peer-reviewed” and “social sciences”), where the study is of “better” quality, see Table 4. Values in grey are not included in the calculations of the precision-weighted average, the arithmetic average, and the median. Stokes et al. (2020) consider two distinct time periods of 24 days to account for the changing magnitude of effects of an exponentially transmitted virus: i) 0-24 days and ii) 14-38 after the first COVID-19 death. We report both of them and use the average in the precision-weighted average.

**Table 13:** Estimates of the effect on COVID-19 mortality of mask mandates.

| Effect on COVID-19 mortality | Description | Standardized estimate<br>(Estimated Averted Deaths /<br>Total Deaths) | Standard<br>error | Weight<br>(1/SE) |
| --- | --- | --- | --- | --- |
| Chernozhukov et al. (2021) | Employee face masks | -34.0% | 8.511% | 12 |
| Spiegel and Tookes (2021b) | Average of below measures | -13.5% | 5.131% | 19 |
|  | Employee face masks | -12.2% | 4.272% | 23 |
|  | Mandatory face masks | -14.9% | 5.866% | 17 |
| An et al. (2021) | Mask mandates | -8.5% | 12.697% | 8 |
|  | Early mandates | -8.0% | 17.246% | 5.8 |
|  | Late mandates | -9.1% | 5.000% | 20 |
| Precision-weighted average (arithmetic average / median) |  | -18.7% (-18.7%/-13.5%) |  |  |
| Sensitivity analysis (quality-adjusted PWA) |  | -19.9% to -12.5% (-18.2%) |  |  |

*Note: \*\* (\*) denote significance at $p < 0.01$ ( $p < 0.05$ ). For studies with several estimates related to the measure, we report all submeasures in grey but focus on the average of these measures and use the average when calculating the PWA. The average standard error is calculated as $\sqrt{(SE_1^2 + SE_2^2 + \dots + SE_n^2)/k}$ , where $k$ is the number of estimates. A negative number corresponds to fewer deaths, so -5% means 5% lower COVID-19 mortality. The quality-adjusted PWA is calculated as the PWA weighted by a quality index, where the score on the quality index for each study is the number of bias dimensions squared (except “peer-reviewed” and “social sciences”), where the study is of “better” quality, see Table 4. Values in grey are not included in the calculations of the precision-weighted average, the arithmetic average, and the median.*

The description of the NPI varies greatly between studies and may not be comparable. Chernozhukov et al. (2021) find that “employee face masks” reduces mortality by 34%, and, thus, do not – like An et al. (2021) – look at a general mask mandate. Spiegel and Tookes (2021b) look at both “employee face masks” and “mandatory face masks,” finding similar effects. We do not include the estimate on “mask recommendation” from Leffler et al. (2020), as it is not a mandated NPI, but interestingly Leffler et al. (2020) find that mask recommendations reduced mortality by 23%, which is in the same order of magnitude we find for mask mandates.

Our findings are in contrast to several other reports and studies. WHO (2019) concludes that “ten RCTs were included in meta-analysis, and there was no evidence that face masks are effective in reducing transmission of laboratory-confirmed influenza” and in their Influenza Pandemic Preparedness Strategy UK Department of Health, Social Services, and Public Safety (2011) states that “in line with the scientific evidence, the Government will not stockpile facemasks for general use in the community”. Liu et al. (2021) conclude in a review that “fourteen of sixteen identified randomized controlled trials comparing face masks to no mask controls failed to find statistically significant benefit in the intent-to-treat populations.” Similarly, a pre-COVID Cochrane review, Jefferson et al. (2020), concludes, “There is low certainty evidence from nine trials (3,507 participants) that wearing a mask may make little or no difference to the outcome of influenza-like illness (ILI) compared to not wearing a mask (risk ratio (RR) 0.99, 95% confidence interval (CI) 0.82 to 1.18). There is moderate certainty evidence that wearing a mask probably makes little or no difference to the outcome of laboratory-confirmed influenza compared to not wearing a mask (RR 0.91, 95% CI 0.66 to 1.26; 6 trials; 3,005 participants)”.^74^

However, it should be noted that even if no effect is found in controlled settings, this does not necessarily imply that mask mandates do not reduce mortality, as other factors may play a role (e.g. wearing a mask may function as a tax on socializing if people are bothered by wearing a face masks when they socialize, or masks may function as a constant reminder of the presence of the pandemic). In a cluster-randomized trial of community-level mask promotion in Bangladesh, Abaluck et al. (2022) found that the intervention (which included free masks, information on the importance of masking, role modeling by community leaders, and in-person reminders for 8 weeks) reduced symptomatic seroprevalence by 9.3%. Another possible explanation is that masks reduce the viral inoculum and that this affects mortality. For example, Bielecki et al. (2021) – in a sort of natural experiment – find that in two groups of Swiss soldiers, soldiers in the one group – those who were *not* physically distancing or wearing surgical masks before being exposed to COVID-19 – had more symptoms (47%) than the other group who *were* physically distancing and wearing surgical masks before being exposed to COVID-19.

#### Other NPIs (cancelling public events, closing public transportation, restrictions on internal movement, “lockdown vs. no lockdown”)

Table 14 presents the estimates for NPIs that are only covered by one study, as well as Leffler et al. (2020)’s estimate on lockdown vs. no lockdown. The estimates are all close-to-zero, but – needless to say – the uncertainty is large. We do note, however, that Leffler et al. (2020) support the other results in the meta-analysis, suggesting the effect of lockdowns of any type is limited.

**Table 14:** Estimates of the effect on COVID-19 mortality of other NPIs.

| Effect on COVID-19 mortality | Description | Standardized estimate<br>(Estimated Averted Deaths /<br>Total Deaths) | Standard error | Weight<br>(1/SE) |
| --- | --- | --- | --- | --- |
| Stokes et al. (2020) | Cancelling public events (average) | 2.0% | 2.165% | 46 |
|  | *Cancelling public events before 1<sup>st</sup> death* | -1.5% | 2.324% | 43 |
|  | *Cancelling public events after 1<sup>st</sup> death* | 5.5%** | 1.994% | 50 |
| Stokes et al. (2020) | Closing public transport (average) | 0.1% | 2.478% | 40 |
|  | *Closing public transport before 1<sup>st</sup> death* | 2.0% | 2.659% | 38 |
|  | *Closing public transport after 1<sup>st</sup> death* | -1.7% | 2.282% | 44 |
| Stokes et al. (2020) | Restricting internal movement (average) | 1.4% | 2.412% | 41 |
|  | *Restricting internal movement before 1<sup>st</sup> death* | 0.5% | 2.588% | 39 |
|  | *Restricting internal movement after 1<sup>st</sup> death* | 2.3% | 2.221% | 45 |
| Leffler et al. (2020) | Lockdown of any type | 1.7% | 9.015% | 11 |

*Note: \*\* (\*) denote significance at $p < 0.01$ ( $p < 0.05$ ). For studies with several estimates related to the measure, we report all submeasures in grey but focus on the average of these measures and use the average when calculating the PWA. The average standard error is calculated as $\sqrt{(SE_1^2 + SE_2^2 + \dots + SE_n^2)/k}$ , where $k$ is the number of estimates. A negative number corresponds to fewer deaths, so -5% means 5% lower COVID-19 mortality. Stokes et al. (2020) consider two distinct time periods of 24 days to account for the changing magnitude of effects of an exponentially transmitted virus: i) 0-24 days and ii) 14-38 after the first COVID-19 death. We report both of them and use the average in the precision-weighted average.*

### 4.4 The overall effect of lockdown policies based on SIPO and specific NPIs

#### Overview of specific NPIs

Table 15 below summarizes our results on SIPO and the other specific NPIs. The central precision-weighted average in column 2 is small for most NPIs and even positive for limiting gatherings. Only masks mandates seem to have a notable effect on mortality rates but note that the estimate is based on just three studies (column 3). Column 4 presents the results of the sensitivity analyses. The precision-weighted averages are generally robust to the sensitivity analyses, and the quality-adjusted PWA generally find a less promising effect than the precision-weighted average (business closures is the only NPI where the quality-adjusted PWA is ‘preferable’ to the precision-weighted average).

**Table 15:** Summary of estimates of specific non-pharmaceutical interventions (NPIs)

| 1. NPI (number of studies) | 2. Precision-weighted average (PWA) | 3. Sensitivity analysis | 4. Quality-adjusted PWA |
| --- | --- | --- | --- |
| SIPO (12) | -2.0% | -4.1% to -1.4% | -1.8% |
| Business closure (5) | -7.5% | -9.3% to -6.6% | -11.9% |
| School closures (4) | -5.9% | -6.2% to -2.5% | -1.2% |
| Limiting gatherings (4) | 5.9% | 4.9% to 8.9% | 9.8% |
| Travel restrictions (5) | -3.4% | -4.7% to -0.4% | 1.1% |
| Mask mandates (3) | -18.7% | -19.9% to -12.5% | -18.2% |
| Public events cancellation (1) | 2.0% | n/a | 2.0% |
| Public transportation closures (1) | 0.1% | n/a | 0.1% |
| Internal movement restrictions (1) | 1.4% | n/a | 1.4% |

*Note: The table summarizes the precision-weighted averages from Table 7, Table 9, Table 10, Table 11, Table 12, Table 13, and Table 14.*

#### The average effect of lockdowns during spring of 2020

The overview in Table 15 allows us to estimate the effect of the average lockdown policies in the spring of 2020. First, we use OxCGRT data to calculate the share of the population which faced each of the NPIs from Table 15 in the spring of 2020. We focus on NPIs in the period between March 16 and April 15, 2020, which would have the greatest impact on deaths, since death rates flattened after this period. We only look at whether each NPI was mandated or not, and not whether it was strict or more lenient (e.g., we code both “2 - Require closing (only some levels or categories, e.g., just high school, or just public schools)” and “3 - Require closing all levels” as “closed schools”). This means that we overestimate the effect if stricter NPIs are more effective than more lenient NPIs. Also, as mentioned earlier, each precision-weighted average risk to be biased towards larger effect, since the estimate in each study may capture the effect of multiple (omitted) NPIs (also see footnote 67).

Based on this approach and with the bias towards overestimating the effect of lockdowns in mind, Table 16 presents the effect of the average lockdown in the spring of 2020. Our calculations suggest that the average lockdown in Europe and the United States – based on estimates for specific NPIs – reduced COVID-19 mortality rates by 10.7% (precision-weighted average) with a span in the sensitivity analysis from 0.7% (worst case) to 16.0% (best case). The quality-adjusted PWA is a 3.2% reduction in mortality rates. This precision-weighted average of 10.7% is larger than the effect found in the studies based on the OxCGRT stringency index (3.2% reduction), but still relatively small and far from the large effects promised by many epidemiological models early in the pandemic, such as Ferguson et al. (2020).^75^ To put the estimate in perspective, there were 188,542 registered COVID-19 deaths in Europe and 128,063 COVID-19 deaths in the United States by June 30, 2020. Thus, the 10.7% corresponds to 23,000 avoided COVID-19 deaths in Europe (best case: 26,000 avoided deaths, worst case: 1,000 avoided deaths) and 16,000 avoided COVID-19 deaths in the United States (best case: 25,000 avoided deaths, worst case: 1,000 avoided deaths). In comparison, there are approximately 72,000 flu deaths in Europe and 38,000 flu deaths in the United States each year.^76^

**Table 16:** Estimates of the effect on COVID-19 mortality of the average lockdown in Europe and in the United States from studies based on specific NPIs.

| 1. NPI (number of studies) | 2. Share of time with mandate (population weighted) | 3. Impact on mortality (PWA · share) | 4. Sensitivity analysis (best case to worst case) | 5. Quality-adjusted PWA |
| --- | --- | --- | --- | --- |
| SIPO (12) | 70% | -1.4% | -2.9% to -1.0% | -1.2% |
| Business closure (5) | 92% | -6.9% | -8.6% to -6.1% | -10.9% |
| School closures (4) | 97% | -5.7% | -6.0% to -2.4% | -1.1% |
| Limiting gatherings (4) | 95% | 5.6% | 4.7% to 8.4% | 9.3% |
| Travel restrictions (5) | 93% | -3.1% | -4.3% to -0.4% | 1.0% |
| Mask mandates (3) | 10% | -1.9% | -2.1% to -1.3% | -1.9% |
| Public events cancellation (1) | 95% | 1.9% | 1.9% to 1.9% | 1.9% |
| Public transportation closures (1) | 14% | 0.0% | 0.0% to 0.0% | 0.0% |
| Internal movement restrictions (1) | 64% | 0.9% | 0.9% to 0.9% | 0.9% |
| **Total impact of the average lockdown policy** |  | **-10.7%** | **-16.0% to -0.7%** | **-3.2%** |

*Note: Column 2 shows the share of the time between March 16 and April 15, 2020, where each NPI was implemented. Column 3 shows the impact of the NPI given the precision-weighted average (PWA) in Table 15 and the share of the time the NPI was implemented, see Column 2. Column 4 shows the best case (where all PWAs are in the lower end of the sensitivity analysis) and the worst case (where all PWAs are in the upper end of the sensitivity analysis), and column 5 shows the quality-adjusted PWA. The total impact of the average lockdown policy is calculated as the product of $(1 - (\text{estimates in column 3})) - 1$ . Implementing all NPIs has an impact of -26.4%.*

## 5 Concluding observations

Public health experts and politicians have – based on forecasts in epidemiological studies such as that of Imperial College London (Ferguson et al. (2020)) – embraced compulsory lockdowns as an effective method for arresting the pandemic. But, have these lockdown policies been effective in curbing COVID-19 mortality? This is the main question answered by our meta-analysis.

Adopting a systematic search and title-based screening, we identified 1,220 studies, which potentially look at the effect of lockdowns on mortality rates. To answer our question, we focused on studies that examine the actual impact of lockdowns on COVID-19 mortality rates based on registered cross-sectional mortality data and a counterfactual difference-in-difference approach. Out of the 1,220 studies, 32 met our eligibility criteria, and standardized estimates for our meta-analysis could be calculated for 22 of the eligible studies.

### 5.1 Conclusions

Overall, our meta-analysis fails to confirm the notion that lockdowns – at least in the spring of 2020 – had a large, significant effect on mortality rates. Studies examining the relationship between lockdown strictness (based on the OxCGRT stringency index) find that the average lockdown in Europe and the United States only reduced COVID-19 mortality by 3.2% compared to the most lenient COVID-19 policy. Shelter-in-place orders (SIPOs) were also ineffective. They only reduced COVID-19 mortality by 2.0%. Based on nine specific NPIs, we estimate that the average lockdown in Europe and the United States in the spring of 2020 reduced mortality by 10.7%. The 3.2% to 10.7% correspond 6,000-23,000 avoided deaths in Europe and 4,000-16,000 avoided deaths in the United States. In comparison, there are approximately 72,000 flu deaths in Europe and 38,000 flu deaths in the United States each year.^77^ Thus, lockdowns in Europe and the United States on average saved lives corresponding to 9%-35% of an average flu season.

Of the specific NPIs, closing non-essential businesses seems to have had some effect (reducing COVID-19 mortality by 7.5%), which is possibly related to the closure of bars. We find that mask mandates had the largest effect (reducing COVID-19 mortality by 18.7%), but the estimate is based on just three studies with heterogeneity in the definition of the mandate. Limiting gatherings were counterproductive and *increased* mortality by 5.9%.

Our measured meta-results are supported by the natural experiments we have been able to identify through our work and by searches in the abstract and citation database Scopus (see Table 17).

**Table 17:**
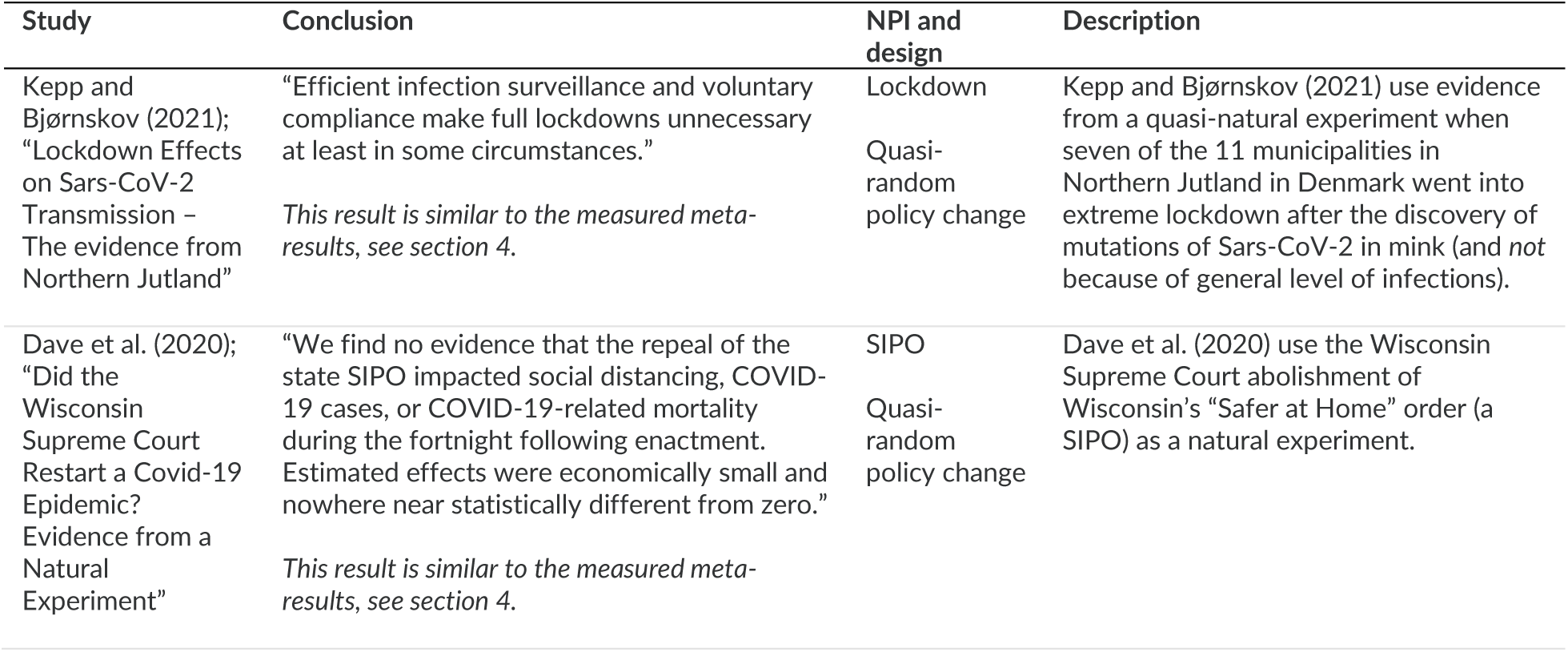
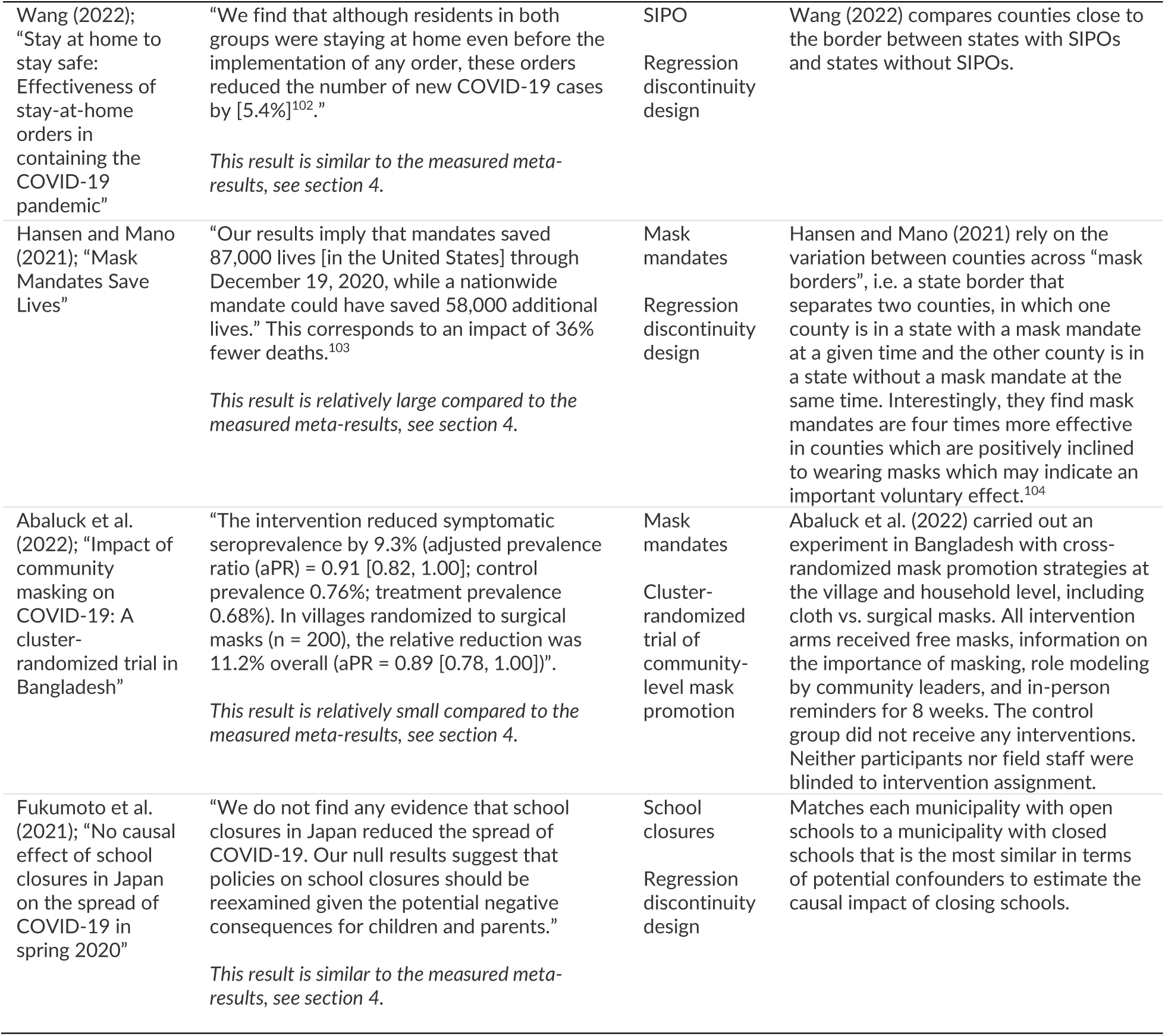
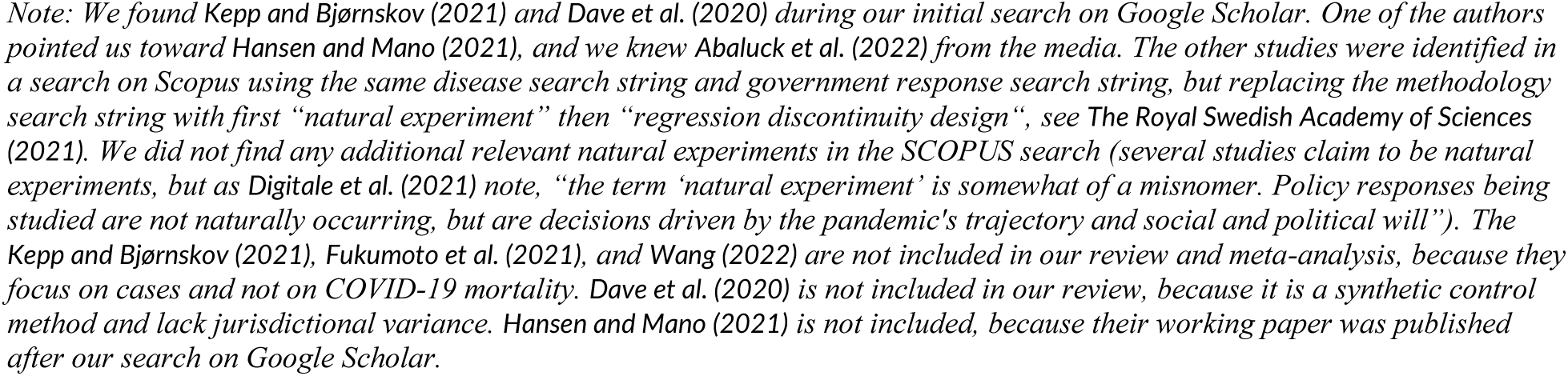
The results in the identified natural experiments are similar to our measured meta-results.

Overall, our meta-analysis supports the conclusion that lockdowns – at least in the spring of 2020 – had little to no effect on COVID-19 mortality.

Throughout the meta-analysis, we have focused on the precision-weighted average as our primary indicator of the efficacy of lockdowns. However, as shown Figure 10, the overall conclusion holds regardless of which studies or measures one chooses to emphasize. The figure presents the effect on mortality in the United States based on the measured estimates from all stringency studies as well as our two central measured estimates for the effect of lockdowns in the spring of 2020 (the precision-weighted average from the stringency studies in Table 5 and the estimate based on specific NPIs in Table 16). We have added the max and min forecasted estimates from Ferguson et al. (2020) for comparison. Even if we cherry-pick the most preferable empirical estimate from Fuller et al. (2021), the effect on mortality is far from the effects promised based on Ferguson et al. (2020) and corresponds to the mortality from less than two influenza seasons.^78^

**Figure 10:**
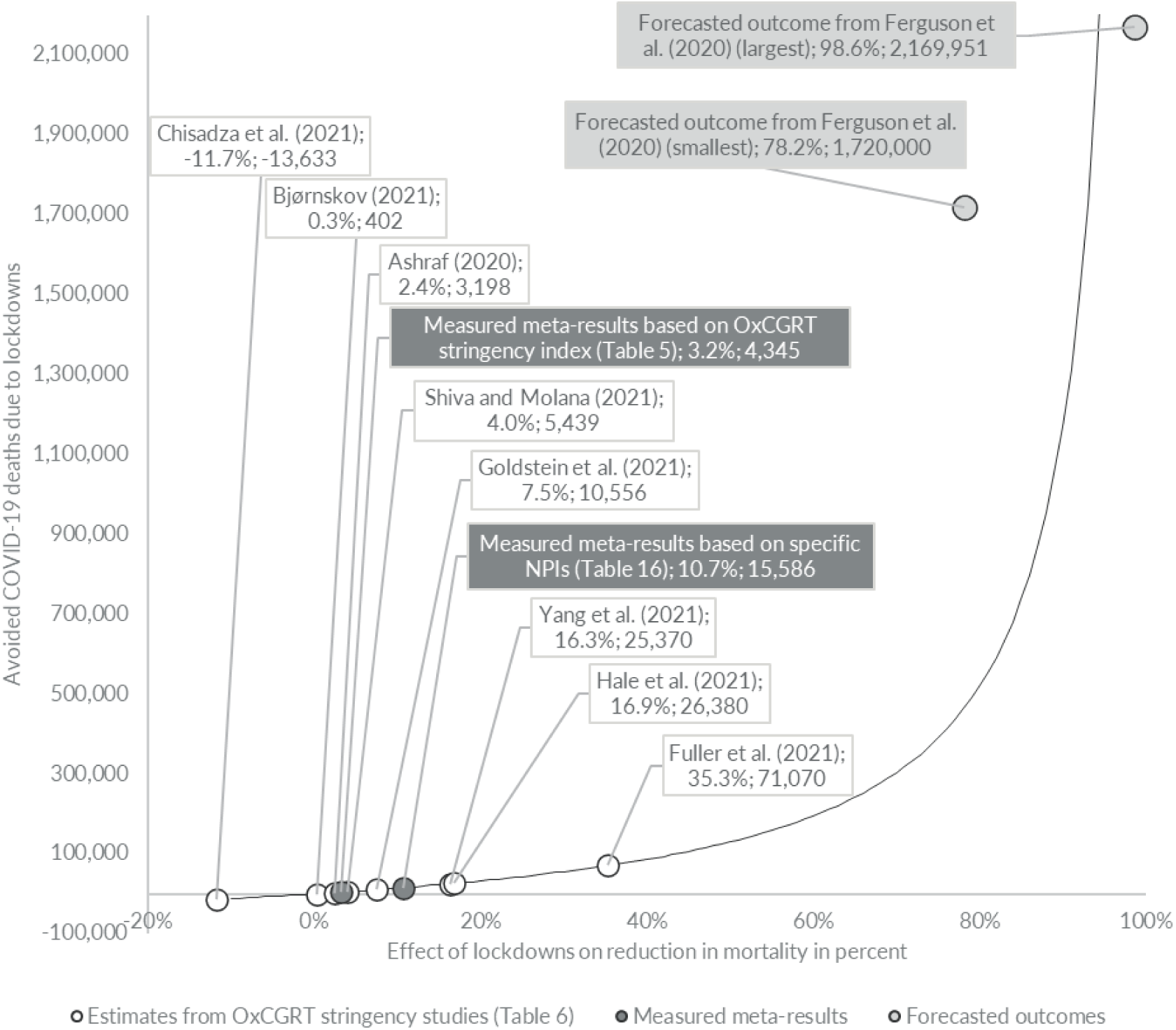
Divergence between avoided number of deaths in the United States as measured by the meta-results, studies based on the OxCGRT stringency index, and the forecasted outcome from Imperial College London. Note: The estimates in the group “Estimates from stringency studies” are from Table 5. For the group “Meta-study PWA” the estimate for “PWA stringency studies” is from Table 5, and “average lockdown spring 2020” is from Table 16. Both estimates illustrate the effect of the average lockdown in Europe and United States in the spring of 2020. The effect of lockdowns on total mortality based on the meta-study’s precision-weighted averages (PWA) is calculated as total COVID-19 deaths by July 1, 2020 (128,063 COVID-19 deaths) × (1/(1-PWA)-1). The relative effect of lockdowns on total mortality based on Ferguson et al. (2020) is calculated as the largest and smallest predicted relative effect multiplied with their mortality estimate of 2.2 million deaths in a “do nothing”-scenario in United States. The estimates from Ferguson et al. (2020) is for a two year period, but the relative effect is largest early in the pandemic.

#### Limitations

Our study has a number of limitations. Most importantly, we cannot say whether NPIs are crucial as a means of signaling the danger of the pandemic. It is possible that – even if stricter lockdowns and each individual NPI are ineffective –the government ‘doing something’ is necessary to spur voluntary behavioral changes. The question is whether people had understood how serious the situation was if governments had not resorted to measures that went beyond the usual. If the answer to this question is that some sort of lockdown was indeed necessary (and there is some evidence suggesting this)^79^, the next obvious question is: How little intervention does it take to send the necessary signal? Our results suggest that the answer to this question is “relatively little”, but how to send the best signal with as little cost to society as possible, is an obvious area for future research.

Another limitation is the limited number of studies. This is especially true for the specific NPIs where our estimates are often based on 3-5 studies. We hope that our work will inspire more researchers to think about ways to examine the effect of specific NPIs so future meta-analyses have more studies to rely on.

Also, we do not analyze the role of timing. As pointed out in the section 2.2, p. 16, and in section 5.2.4, p. 71, we believe that many studies examining the role of timing are fundamentally flawed because they do not distinguish between voluntary behavioral changes and lockdowns (i.e. mandatory behavioral changes). We also point out that even if there *is* an optimal timing, we cannot be sure that democratic governments will ever be able to react in time on this information and implement the NPIs accordingly. However, these are problems that may be addressed by future research. In any case researchers should find ways to distinguish between the effects of voluntary behavioral changes (possibly spurred by signaling) and those of lockdowns.

Our meta-analysis shows that some NPIs may have a measurable effect on mortality. Mask mandates especially look relatively effective, although our estimate is based on just three studies and the 18.7% is still a relatively low effect compared to the effects promised by many epidemiological models early in the pandemic.^80^ However, that an NPI has a measurable effect on mortality is a necessary – but not sufficient – requirement to make the policy beneficial and desirable. Also, there is some evidence that mask recommendations can be sufficient to reap much of the effect of mask mandates. Thus, future research is needed to estimate the broader costs of mask mandates – including effects on welfare, trust etc. – before one can conduct an actual cost-benefit analysis, which can answer whether mask mandates are a desirable policy.

We only look at mortality. It is possible that there are other benefits related to lockdowns which are not captured in the studies looking at mortality rates. For example, Banholzer et al. (2022) believe that “interventions that reduce the number of new infections can have downstream effects on various outcomes, including disease-related deaths, cases of severe illness and hospitalizations, cases with long-term health effects after infection, the efficiency of testing and contact tracing” etc. While this may be true, we believe it is unlikely that the effect of lockdowns on infections has been so different from the effect of lockdowns on mortality that it changes the overall conclusion. Even if the effect on infections is two or three times larger than the effect on deaths, the overall effect is limited and far from the effect promised based on model studies, see Figure 10. However, only future research can tell whether this immediate assessment holds true.

We also restrict our search strategy to studies using a “counterfactual difference-in-difference approach”. We believe difference-in-difference studies are better suited than other widely used empirical methods to examine the true effect of lockdowns because they allow us to leave out the effect of voluntary behavior changes. There is, however, no doubt that the results from other study methods are of great interest because they can give us insights with regard to e.g., the importance of voluntary behavior changes. We welcome future research on these other methodologies.

### 5.2 Discussion

#### 5.2.1 Conclusions are in line with other reviews

Overall, we conclude that stricter lockdowns are not an effective way of reducing mortality rates during a pandemic, at least not during the first wave of the COVID-19 pandemic. Our results are in line with the World Health Organization Writing Group (2006), stating, “Reports from the 1918 influenza pandemic indicate that social-distancing measures did not stop or appear to dramatically reduce transmission […] In Edmonton, Canada, isolation and quarantine were instituted; public meetings were banned; schools, churches, colleges, theaters, and other public gathering places were closed; and business hours were restricted without obvious impact on the epidemic.”

Our findings are also in line with the conclusion in Allen (2021): “The most recent research has shown that lockdowns have had, at best, a marginal effect on the number of Covid-19 deaths.” Poeschl and Larsen (2021) conclude that “interventions are generally effective in mitigating COVID-19 spread.” but 9 of the 43 (21%) results they review find “no or uncertain association” between lockdowns and the spread of COVID-19^81^, suggesting that the impact of lockdowns is limited and not that far from zero, which contradicts their conclusion.^82^ Based on two interrupted time series studies, Iezadi et al. (2021) find that overall NPIs reduced daily ICU admissions by 16.5%. Mendez-Brito et al. (2021) find that school closing is the most effective measure, although only 14 out of 24 studies (58%) found an association between school closures and number of cases, suggesting a limited effect.^83^ Herby (2021a) concludes that “mandated behavior changes accounts for only 9% (median: 0%) of the total effect on the growth of the pandemic stemming from behavioral changes. The remaining 91% (median: 100%) of the effect was due to voluntary behavior changes.”

The findings contained in Johanna et al. (2020) are in contrast to our results. They conclude that “for lockdown, ten studies consistently showed that it successfully reduced the incidence, onward transmission, and mortality rate of COVID-19”. The driver of the difference is three-fold. First, Johanna et al. (2020) include modelling studies (10 out of a total of 14 studies), which we have explicitly excluded. Second, they included interrupted time series studies (3 of 14 studies), which we also exclude. Third, the only study using a difference-in-difference approach (as we have done) is based on data collected before May 1^st^, 2020. We should mention that our results indicate that early studies find relatively larger effects compared to later studies.

#### 5.2.2 Causality or correlation?

As pointed out by Bjørnskov (2021a), there is a potential endogeneity problem (also referred to as reverse causality) “which derives from the nature of political reactions to the virus that could rely on the reported number of infections. If an increase in the reported infection rate leads government to introduce lockdown policies, and if a declining reported infection rate subsequently leads them to ease the lockdown, the estimated association between policy stringency and mortality is biased.”

Several studies explicitly claim that they examine the actual causal relationship between lockdowns and COVID-19 mortality. Some studies use instrumental variables^84^, lagged dependents^85^, or other techniques to establish a causal relationship, while others make causality probable using arguments.^86^ Sebhatu et al. (2020) show that government policies are strongly driven by the policies initiated in neighboring countries rather than by the severity of the pandemic in their own countries.

In short, Sebhatu et al. (2020) show that it is not the severity of the pandemic that drives the adoption of lockdowns, but rather the propensity to copy policies initiated by neighboring countries. Sebhatu et al. (2020) also find that the death rate predicts the stringency of countries’ policy adoptions, but the effect is small explaining only 2.1 stringency points on average (in comparison, the gap between the strictest and most lenient lockdown in Europe was between 67 to 92 stringency points in the period from March 16 to April 15, 2020).^87^ The very low mortality rates on the day of lockdown (defined as the day when the OxCGRT stringency index exceeds 60) is illustrated in Figure 11.

**Figure 11:**
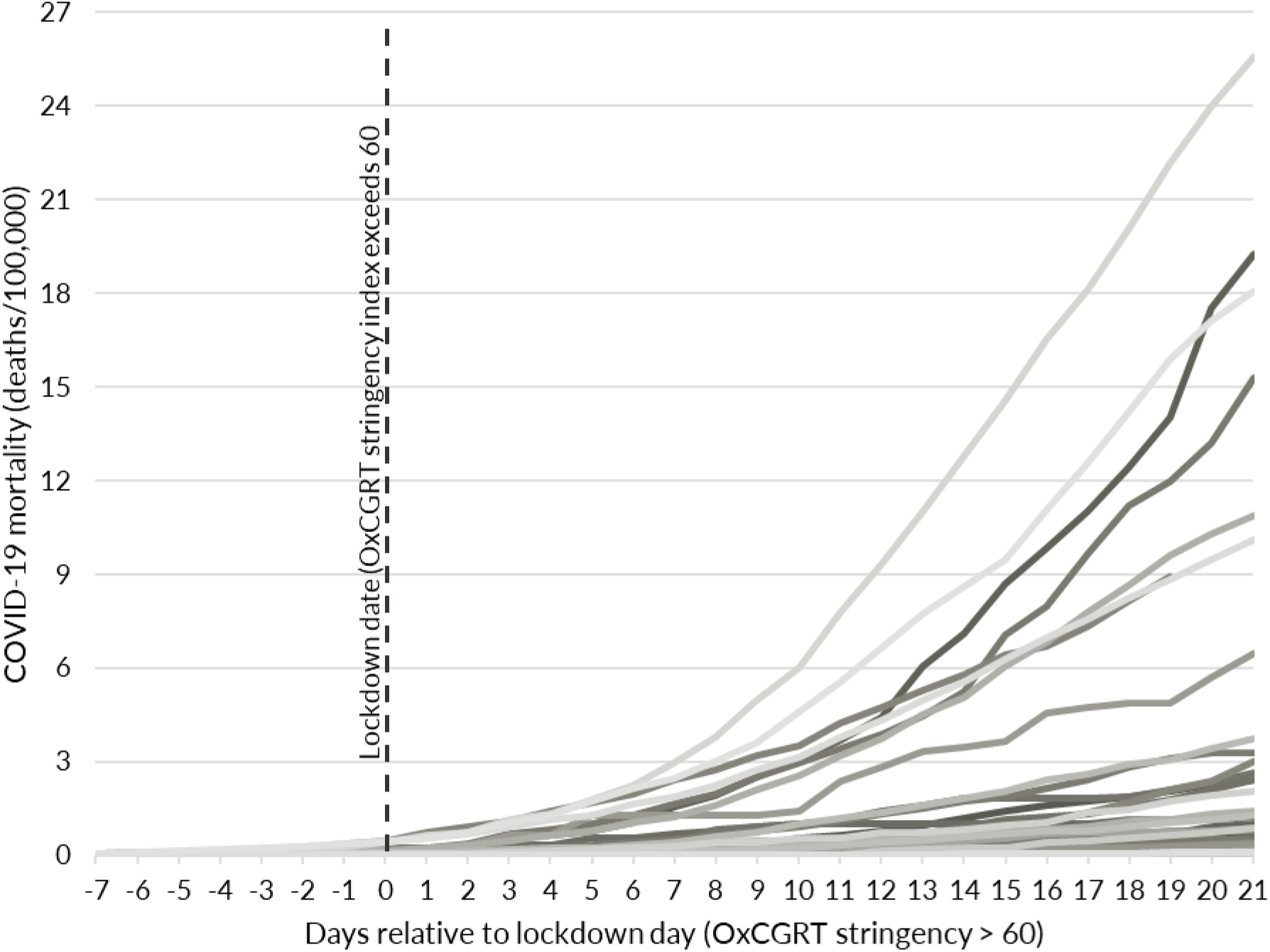
Mortality rates in European countries were very low prior to lockdown-decisions. Source: Our World in Data (2022). Note: The figure shows the development in mortality in 37 European countries with more than 100,000 inhabitants. Day 0 is the day where the OxCGRT stringency index crosses 60

Note that decisions were often taken days in advance of implementation (e.g. Denmark announced the lockdown March 11, 2020, but schools closed 5 days later (March 16) and businesses seven days later (March 18)), where mortality rates were even lower (and knowledge even more sparse) and the reporting of deaths was in some cases delayed. This lag between decision and implementation is non-negligible. On average, mortality rates were about 50% lower three days prior to lockdown and 75% lower five days prior to lockdown.

Also, most eligible studies examine the first wave, where most countries – due to limited testing capabilities – had little information about the progress of the pandemic, and thus, the policy response in a given country is unlikely to be greatly caused by the severity of the pandemic. Bjørnskov (2021a) points to governments’ ability to react quickly and note that “although one might think that policy making reacts quickly to changing mortality during an emergency, exploring the determinants of changes in the stringency indices reveals that an increase in the contemporaneous mortality or an increase in the reported number of Sars-CoV-2 cases was not associated with stricter lockdown measures” and conclude that “it is highly unlikely that there is a substantial endogeneity problem in the following as mortality changes only affect policy changes with a three-week lag, and as policy changes cannot affect the mortality rate before another two to three weeks have passed. As such, any bias is likely to be small and practically negligible.” Finally, 11 of the 22 studies in the meta-analysis handles the causality question, but their results are not much different from the other 11 studies, implying that causality is not a major problem.

Hence, we believe there is a strong case for a causal relationship in our results and that what the studies examine is the effect (of the strictness) of lockdowns on mortality and not the opposite (mortality rates’ effect on (the strictness of) lockdowns, although the issue can never be finally settled with observational studies.^88^

#### 5.2.3 Why are the effects of lockdowns limited?

Our main conclusion invites a discussion of some issues. Our review does not point out *why* lockdowns did not have the effect promised by the epidemiological models of Imperial College London (e.g., Ferguson et al. (2020)). But it is evident that modelers around the globe failed to accurately forecast the development of the pandemic.

One example is the projections of COVID-19 inpatients that were published during the Danish negotiations of a reopening in the spring of 2021. Figure 12 below compares the projections to actual data following the reopening. Not only did the modelers fail to project the number of COVID-19 inpatients following the reopening of the economy, the actual outcome was below even the most optimistic lockdown scenario (the lower bound of the grey shaded area). It should be noted that this forecasting failure was in no way unique to the Danish health authorities. Most health authorities and expert modelers failed to correctly project the development of the pandemic.^89^

**Figure 12:**
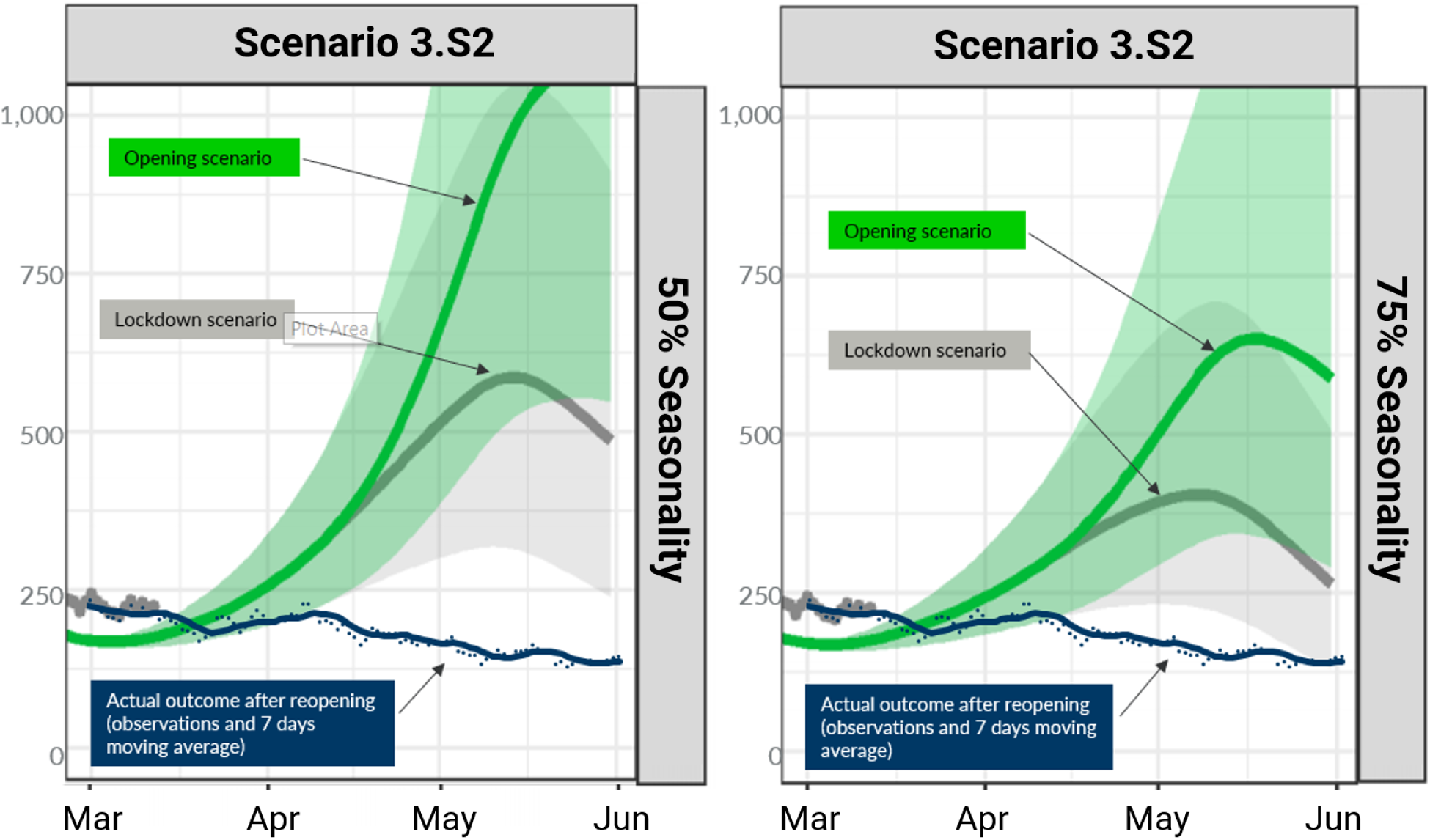
Model forecasts of COVID-19 inpatients with and without reopening compared to actual outcomes (Denmark, 2021) Source: Statens Serum Institut (2021) and Danmarks Statistik (2022). Note: The green line shows the projections if the economy was reopened, while the grey line shows the projections with continued lockdown. The blue dots and line show the actual development after the economy was reopened. “50% sæson”and “75% sæson” denotes how much seasonal variation is included. Text has been translated from Danish to English.

We propose four factors that might explain the difference between our conclusions and the views embraced by some epidemiologists.

##### People respond voluntarily to dangers

First, people respond to dangers outside their door when they are aware of it. When a pandemic rages, people engage in social distancing regardless of what the government mandates. In economic terms, you can say that the demand for costly disease prevention efforts like social distancing and increased focus on hygiene is high when infection rates are high.^90^ On the contrary, when infection rates are low, the demand is low and it may even be morally and economically rational not to comply with mandates like SIPOs, which are difficult to enforce.

Herby (2021a) reviews studies that distinguish between mandatory and voluntary behavioral changes. He finds that – on average – voluntary behavioral changes are 10 times as important as mandatory behavioral changes in combating COVID-19. Andersen et al. (2020) find that consumer spending fell almost as much in Sweden as in Denmark despite Sweden having a very limited lockdown.^91^ Interestingly, the response in Sweden was especially large among 70+year-olds and even larger than in Denmark – possibly due to the larger outbreak in Sweden. Chetty et al. (2020) show that high-income individuals reduced spending sharply in mid-March 2020, particularly in areas with high rates of COVID-19 infection and in sectors that require physical interaction. By comparing counties with and without restrictions, Goolsbee and Syverson (2021) conclude that only 7%-points of the 60%-point decline in business activity could be attributed to legal restrictions, and that the shift was highly tied to the number of COVID deaths in the county. Most of the decline resulted from consumers voluntarily choosing to avoid stores and restaurants. The point from Andersen et al. (2020), Chetty et al. (2020) and Goolsbee and Syverson (2021) is illustrated in below figure from Maas (2020).

Gupta et al. (2020) find that “information-based policies and events such as first cases had the largest effects”. They also find that SIPOs did not affect social distancing (as measured by a mixing index which – based on cell phone data – measured the exposure of a smart device to other devices). At a first glance this seems to be in conflict with the findings by Joshi and Musalem (2021) who find that SIPOs increases time spent at home, but one obvious explanation is that people stop mixing voluntarily before they are mandated to stay at home, so the SIPOs do not increase social distancing (and thus, reduce infections) but only increase time spent at home. After all, there are several ways you can leave your house without mixing with others. Bor et al. (2021) find that intrinsic motivations related to the severity of the pandemic (as measured by national case numbers) play a significant role when citizens increase their attention to the health authorities’ advice during an epidemic.

These voluntary behavioral changes may also explain why epidemiological model simulations such as Ferguson et al. (2020) – which do not model behavior endogenously^92^ – fail to forecast the effect of lockdowns. Voluntary behavioral changes also explain why the flu almost disappeared in Denmark in March 2020 before a single restriction was implemented. In Denmark, schools closed by March 16^th^ and businesses by March 18^th^. But at these dates the share of positive influenza tests in Denmark had dropped from about 25% – a level which had been more or less constant for two months – before March 11^th^, when the Danish prime minister held a press conference, to 5%-10% (Statens Serum Institut (2020)). As we showed in Figure 8, p. 20, the same pattern was seen in Norway and Sweden.

Similar drops in the share of positive influenza tests were seen in Norway and Sweden (Emborg et al. (2021), also see Figure 8 on p. 72). In the United States, Ziedan et al. (2020) find that “aggregate trends in outpatient visits show a 40% decline after the first week of March 2020, only a portion of which is attributed to state policy.” Tsai and Tzu-Ting (2021) find similar results for Taiwan. Overall, we believe that Allen (2021) is correct, when he concludes, “The ineffectiveness [of lockdowns] stemmed from individual changes in behavior: either non-compliance or behavior that mimicked lockdowns.”

##### Mandates only regulate a fraction of our potential contagious contacts

Second, mandates only regulate a fraction of our potential contagious contacts. Figure 14 illustrates infection locations in Germany during the early pandemic. The figure shows that most of the infections in Germany assigned to an outbreak (defined as at least two cases) occurred in homes (including homes for the elderly), hospitals, and workplaces that were not subject to general restrictions applied throughout the society and where potentially effective interventions, such as handwashing, coughing etiquette, ventilation, distancing etc. could neither be regulated nor enforced but relied solely on voluntary behavioral changes. In total, 77% of infections occurred in homes, hospitals, and workplaces, and the share was – despite the first weeks of the pandemic when awareness was low – large (above 60%) despite variations in the use of NPIs.

**Figure 13:**
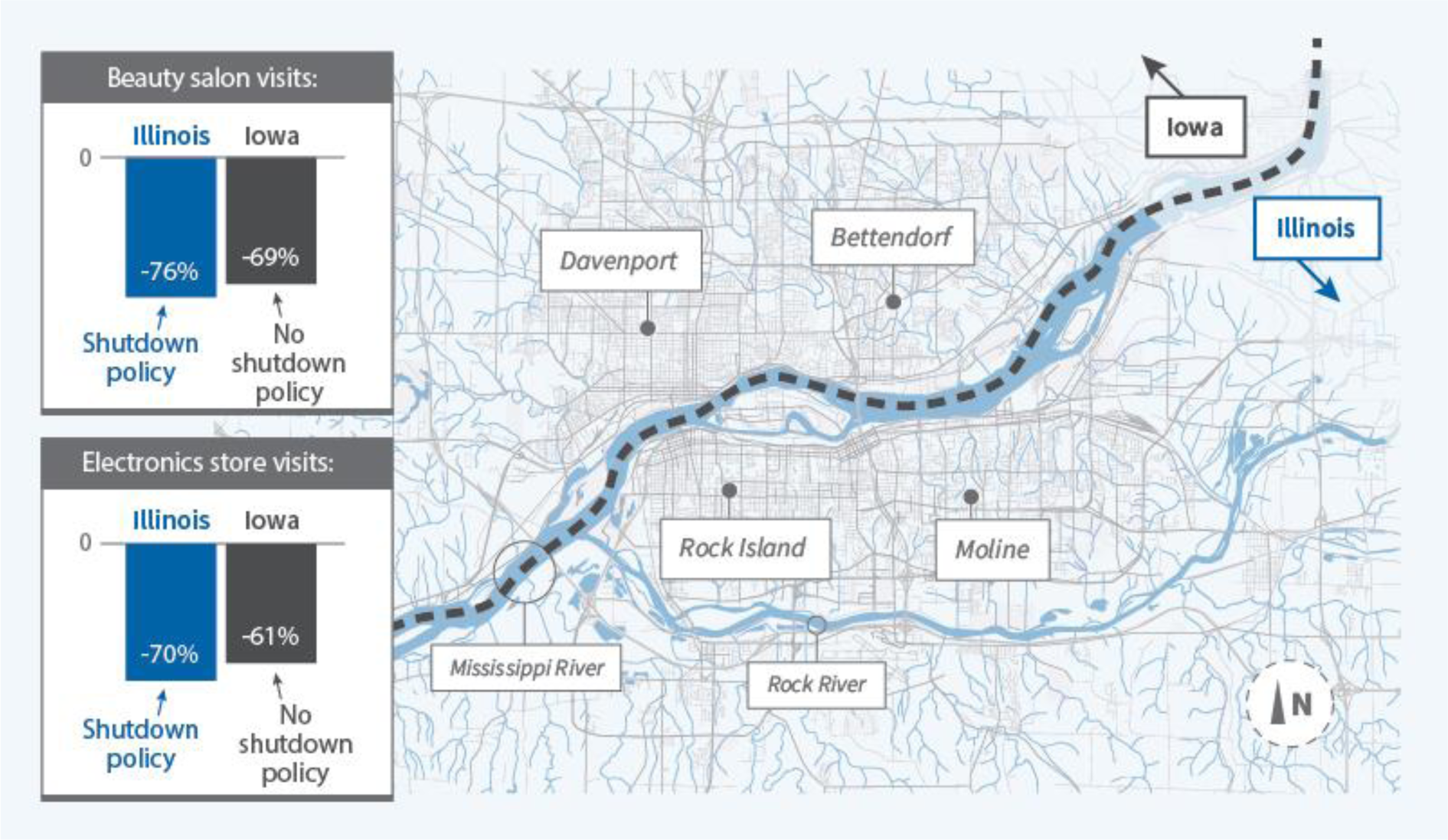
Lockdown-policy differences and consumer activity in Iowa and Illinois. Source: Maas (2020) based on data from SafeGraph.

**Figure 14:**
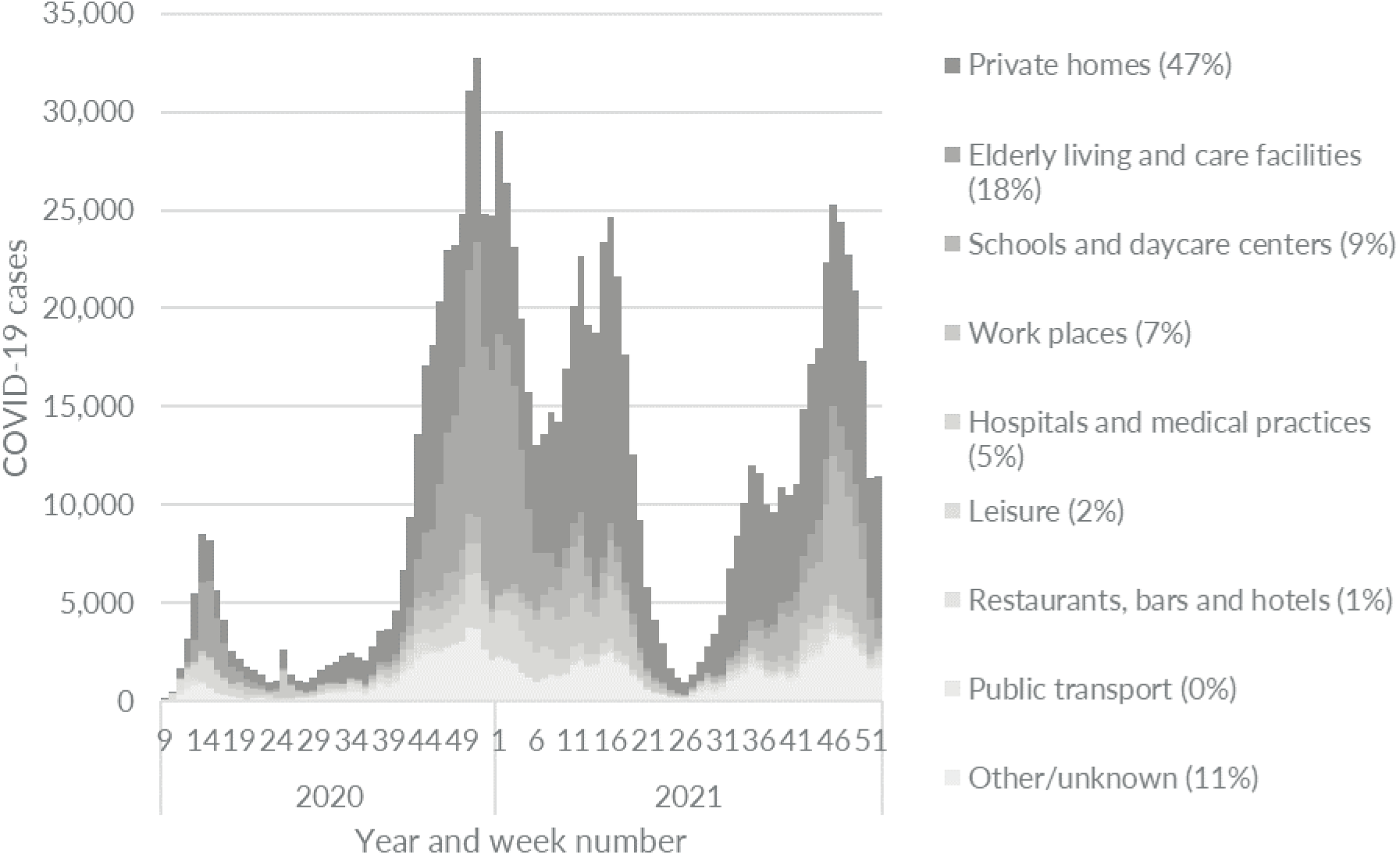
Homes, hospitals and workplaces were the main drivers of infections in Germany and the location for 77% of all infections. Source: Robert Koch Institut (2022) Note: Laboratory-confirmed COVID-19 cases assigned to an outbreak by infection setting and reporting week. Number in parentheses is the share of total cases during the covered period. The data covers COVID-19 outbreaks with two or more cases which includes about 15% of all cases of infection in Germany.

The data in Figure 14 only covers COVID-19 outbreaks with two or more cases, which includes about 15% of all cases of infection in Germany. But data from other countries show similar patterns. Lee et al. (2020) write that “early contact tracing studies and a large study of more than 59,000 case contacts in South Korea found household contacts to be greater than six times more likely to be infected with SARS-CoV-2 than other close contacts” and Zhao et al. (2020) find that “69.2% of total cases were clustered in a home, apartment or residential estate.”

In a Danish matched case-control study based on data from November 2020, Munch et al. (2022) find that contact with an infected person at home or at work was a substantial risk factor.^93^ They find no infection risk associated with community exposures such as shopping at supermarkets, travel by public transport, dining at restaurants, and private social events with few participants. The results were confirmed by Lendorf (2021) who found that 69% were infected at places with no general restrictions. ^94^ Both Munch et al. (2022) and Lendorf (2021) are based on data from a period with relatively few infections and some restrictions (gatherings limited to 10 persons, restaurants and bars etc. had to close at 10 pm, and face masks were mandatory indoors, except when seated). Nevertheless, their results illustrate that people in countries like Denmark, Finland, and Norway – countries that allowed people to go to work, use public transport, and meet privately at home during the pandemic – had ample opportunities to legally meet with – and get infected by – others. Still, these countries experienced relatively low COVID-19 mortality rates.

##### Behavioral responses may counteract any initial effect of lockdowns

Third, even if lockdowns are successful in initially reducing the spread of COVID-19, the behavioral responses may counteract the effect as people respond to the lower risk by changing behavior. As Atkeson (2021) points out, the economic intuition is straightforward. If closing bars and restaurants causes the prevalence of the disease to fall toward zero, the demand for costly disease prevention efforts like social distancing and increased focus on hygiene also falls towards zero, and the disease will return.^95^ As pointed out by Deaton and Cartwright (2018), randomization “does not relieve us of the need to think about (observed or unobserved) covariates”. Also, this kind of second-order behavior response may also explain why closing down non-essential businesses simply reallocates consumer visits away from “nonessential” to “essential” businesses, as shown by Goolsbee and Syverson (2021), with limited impact on the total number of contacts.^96^And this probable behavior response to changes in infection levels limits the knowledge we can obtain from randomized control trials examining specific NPIs (if for example, masking children in schools reduces the infections among children and teachers, this does not necessarily imply that masking children reduces infection rates overall).

Also, Joshi and Musalem (2021) find that the effect of SIPOs on mobility decreases as time passes and infection rates drop.

##### Some NPIs may have led to unintended consequences

Fourth, unintended consequences may play a larger role than recognized. We already pointed to the possible unintended consequence of SIPOs, which may isolate an infected person at home with his/her family where he/she risks infecting family members with a higher viral load, causing more severe illness. But often, lockdowns have limited peoples’ access to safe (outdoor) places such as beaches, parks, and zoos, or included outdoor mask mandates or strict outdoor gathering restrictions, pushing people to meet at less safe (indoor) places. Indeed, we do find evidence that limiting gatherings was counterproductive and increased COVID-19 mortality by 5.9%, see Table 11.

#### 5.2.4 Objections to the results of the meta-analysis

Our results and conclusions go against the conventional wisdom that lockdowns were effective in reducing COVID-19 mortality, and, indeed, the first version of our study did spawn a wide range of objections to our measured meta-results. We address the most important of these in this section.

##### The “timing of lockdowns is crucial”-objection

One objection to our conclusions is that we do not look at the role of timing. If timing is very important, differences in timing may empirically overrule any differences in lockdowns. We first note that this objection does not necessarily contradict our results. If timing is very important relative to strictness, this suggests that well-timed, but very mild, lockdowns should work as well as, or better than, less well-timed but strict lockdowns. This is not in contrast to our conclusion, as the studies we reviewed analyze the effect of lockdowns when compared to doing very little (see section 3.1 p. 23 for further discussion). However, there is little solid evidence supporting the timing thesis, because it is inherently difficult to analyze (see section 2.2 p. 16 for further discussion).

In Figure 7 we showed that all countries and states that were hit late by the pandemic experienced low COVID-19 mortality rates. This pattern – where areas hit late in the pandemic also have lower deaths rates – was also found during the Spanish Flu in 1918. Figure 15 below shows how cities in the United States that were hit early by the Spanish Flu in the autumn of 1918 also experienced large excess mortality. The data is from Hatchett et al. (2007) and Markel et al. (2007), who both conclude that the low excess mortality was due to lockdowns early in the pandemic. But, as the figure clearly shows, cities which implemented lockdowns early in the pandemic were also hit relatively late compared to other cities making it difficult to assess whether the measured effect of early lockdowns is related to lockdowns or is related to voluntary behavior changes instead.

**Figure 15:**
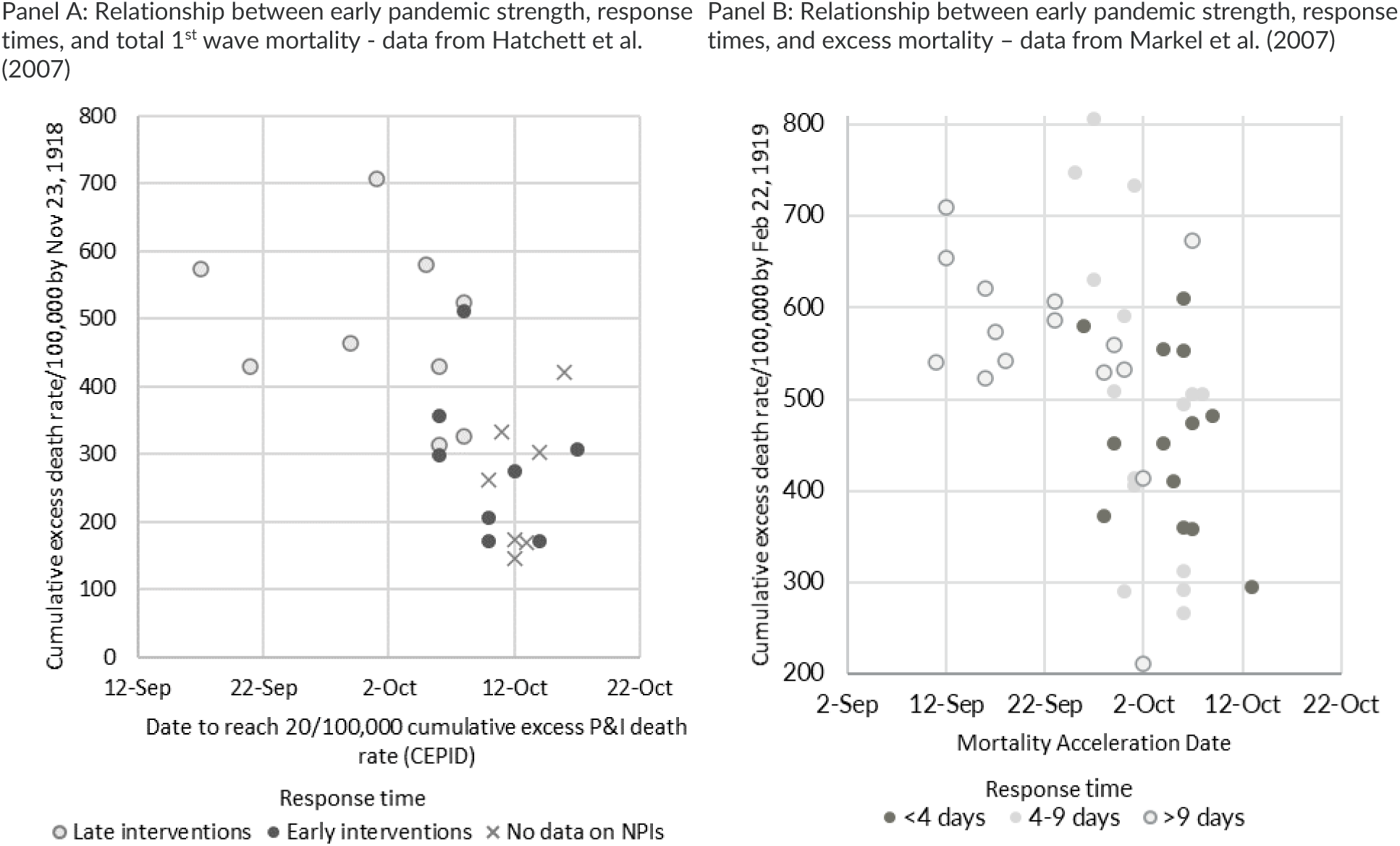
Cities hit late by the Spanish Flu in 1918 experienced lower excess mortality. Source: Hatchett, et al. (2007) and Markel et al. (2007) Note: The figure illustrates that cities that were hit late by the 1918 Spanish Flu pandemic generally experienced lower excess death rates despite response times.

Hatchett et al. (2007) touch on the subject by noting that “in addition, cities whose epidemics began later tended to intervene at an earlier stage of their epidemics, presumably because local officials in these cities observed the effects of the epidemic along the Eastern seaboard and resolved to act quickly”. But if local officials observed the effects on the east coast, many ordinary people probably did too, spurring – or at least laying the groundwork for – voluntary behavioral changes. Indeed, if we exclude cities which were hit early, the average excess mortality in Markel et al. (2007) is similar across response times indicating that information and voluntary behavioral changes could be driving their results.^97^

Also, even if it can be empirically stated that a well-timed lockdown is effective in combating a pandemic, it is doubtful that this information will ever be useful from a policy perspective. If lockdowns are effective as long as they are well timed, our results – which show the average effect of lockdowns in Europe and the United States – show that governments, on average. were unable to time lockdowns properly to obtain a substantial effect on mortality.^98^ The problem of proper timing is well known from the debate about discretionary economic stabilization policies. Selecting the proper timing for such measures have proved a disappointment.^99^ Thus, discretionary approaches have largely been abandoned for rule-based stabilization policies.

##### The “your results do not apply to later waves and variants”-objection

Another objection has been that the studies included in the meta-analysis cover, for the most part, the first wave, and that one can imagine that lockdowns may be effective against later waves or variants. This objection is a hypothesis, and only future research can show if this hypothesis is true or false. We note that even if future research falsifies this hypothesis, yet another hypothesis can then be proposed: that – again – the historical evidence is not relevant for future waves and variants. In the end, it will be a political discussion how heavy such speculations should weigh when making decisions, when historical data find limited effects of lockdowns.

##### The “there are too few studies to know for sure”-objection

Several commentators have pointed out that our conclusion is based on relatively few studies. While more studies are always better, we have included all existing empirical evidence and covered far more studies than needed to e.g., bring a new drug to the market.^100^ It is worth noting that optimal sample sizes can be and are surprisingly small in number (see Hanke and Mehrez (1979)). Communicable diseases can be handled using pharmaceutical interventions and/or non-pharmaceutical interventions. From a scientific and political perspective, the evidence required to implement either of these interventions should not differ. Hence, it is inconsistent to have a political regime where pharmaceutical interventions *may* only be used if one can prove they are *effective* and that negative side effects are *small*, while *non-*pharmaceutical interventions *will* be used unless one can prove they are *ineffective* and that negative side effects are *large*.

##### The “we cannot be sure without randomized control trials”-objection

Another objection is that our findings are not based on randomized control trials (RCTs). The obvious response is that a RCT has never been conducted for lockdowns. Therefore, we are limited to observational studies.

We agree that, preferably, the effect of lockdowns should be tested using randomized control trials (RCT), although Deaton and Cartwright (2018) argue that “the lay public, and sometimes researchers, put too much trust in RCTs over other methods of investigation”. Unfortunately, there are very few classic RCT studies except for some little relevant mask studies (see Hirt et al. (2022)). Given the lack of RCTs, observational studies are our best way to know if lockdowns work. We only have one source of evidence: history, with all its covariates and missing/bad data. If we reject that source, we have nothing to rely on. RCT is not the only kind of research that can improve policy, and, as Caplan (2022) puts it: “What’s the empirical evidence that RCTs actually improve policy?”

However, we have – after we, among other things, conducted a search on Scopus^101^ – knowledge of a few “RCT like” studies which use natural experiments to examine the effects of lockdowns, mask mandates, school closures and SIPOs. The studies are described in Table 17 and overall find conclusions very similar to the measured meta-results.

Overall, the results from the natural experiments in Table 17 are similar to our own conclusions. They, for the most part, only find marginal effects of lockdowns and NPIs, except for Hansen and Mano (2021) who examine mask mandates. These studies do not live up to our eligibility criteria and are not included in the meta-analysis (see notes to Table 17) but should be noteworthy given their credibility.

##### The “every time a country has locked down, the mortality rate has dropped”-objection

We agree that the general pattern has been that mortality rates usually – but not always – drop after lockdowns are imposed. However, this does not imply causality, as people voluntarily change behavior when responding to information. Also, while this has been the general pattern, there are examples which, as a minimum, question the causality in the argument. Figure 15 shows daily deaths in Slovenia and Slovakia during the 2020/21 winter. Both Slovenia and Slovakia introduced strict lockdowns in late October 2020. On October 24^th^, Slovakia had issued SIPOs, limited gatherings, banned indoor eating and drinking at restaurants, closed schools, and issued mask mandates.^105^ Nevertheless, the death toll continued to rise, and the death rate stayed above pre-lockdown levels for at least six months. Slovenia experienced a more classic wave, but the daily death rate did not peak until 6 weeks after the lockdown, making it unlikely to be caused by the lockdown (usually, it takes 3-4 weeks from infection to death.)

##### The “what about Zero-Covid-countries”-objection?

Some have pointed to countries like Australia and New Zealand, which have followed a strategy with very strict lockdowns as a response to even relatively few infections (known by many as a “Zero-Covid”-strategy). For example, Melbourne’s SIPO in response to the Delta strain lasted 262 days.^106^ Comparing COVID-19 mortality rates in Australia to mortality rates in Europe and the United States, this Zero-Covid strategy appears to be effective when measured by COVID-19 mortality rates. But as illustrated in Figure 17, the immediate effectiveness is less obvious compared to other island countries of which at least some have used more lenient COVID-19 policies (e.g. by December 31, 2021, Iceland had never issued a SIPO and only closed schools for 82 days in total, whereas New Zealand (/Australia) had issued a SIPO for 191 (/363) days and closed schools for 200 (/396 days).^107^ We therefore caution against attributing the low mortality rates in New Zealand and Australia to strict lockdowns when more obvious explanations – such as being island countries – may explain the differences.

**Figure 16:**
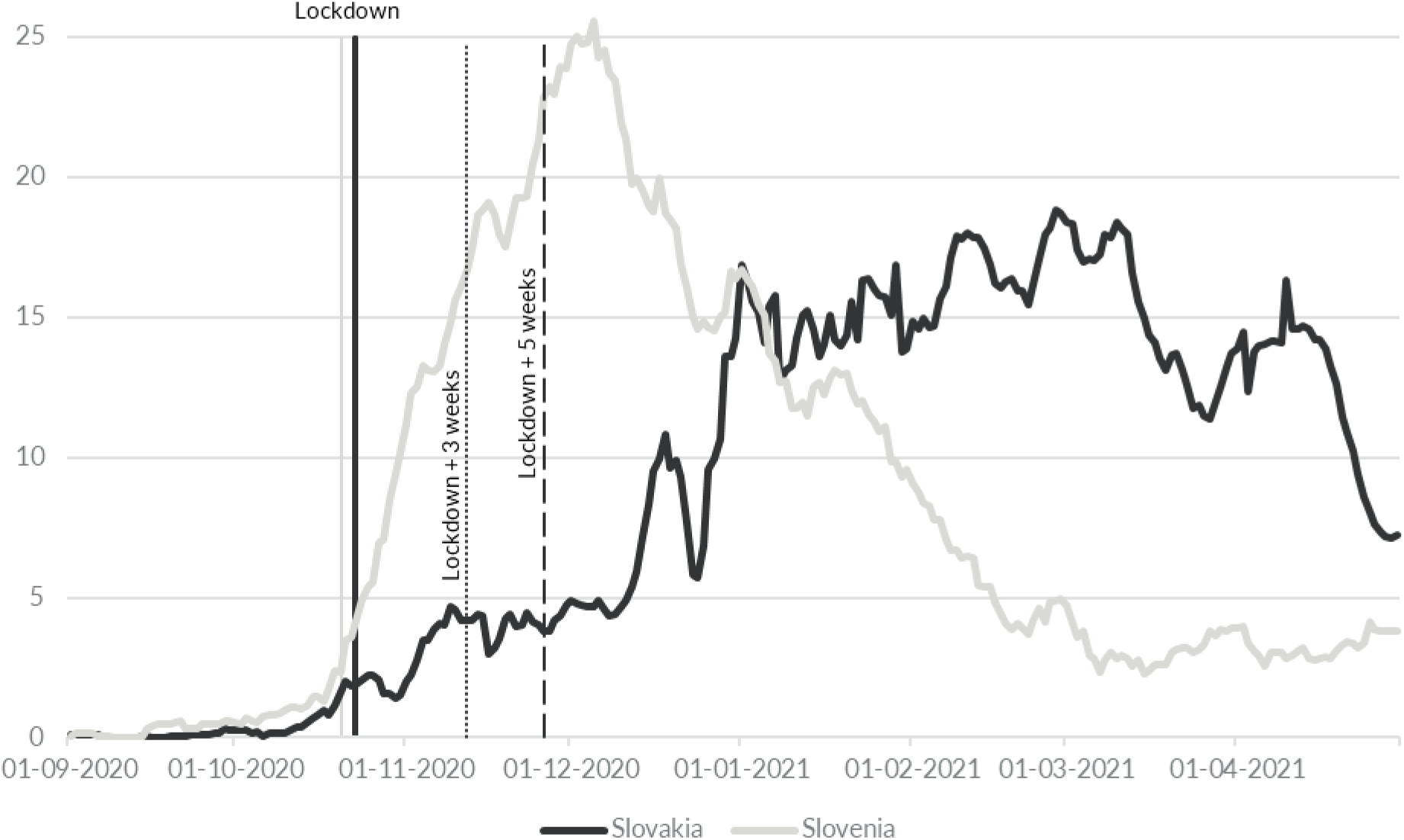
Lockdowns in Slovakia and Slovenia did not make mortality rates drop. Source: Our World in Data (2022) Note: Daily deaths are shown as seven days average. The black (Slovakia) and grey (Slovenia) vertical lines illustrates the day when the OxCGRT stringency index surpassed 70. The dotted and dashed lines show approximately three and five weeks after lockdowns were introduced. Source: Our World in Data and Folkhälsomyndigheten.

**Figure 17:**
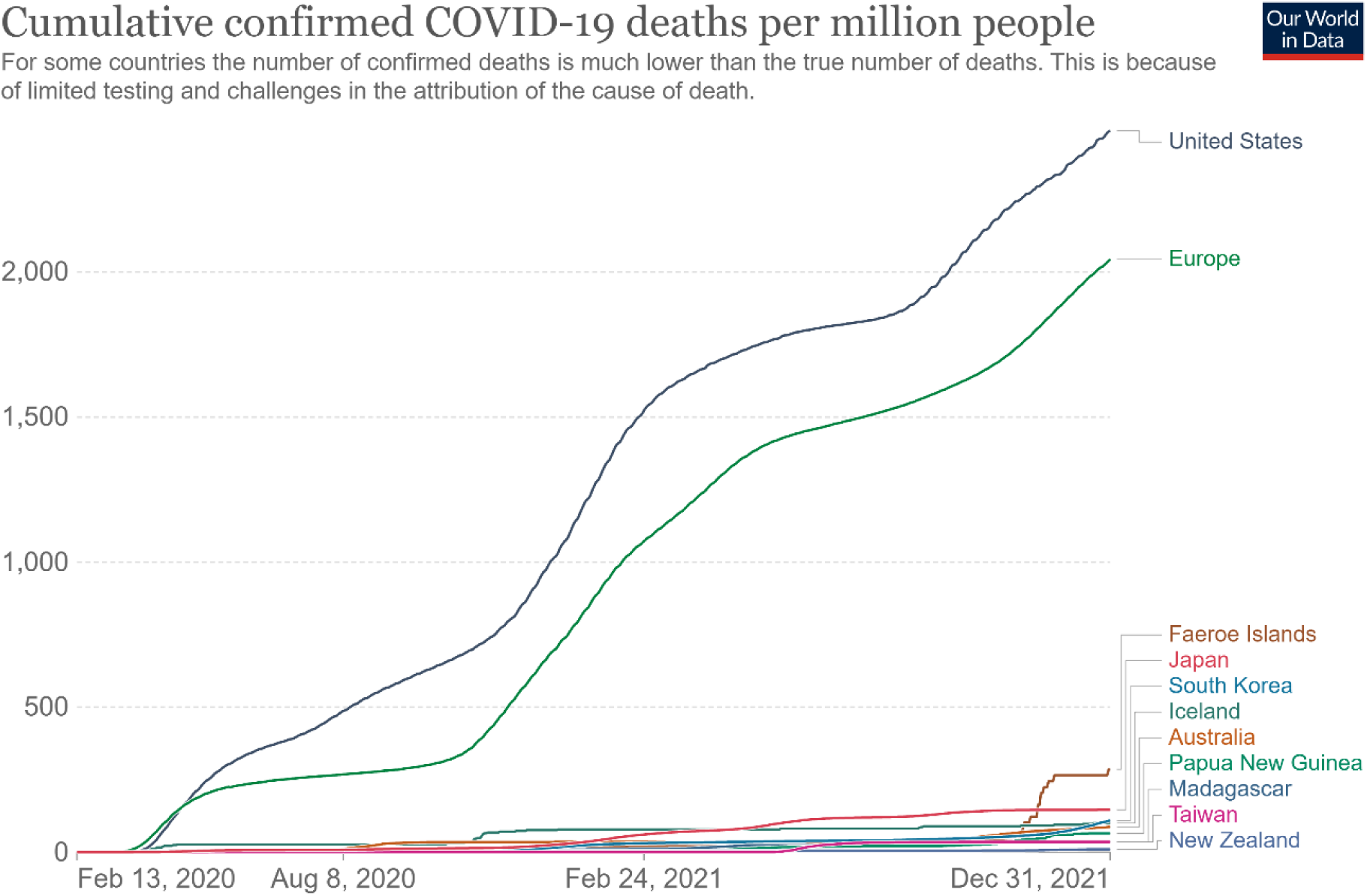
COVID-19 mortality rates have been relatively low in several island countries despite significant differences in their lockdown policies (2020-2021) Source: Our World in Data (2022). Note: South Korea is included as it is de facto an island.

#### 5.2.5 Other factors are more likely to explain the differences in COVID-19 mortality between countries than differences in lockdowns

But what else explains the differences between countries, if not differences in lockdown policies? Differences in population age and health, quality of the health sector, and the like are obvious factors. But several studies point at less obvious factors, such as culture, communication, and coincidences. For example, Frey et al. (2020) show that for the same policy stringency, countries with more obedient and collectivist cultural traits experienced larger declines in geographic mobility relative to their more individualistic counterparts. Using data from Germany, Laliotis and Minos (2020) show that the spread of COVID-19 and the resulting deaths in predominantly Catholic regions with stronger social and family ties were much higher when compared to non-Catholic ones at the local NUTS 3 level.^108^ Albæk (2021) notes that trust in others seems to be an important factor, see Figure 18.^109^ And Bollyky et al. (2022) find that “measures of trust in the government and interpersonal trust, as well as less government corruption, had larger, statistically significant associations with lower standardized infection rates”. Thornton (2022) finds that “if all societies had trust in government at least as high as Denmark, which is in the 75th percentile, the world would have experienced 13% fewer infections. If social trust—trust in other people—reached the same level, the effect would be even larger, with 40% fewer infections globally.” Similar results are found in several other studies.

**Figure 18:**
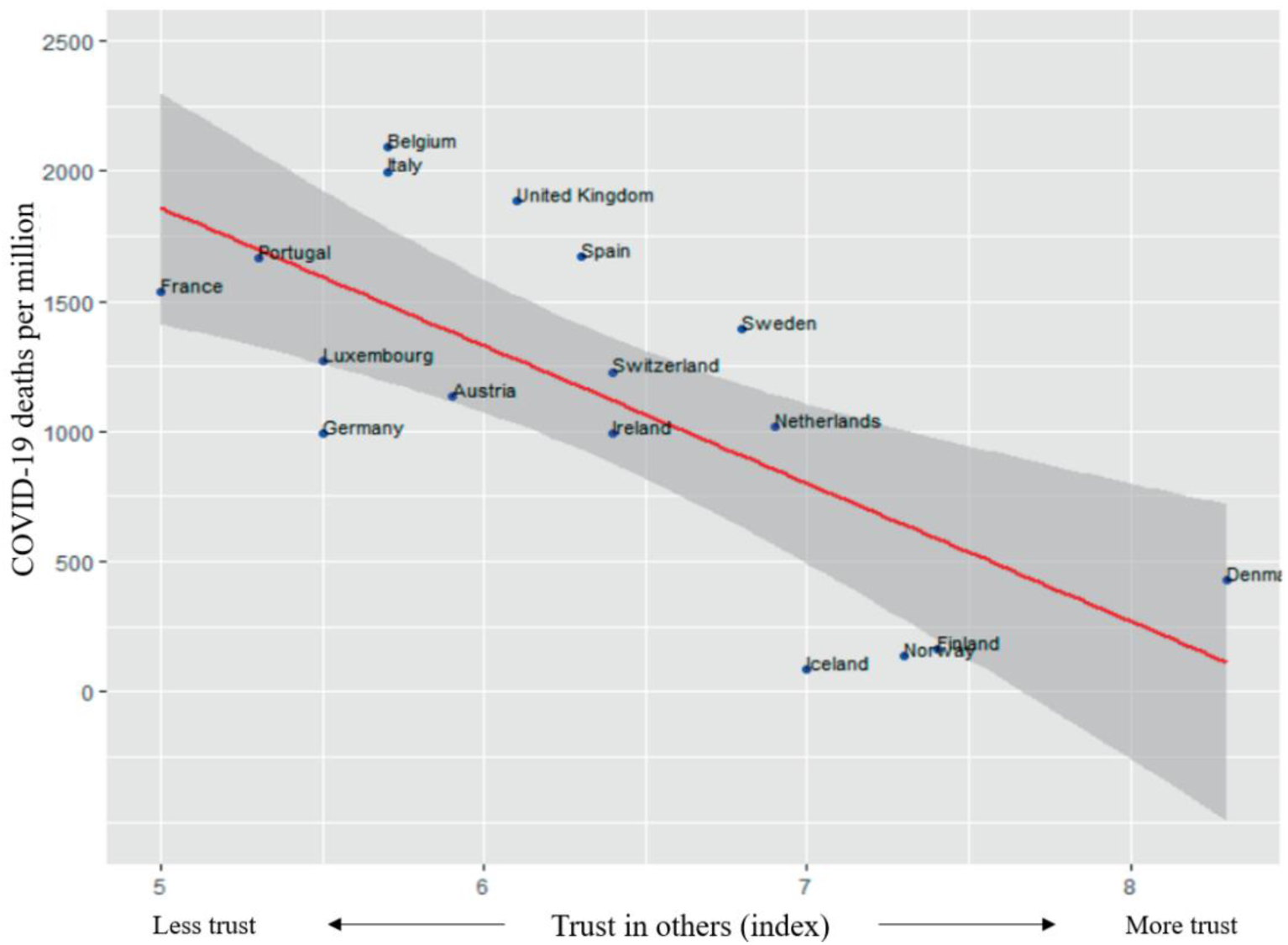
Countries with more trust in others experienced lower COVID-19 mortality rates. Source: Albæk (2021) Note: Axis titles have been translated from Danish to English.

Government communication may also have played a large role. Compared to its Scandinavian neighbors, the communication from Swedish health authorities was far more subdued and embraced the idea of public health vs. economic trade-offs. An illustration of the differences in perspective on the coming pandemic was visible on March 7^th^, 2020, when the Danish final in the European Song Contest – based on national health authorities’ strong recommendations – was held *without* audience in Denmark but – again based on national health authorities’ recommendations – *with* audience in Sweden.^110^

This clearly illustrates differences in the health authorities’ assessment of the risk, and this difference may explain why Helsingen, Refsum, et al. (2020) found, based on questionnaire data collected from mid-March to mid-April, 2020, that even though the daily COVID-19 mortality rate was more than four times higher in Sweden than in Norway, Swedes were less likely than Norwegians to not meet with friends (55% vs. 87%), avoid public transportation (72% vs. 82%), and stay home during spare time (71% vs. 87%). That is, despite a more severe pandemic, Swedes were less affected in their daily activities (legal in both countries) than Norwegians.

Many other factors may be relevant, and we should not underestimate the importance of coincidences. An interesting example illustrating this point is found in Arnarson (2021) and Björk et al. (2021), who show that areas in Europe where the winter holiday was relatively late (in week 9 or 10 rather than week 6, 7 or 8) were hit especially hard by COVID-19 during the first wave because the virus outbreak in the Alps could spread to those areas with ski tourists. Arnarson (2021) shows that the effect persisted in later waves. The importance of the timing of the winter holiday is illustrated in Figure 19 borrowed from Andersson (2022), which illustrate 1) excess mortality in Swedish regions (län) where the winter holiday was in week 9, 2) excess mortality in Swedish regions where the winter holiday was in other weeks, and 3) excess mortality in other Nordic countries. The figure illustrates how excess deaths in Sweden in the spring of 2020 were primarily driven by regions with winter holidays in week 9, while excess mortality in other regions were comparable to the excess mortality in other Nordic countries.

**Figure 19:**
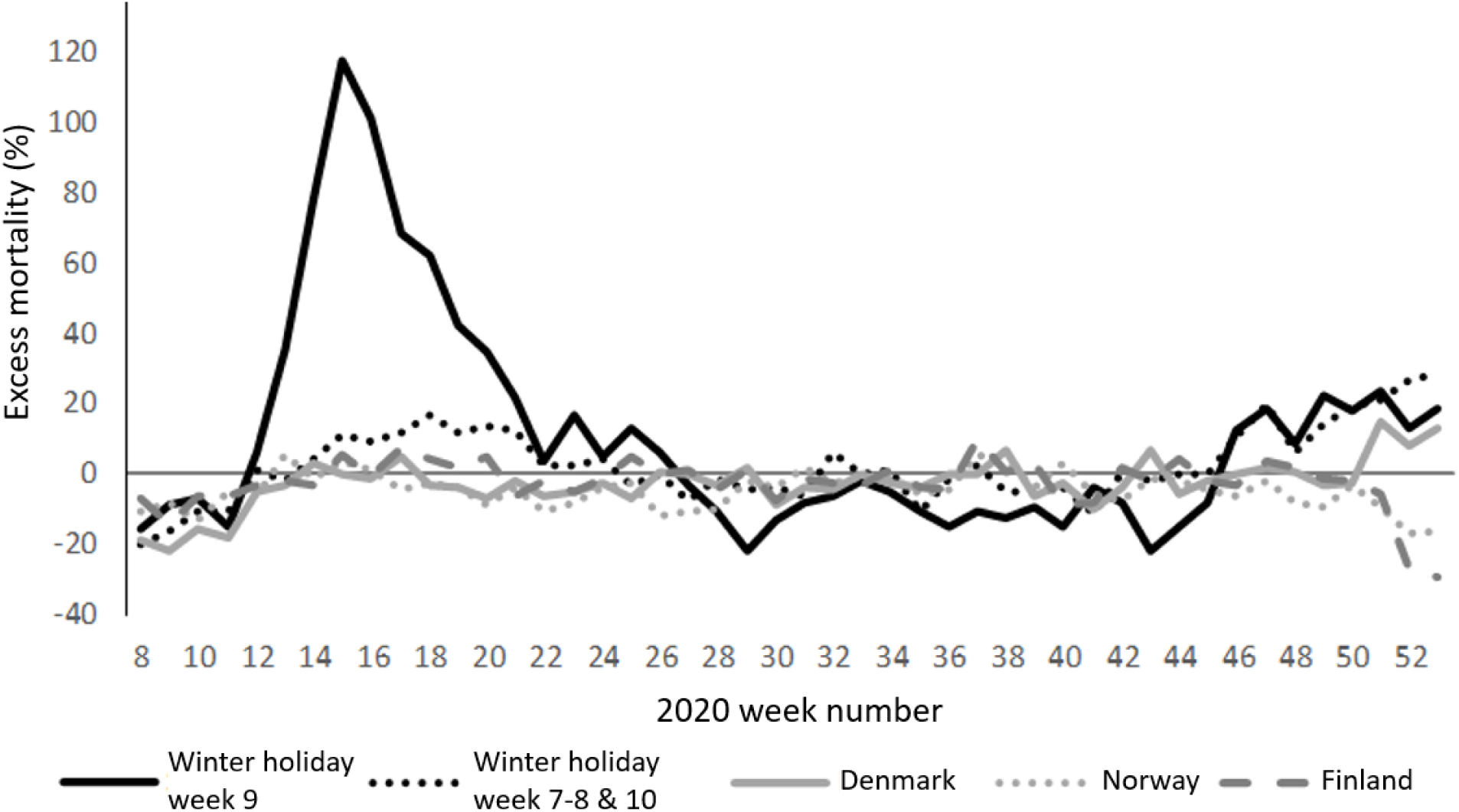
The excess mortality in Sweden in the spring of 2020 emerged primarily in regions with winter holidays in week 9, when ski tourists were unknowingly exposed to a COVID-19 virus outbreak in the Alps. Source: Andersson (2022) Note: Axis titles and legends have been translated from Swedish to English.

Also, Sweden had more frail elders due to very mild flu seasons in 2018/19 and 2019/20 as well as very few deaths during the 2019 summer compared to earlier years and compared to other Nordic countries (see Herby (2020)) which affected mortality in Sweden (see Juul et al. (2022); Zahran et al. (2022)). Had the winter holiday in Sweden been in week 7 or week 8 as in Denmark and had mortality rates in Sweden in 2018/19 and 2019/20 been comparable to mortality rates in Denmark, the Swedish COVID-19 situation could have turned out very differently.^111^

### 5.3 Policy implications

In the early stages of a pandemic, before the arrival of vaccines and new treatments, a society can respond in two ways: mandated behavioral changes or voluntary behavioral changes. Our study finds little to no positive health effects of mandated behavioral changes (lockdowns). This should draw our focus to the role of voluntary behavioral changes. Here, more research is needed to determine how voluntary behavioral changes can be supported. But, it should be clear that one important role for government authorities is to provide information etc. so that citizens can voluntarily respond to the pandemic in a way that mitigates their exposure.^112^

Finally, allow us to broaden our perspective after presenting our meta-analysis that focuses on the following question: “What does the evidence tell us about the effects of lockdowns on mortality?” We provide a firm answer to this question: Our study finds that lockdowns had little to no effect in reducing COVID-19 mortality.

The use of lockdowns is a unique feature of the COVID-19 pandemic. Lockdowns have not been used to such a large extent during any of the pandemics of the past century. However, lockdowns during the initial phase of the COVID-19 pandemic have had devastating effects.^113^ They have contributed to reducing economic activity, raising unemployment, reducing schooling^114^, causing political unrest^115^, contributing to domestic violence, loss of life quality^116^, and the undermining of liberal democracy. These costs to society must be compared to the benefits of lockdowns, which our meta-analysis has shown are little to none.^117^

Such a standard benefit-cost calculation leads to a strong conclusion: until future research based on credible empirical evidence can prove that lockdowns have large and significant reductions in mortality, lockdowns should be rejected out of hand as a pandemic policy instrument.

## Data Availability

All data produced in the present study are available upon reasonable request to the authors

## Appendix I. Supplementary information

### Excluded studies

Below is a list will the studies excluded during the eligibility phase of our identification process and a short description of our basis for excluding the study.

**Table 18:** Studies excluded in the meta-study identification process.

| 1. Study (Author & title) | 2. Reason for exclusion |
| --- | --- |
| Alemán et al. (2020); "Evaluating the effectiveness of policies against a pandemic" | Too few observations |
| Alshammari et al. (2021); "Are countries' precautionary actions against COVID-19 effective? An assessment study of 175 countries worldwide" | Is purely descriptive |
| Amuedo-Dorantes et al. (2020); "Timing is Everything when Fighting a Pandemic: COVID-19 Mortality in Spain" | Duplicate |
| Amuedo-Dorantes et al. (2021); "Early adoption of non-pharmaceutical interventions and COVID-19 mortality" | Only looks at timing |
| Amuedo-Dorantes et al. (2021); "Timing of social distancing policies and COVID-19 mortality: county-level evidence from the U.S." | Only looks at timing |
| Amuedo-Dorantes, Kaushal and Muchow (2020); "Is the Cure Worse than the Disease? County-Level Evidence from the COVID-19 Pandemic in the US" | Duplicate |
| Amuedo-Dorantes, Kaushal and Muchow (2021); "Timing of social distancing policies and COVID-19 mortality: county-level evidence from the US" | Only looks at timing |
| Aparicio and Grossbard (2021); "Are Covid Fatalities in the US Higher than in the EU, and If so, Why?" | Not difference-in- |
| Arruda et al. (2021); "ASSESSING THE IMPACT OF SOCIAL DISTANCING ON COVID-19 CASES AND DEATHS IN BRAZIL: AN INSTRUMENTED | Social distancing (not |
| Auger et al. (2020); "Association between statewide school closure and COVID-19 incidence and mortality in the US" | Uses a time series |
| Bakolis et al. (2021); "Changes in daily mental health service use and mortality at the commencement and lifting of COVID-19 'lockdown' policy in 10 | Uses a time series |
| Bardey, Fernández and Gravel (2021); "Coronavirus and social distancing: do non-pharmaceutical-interventions work (at least) in the short run?" | Only looks at timing |
| Berardi et. Al. (2020); "The COVID-19 pandemic in Italy: policy and technology impact on health and non-health outcomes" | Too few observations |
| Bhalla (2020); "Lockdowns and Closures vs COVID-19: COVID Wins" | Uses modelling |
| Bharati and Fakir (2020); "Pandemic Catch-22: How effective are mobility restrictions in halting the spread of COVID-19 in developing countries" | Duplicate |
| Björk et al. (2021); "Impact of winter holiday and government responses on mortality in Europe during the first wave of the COVID-19 pandemic" | Only looks at timing |
| Bongaerts et al. (2021); "Closed for business: The mortality impact of business closures during the Covid-19 pandemic" | Too few observations |
| Bongaerts et al. (2021); "Closed for business: The mortality impact of business closures during the Covid-19 pandemic" | Duplicate |
| Bongaerts, Mazzola and Wagner (2020); "Closed for business" | Duplicate |
| Born, Dietrich and Müller (2021); "The lockdown effect: A counterfactual for Sweden" | Synthetic control study |
| Born, Dietrich and Müller (2021); "The lockdown effect: A counterfactual for Sweden" | Duplicate |
| Borri et al. (2020); "The "Great Lockdown": Inactive workers and mortality by Covid-19" | Too few observations |
| Bushman et al. (2020); "Effectiveness and compliance to social distancing during COVID-19" | Social distancing (not |
| Canatay et al. (2021); "Critical country-level determinants of death rate during Covid-19 pandemic" | Not difference-in- |
| Caselli et al. (2020); "From the lockdown to the new normal: An analysis of the limitations to individual mobility in Italy following the Covid-19 crisis" | Do not look at mortality |
| Castaneda and Saygili (2020); "The effect of shelter-in-place orders on social distancing and the spread of the COVID-19 pandemic: a study of Texas" | Uses a time series |
| Cerqueti et al. (2021); "The sooner the better: lives saved by the lockdown during the COVID-19 outbreak. The case of Italy" | Synthetic control study |
| Chaudhry et al. (2020); "A country level analysis measuring the impact of government actions, country preparedness and socioeconomic factors on | Not difference-in- |
| Chernozhukov, Kasahara and Schrimpf (2021); "Mask mandates and other lockdown policies reduced the spread of COVID-19 in the US" | Duplicate |
| Chin et al. (2020); "Effects of non-pharmaceutical interventions on COVID-19: A Tale of Three Models" | Uses modelling |
| Cho (2020); "Quantifying the impact of nonpharmaceutical interventions during the COVID-19 outbreak: The case of Sweden" | Synthetic control study |
| Ciminelli and Garcia-Mandico (2020); "When and how do business shutdowns work? Evidence from Italy's first COVID-19 wave" | Too few observations |
| Ciminelli and Garcia-Mandico (2021); "Business shutdowns and covid-19 mortality" | Duplicate |
| Coccia (2020); "The effect of lockdown on public health and economic system: findings from first wave of the COVID-19 pandemic for designing | Only looks at timing |
| Coccia (2021); "Different effects of lockdown on public health and economy of countries: Results from first wave of the COVID-19 pandemic" | Too few observations |
| Canyon and Thomsen (2021); "COVID-19 in Scandinavia" | Synthetic control study |
| Canyon et al. (2020); "Lockdowns and COVID-19 deaths in Scandinavia" | Too few observations |
| Dave et al. (2020); "Did the Wisconsin Supreme Court restart a COVID-19 epidemic? Evidence from a natural experiment" | Synthetic control study |
| Delis, Iosifidi and Tasiou (2021); "Efficiency of government policy during the COVID-19 pandemic" | Do not look at mortality |
| Dey et al. (2021); "Lag time between state-level policy interventions and change points in COVID-19 outcomes in the United States" | Uses a time series |
| Dreher et al. (2021); "Policy interventions, social distancing, and SARS-CoV-2 transmission in the United States: a retrospective state-level analysis" | Do not look at mortality |
| Duchemin, Veber and Boussau (2020); "Bayesian investigation of SARS-CoV-2-related mortality in France" | Uses modelling |
| Fair et. Al. (2021); "Estimating COVID-19 cases and deaths prevented by non-pharmaceutical interventions in 2020-2021, and the impact of individual | Uses modelling |
| Filiis (2020); "The impact of government policies effectiveness on the officially reported deaths attributed to covid-19." | Student paper |
| Fountoulakis et al. (2020); "Factors determining different death rates because of the COVID-19 outbreak among countries" | Not difference-in- |
| Fowler et al. (2021); "Stay-at-home orders associate with subsequent decreases in COVID-19 cases and fatalities in the United States" | Duplicate |
| Friedson et al. (2020); "Did California's shelter-in-place order work? Early coronavirus-related public health effects" | Duplicate |
| Friedson et al. (2020); "Shelter-in-place orders and public health: evidence from California during the COVID-19 pandemic" | Synthetic control study |
| Fuss, Weizman and Tan (2020); "COVID19 pandemic: how effective are interventive control measures and is a complete lockdown justified? A | Do not look at mortality |
| Ghosh, Ghosh and Narymanchi (2020); "A Study on The Effectiveness of Lock-down Measures to Control The Spread of COVID-19" | Synthetic control study |
| Glogowsky et al. (2021); "How Effective Are Social Distancing Policies? Evidence on the Fight Against COVID-19" | Only looks at timing |
| Glogowsky, Hansen and Schächtele (2020); "How effective are social distancing policies? Evidence on the fight against COVID-19 from Germany" | Duplicate |
| Glogowsky, Hansen and Schächtele (2020); "How Effective Are Social Distancing Policies? Evidence on the Fight Against COVID-19 from Germany" | Duplicate |
| Gordon, Grafton and Steinshamn (2021); "Cross-country effects and policy responses to COVID-19 in 2020: The Nordic countries" | Do not look at mortality |
| Gordon, Grafton and Steinshamn (2021); "Statistical Analyses of the Public Health and Economic Performance of Nordic Countries in Response to the | Too few observations |
| Guo et al. (2020); "Social distancing interventions in the United States: An exploratory investigation of determinants and impacts" | Duplicate |
| Hale et al. (2020); "Global Assessment of the Relationship between Government Response Measures and COVID-19 Deaths" | Duplicate |
| Huber and Langen (2020); "The impact of response measures on COVID-19-related hospitalization and death rates in Germany and Switzerland" | Duplicate |
| Huber and Langen (2020); "Timing matters: the impact of response measures on COVID-19-related hospitalization and death rates in Germany and | Only looks at timing |
| Hunter et al. (2021); "Impact of non-pharmaceutical interventions against COVID-19 in Europe: A quasi-experimental non-equivalent group and time- | Not difference-in- |
| Jain et al. (2020); "A comparative analysis of COVID-19 mortality rate across the globe: An extensive analysis of the associated factors" | Do not look at mortality |
| Juraneck and Zoutman (2021); "The effect of non-pharmaceutical interventions on the demand for health care and mortality: evidence on COVID-19 in | Too few observations |
| Juraneck and Zoutman (2021); "The Effect of Non-Pharmaceutical Interventions on the Demand for Health Care and Mortality: Evidence on COVID-19 in | Duplicate |
| Juraneck and Zoutman (2021); "The effect of non-pharmaceutical interventions on the demand for health care and on mortality: evidence from COVID- | Duplicate |
| Kakpo and Nuhu (2020); "Effects of Social Distancing on COVID-19 Infections and Mortality in the US" | Social distancing (not |
| Kapitsinis (2021); "The underlying factors of excess mortality in 2020: a cross-country analysis of pre-pandemic healthcare conditions and strategies to | Not difference-in- |
| Kapoor and Ravi (2020); "Impact of national lockdown on COVID-19 deaths in select European countries and the US using a Changes-in-Changes | Too few observations |
| Khan et al. (2021); "Assessing the Impact of Policy Measures in Reducing the COVID-19 Pandemic: A Case Study of South Asia" | Too few observations |
| Khatiawada and Chalise (2020); "Evaluating the efficiency of the Swedish government policies to control the spread of Covid-19." | Student paper |
| Korevaar et al. (2020); "Quantifying the impact of US state non-pharmaceutical interventions on COVID-19 transmission" | Do not look at mortality |
| Kumar et. Al. (2020); "Prevention-Versus Promotion-Focus Regulatory Efforts on the Disease Incidence and Mortality of COVID-19: A Multinational | Do not look at mortality |
| Langeland et al. (2021); "The Effect of State Level COVID-19 Stay-at-Home Orders on Death Rates" | Not difference-in- |
| Le et al. (2020); "Impact of government-imposed social distancing measures on COVID-19 morbidity and mortality around the world" | Uses a time series |
| Liang et al. (2020); "Covid-19 mortality is negatively associated with test number and government effectiveness" | Not effect of lockdowns |
| Mader and Rütternauer (2021); "The effects of non-pharmaceutical interventions on COVID-19-related mortality: A generalized synthetic control | Synthetic control study |
| Matzinger and Skinner (2020); "Strong impact of closing schools, closing bars and wearing masks during the Covid-19 pandemic: results from a simple | Uses modelling |
| Mccafferty and Ashley (2020); "Covid-19 Social Distancing Interventions by State Mandate and their Correlation to Mortality in the United States" | Duplicate |
| Medline et al. (2020); "Evaluating the impact of stay-at-home orders on the time to reach the peak burden of Covid-19 cases and deaths: does timing | Only looks at timing |
| Mu et al. (2020); "Effect of social distancing interventions on the spread of COVID-19 in the state of Vermont" | Uses modelling |
| Nakamura (2020); "The Impact of Rapid State Policy Response on Cumulative Deaths Caused by COVID-19" | Student paper |
| Neidhöfer and Neidhöfer (2020); "The effectiveness of school closures and other pre-lockdown COVID-19 mitigation strategies in Argentina, Italy, and | Synthetic control study |
| Oliveira (2020); "Does Staying at Home Save Lives? An Estimation of the Impacts of Social Isolation in the Registered Cases and Deaths by COVID-19 in | Social distancing (not |
| Palladina et al. (2020); "Effect of Implementation of the Lockdown on the Number of COVID-19 Deaths in Four European Countries" | Uses a time series |
| Palladina et al. (2020); "Effect of timing of implementation of the lockdown on the number of deaths for COVID-19 in four European countries" | Duplicate |
| Palladino et al. (2020); "Excess deaths and hospital admissions for COVID-19 due to a late implementation of the lockdown in Italy" | Uses a time series |
| Pan et al. (2021); "Heterogeneity in the Effectiveness of Non-pharmaceutical Interventions During the First SARS-CoV2 Wave in the United States" | Duplicate |
| Peixoto et al. (2020); "Rapid assessment of the impact of lockdown on the COVID-19 epidemic in Portugal" | Uses modelling |
| Piovani et. Al. (2021); "Effect of early application of social distancing interventions on COVID-19 mortality over the first pandemic wave: An analysis of | Only looks at timing |
| Porto et al. (2022); "Lockdown, essential sectors, and Covid-19: Lessons from Italy" | Too few observations |
| Reinbold (2021); "Effect of fall 2020 K-12 instruction types on CoViD-19 cases, hospital admissions, and deaths in Illinois counties" | Synthetic control study |
| Renne, Roussellet and Schwenkler (2020); "Preventing COVID-19 Fatalities: State versus Federal Policies" | Uses modelling |
| Shanmugam et al. (2021); "A report card on prevention efforts of covid-19 deaths in US" | Not difference-in- |
| Siedner et al. (2020); "Social distancing to slow the US COVID-19 epidemic: Longitudinal pretest–posttest comparison group study" | Duplicate |
| Siedner et al. (2020); "Social distancing to slow the US COVID-19 epidemic: Longitudinal pretest–posttest comparison group study" | Uses a time series |
| Silva, Filho and Fernandes (2020); "The effect of lockdown on the COVID-19 epidemic in Brazil: evidence from an interrupted time series design" | Uses a time series |
| Stamam et al. (2020); "IMPACT OF LOCKDOWN MEASURE ON COVID-19 INCIDENCE AND MORTALITY IN THE TOP 31 COUNTRIES OF THE | Uses a time series |
| Steinegger et al. (2021); "Retrospective study of the first wave of COVID-19 in Spain: analysis of counterfactual scenarios" | Only looks at timing |
| Stephens et al. (2020); "Does the timing of government COVID-19 policy interventions matter? Policy analysis of an original database." | Only looks at timing |
| Stockenhuber (2020); "Did We Respond Quickly Enough? How Policy-Implementation Speed in Response to COVID-19 Affects the Number of Fatal | Not difference-in- |
| Supino et al. (2020); "The effects of containment measures in the Italian outbreak of COVID-19" | Uses a time series |
| Thayer et al. (2021); "An interrupted time series analysis of the lockdown policies in India: a national-level analysis of COVID-19 incidence." | Uses a time series |
| Timelli and Girardi (2021); "Effect of timing of implementation of containment measures on Covid-19 epidemic. The case of the first wave in Italy" | Only looks at timing |
| Toya and Skidmore (2021); "A Cross-Country Analysis of the Determinants of Covid-19 Fatalities" | Not difference-in- |
| Trivedi and Das (2020); "Effect of the timing of stay-at-home orders on COVID-19 infections in the United States of America" | Only looks at timing |
| Tsai et al. (2021); "Coronavirus Disease 2019 (COVID-19) Transmission in the United States Before Versus After Relaxation of Statewide Social | Uses a time series |
| Umer and Khan (2020); "Evaluating the Effectiveness of Regional Lockdown Policies in the Containment of Covid-19: Evidence from Pakistan" | Too few observations |
| VoPham et al. (2020); "Effect of social distancing on COVID-19 incidence and mortality in the US" | Do not look at mortality |
| Wu and Wu (2020); "Stay-at-home and face mask policies intentions inconsistent with incidence and fatality during US COVID-19 pandemic" | Too few observations |
| Xu et al. (2020); "Associations of Stay-at-Home Order and Face-Masking Recommendation with Trends in Daily New Cases and Deaths of Laboratory- | Do not look at mortality |
| Yehya et al. (2021); "Statewide Interventions and Coronavirus Disease 2019 Mortality in the United States: An Observational Study" | Is purely descriptive |
| Yehya, Venkataramani and Harhay (2020); "Statewide Interventions and Coronavirus Disease 2019 Mortality in the United States: An Observational | Only looks at timing |
| Yilmazkuday (2021); "Stay-at-home works to fight against COVID-19: international evidence from Google mobility data" | Social distancing (not |
| Ylli et al. (2020); "The lower COVID-19 related mortality and incidence rates in Eastern European countries are associated with delayed start of | Not effect of lockdowns |
| Zimmerman and Anderson (2021); "Association of the timing of school closings and behavioral changes with the evolution of the coronavirus disease | Uses a time series |

### Interpretation of estimates and conversion to standardized estimates

In Table 19, we describe for each study how we interpret their results and convert the estimates to our standardized estimate. For studies not included in the meta-analysis, we describe why. Standard errors are converted such that the t-value, calculated based on standardized estimates and standard errors, is unchanged. When confidence intervals are reported rather than standard errors, we calculate standard errors using t-distribution with ∞ degrees of freedom (i.e., 1.96 for 95% confidence interval).

**Table 19:**
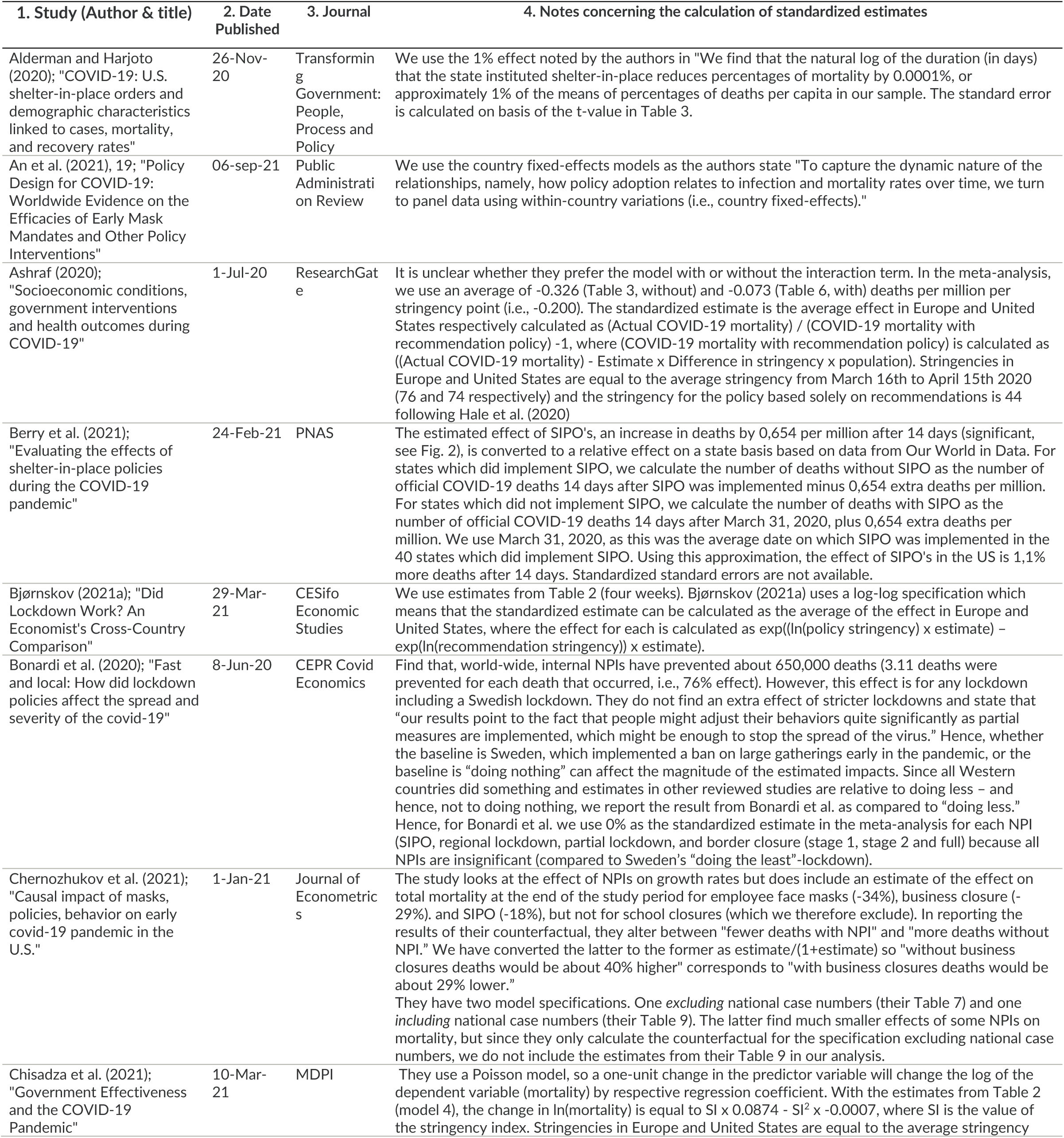
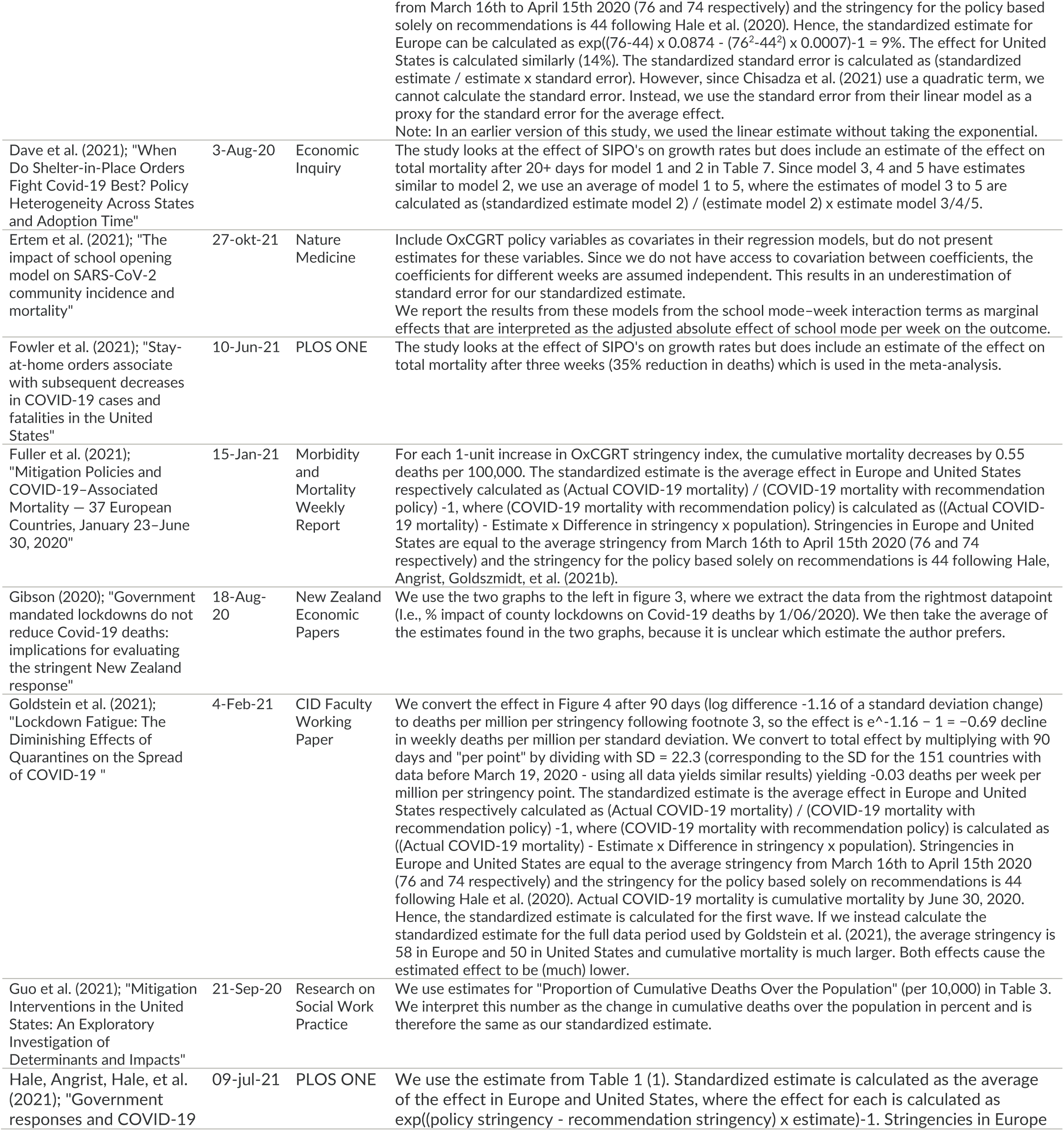
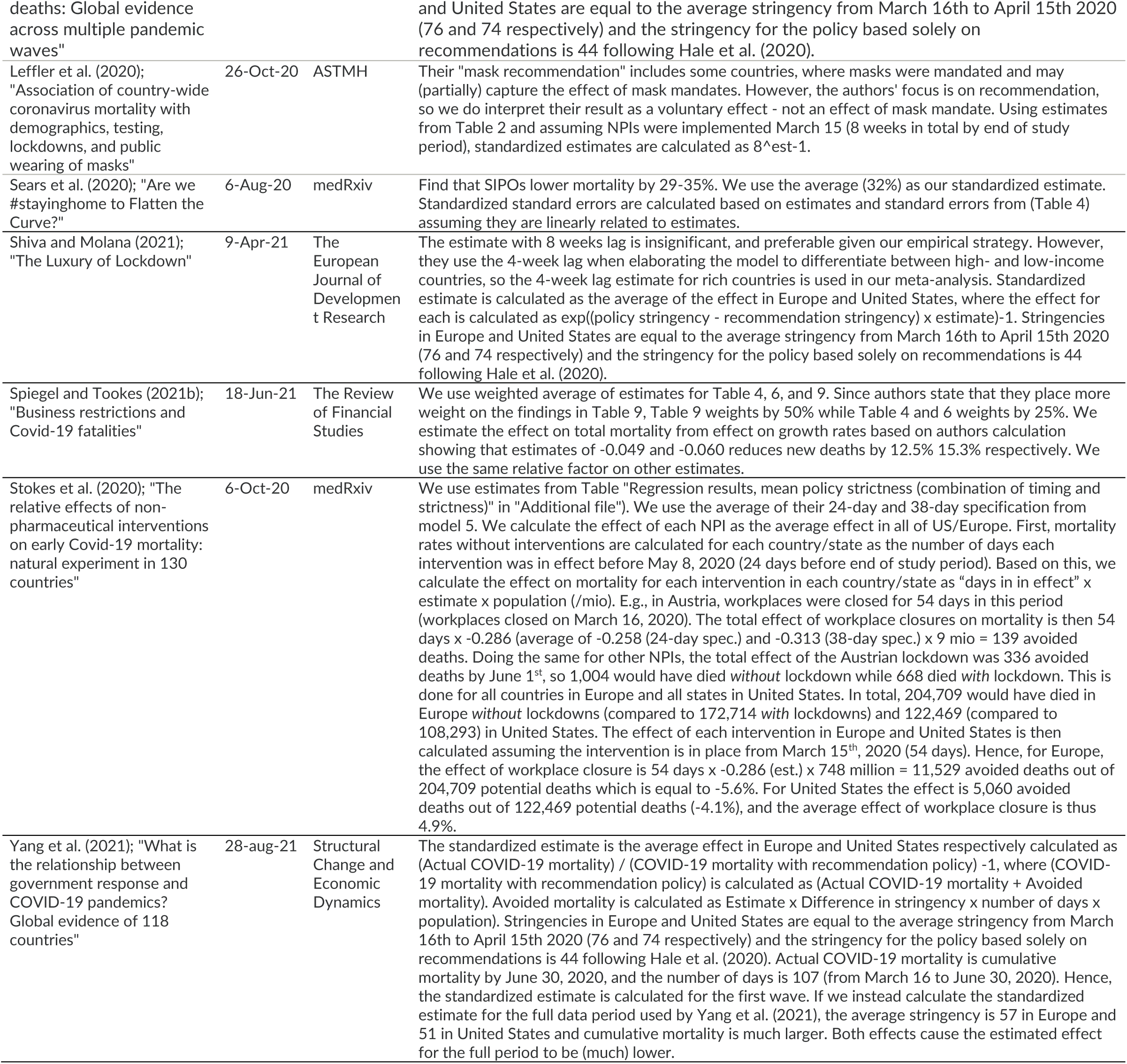
Notes concerning the standardization of results of the studies included in the meta-analysis.

## Appendix II: The Anatomy of the Negative Spin-Meisters

The Johns Hopkins Institute for Applied Economics, Global Health, and the Study of Business Enterprise, which one of us (Hanke) founded and co-directs, published “A Systematic Literature Review and Meta-Analysis of the Effects of Lockdowns on COVID-19 Mortality” in its *Studies in Applied Economics* working paper series on January 21, 2022. The working paper’s findings – that lockdowns had little to no public health effects measured by mortality – and its policy conclusions – that lockdown policies are ill-founded and should be rejected out of hand – attracted considerable attention in the media, in the White House and halls of the U.S. Congress, among public health experts, and within the chattering classes around the world.

But, it was the strong endorsement of Dr. Marty Makary, a distinguished Professor of Medicine at the Johns Hopkins School of Medicine, during his February 2 appearance on *Tucker Carlson Tonight* that set off a media firestorm. Indeed, on February 3, the Science Media Centre in London issued a press release, Science Media Centre (2022), with statements by Prof. Neil Ferguson, Dr. Seth Flaxman, Prof. Samir Bhatt – all affiliated with Imperial College London and authors of two of the studies (Ferguson et al. (2020) and Flaxman et al. (2020)), we implicitly criticized – and Prof. David Paton (Nottingham University Business School). The release contained several criticisms of our working paper from the Imperial College team, many of which were unscientific^118^ or clearly flawed.^119,120^ Those were authored by Prof. Ferguson, Dr. Flaxman, and Prof. Bhatt. The press release also contained positive comments by Prof. Paton.

The accompanying Figure 20 denotes five of the most scientific criticisms raised in Science Media Centre (2022) as well as five criticisms raised by follow-up “fact checkers”. Although we believe that the criticisms are with little or no merit, we are not going to engage in a critique of them in this Appendix, as the relevant criticisms are dealt with, either directly or indirectly, in the text of this working paper.^121^ Our purpose is to illustrate how biased and politicized the media was when it came to its reportage of our working paper.

**Figure 20:**
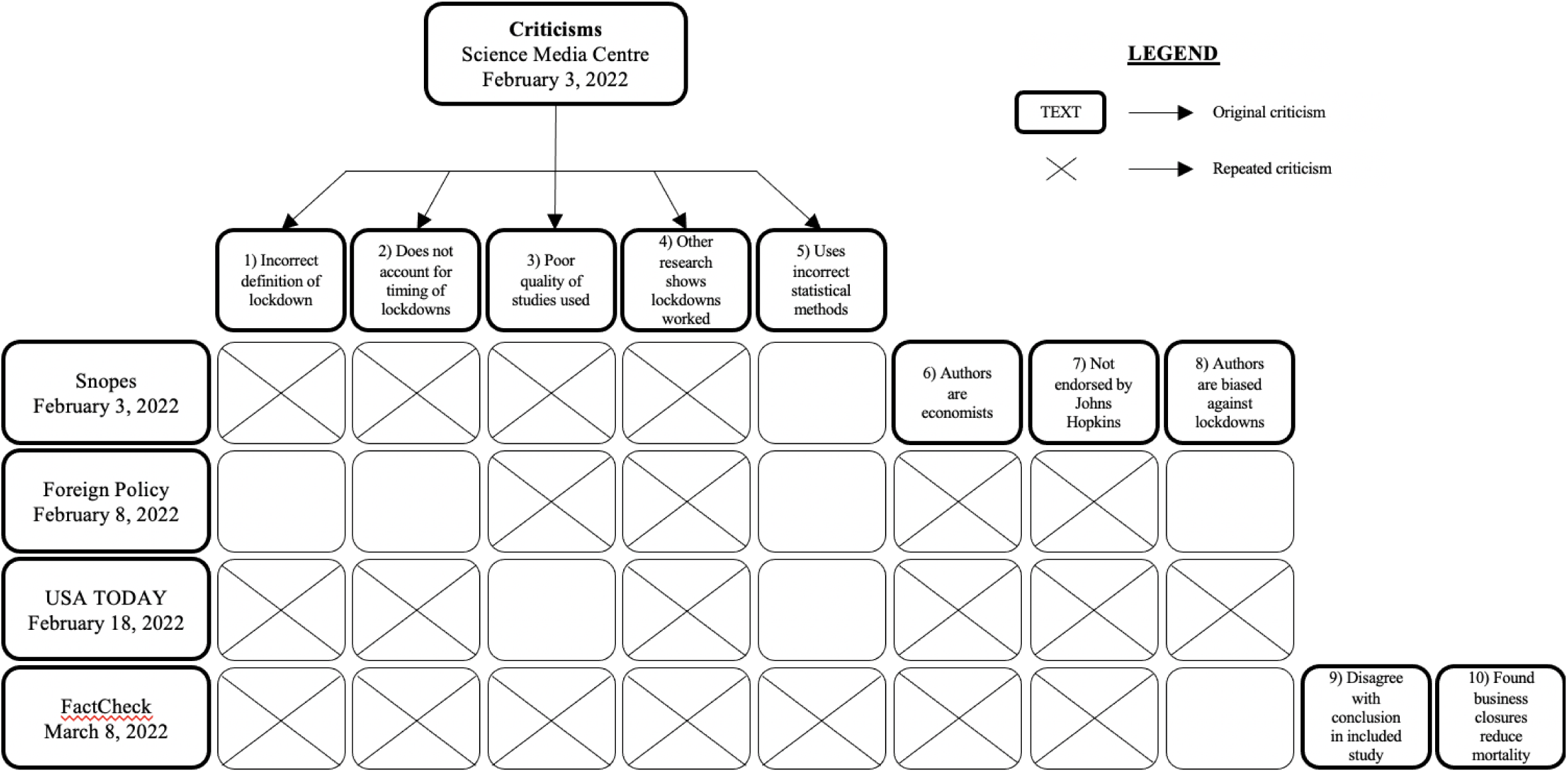
Flow chart for criticisms raised in Science Media Centre.

*Snopes*, which advertises itself as a “fact-checking” website, was quick to jump on the bandwagon. A few hours after the Science Media Centre press release, *Snopes* published a report that contained eight criticisms, Evon (2022). Of those, five were lifted from the Science Media Centre press release. And three new but irrelevant “criticisms” were added, see Figure 20 and footnote 121.

On February 8, *Foreign Policy* published a critique, Garrett (2022). It copied three of the Science Media Centre’s criticisms and two of the new ones added by *Snopes*. In short, *Foreign Policy*’s article presented nothing new.

Then, *USA Today* entered the picture on February 18, Sadeghi (2022). The fact checkers at *USA Today* offered seven criticisms—four copied from the Science Media Centre and three from *Snopes*.

On March 8, *FactCheck* presented ten criticisms, Robertson (2022). They included six from the Science Media Centre (including all five that we identified in Figure 20), two from *Snopes*, and two new confused and misleading criticisms, see Figure 20 and footnote 121.

There was, of course, a great deal of reportage about our working paper that appeared in the early February-April 2022 period. This material was highly repetitive, echoing material presented on February 3 in either the Science Media Centre press release or the *Snopes* report. There were several similar reports, but to avoid repetition ourselves, we limit our review to the five reports contained in Figure 20.

The striking feature of the media flow is its unoriginality. Indeed, there is little evidence that the post-Science Media Centre press release authors even engaged in basic “primary” reading of our text, let alone any “critical” reading of the text. What is evident is what Martin Heidegger identified in his 1927 magnum opus *Being and Time* as “idle talk” (“Gerede”). This is inauthentic discourse, simply adopting and circulating others’ opinions about something without ever engaging in even reading a primary text.

If that was not bad enough, the post-Science Media Centre critiques are clearly biased as they all exclude any mention of Prof. Paton’s favorable appraisals of our working paper. For example, Prof. Paton made the following four points in Science Media Centre (2022):

1. “Both parts of the paper (systematic review and the meta analysis) make a significant contribution to our understanding of lockdown effects.”
2. “Key to a systematic review like this are the sets of search & exclusion criteria. The paper is very transparent about this which is good. They focus on difference-in-difference empirical studies. i.e. they look at papers which compare the impact of an intervention on mortality by looking before & after, but relative to other areas which did not have the intervention. As a result, modelling studies (like the well-known Flaxman Nature paper) are excluded. That is not controversial.”
3. “[The result] is pretty consistent with other, non-systematic reviews (e.g. Herby & Allen) which is reassuring. It is also consistent with the (few) studies which look at the impact on overall excess mortality.”
4. “More marginal in my view is their exclusion of synthetic control method (SCM) papers. Some of these papers find a significant impact of NPIs on mortality so including them might have led to somewhat higher average mortality effects. The paper gives a robust defence of their exclusion, but I think you would get people on both sides of that debate.”

As illustrated in Figure 21, none of these positive comments saw the light of day in the “fact checking” that followed the Science Media Centre’s press release on February 3. That’s because the new cottage industry called “fact checking” has arguably become highly politicized. As a result, there is not much fact checking, but rather opinions about whether the so-called fact checkers agreed or disagreed with the policy implications or conclusions of what they are supposed to be fact checking. So, for the most part, fact checkers were not engaged in fact checking, but were engaged in publishing opinion and narrative. By hiding behind the shroud of “facts” and “fact-checking,” they have attempted to cast doubt, in our case, via innuendo.

**Figure 21:**
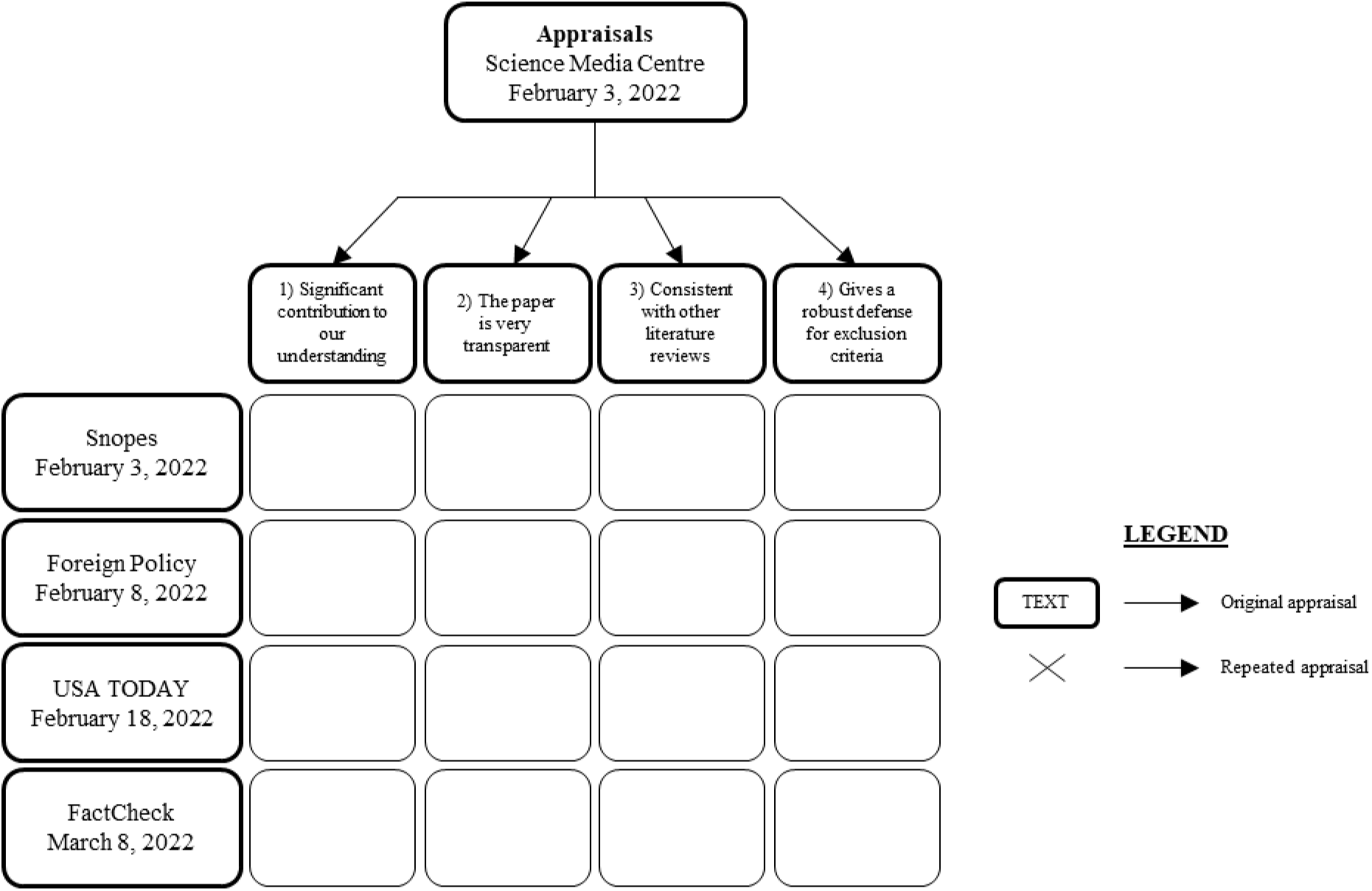
Flow chart for appraisals raised in Science Media Centre.

In closing this Appendix, we would like to indicate that we received extensive private reviews and comments on our working paper. After all, that’s the purpose of publishing working papers. Our professional correspondents did engage in a serious primary reading of our working paper and made many useful comments and suggestions. Most of their names appear in our thank-you note following the abstract.

We engaged in a thorough review and revision of our January 21, 2022 working paper. We can report that one error of commission was found in the original. It was not detected by any fact checkers or by those we corresponded with, but by us. The error was a computational error that involved logarithms. It was “small” and did not materially affect our results.

## List of acronyms

COVID-19: Coronavirus Disease 2019
NPI: Non-pharmaceutical intervention
OxCGRT: Oxford COVID-19 Government Response Tracker
PHEIC: Public Health Emergency of International Concern
PRISMA: Preferred Reporting Items for Systematic Reviews and Meta-Analyses
PWA: Precision-weighted average
R_t_: The effective reproductive number. This is the expected number of new infections caused by an infectious individual in a population at time *t*.
SE: Standard errors
SIPO: Shelter-in-place order
SIR: Susceptible-Infected-Recovered

## Footnotes

* We have received helpful comments from Douglas Allen, Torben M. Andersen, Fredrik N. G. Andersson, Andreas Bergh, Jonas Björk, Christian Bjørnskov, Joakim Book, Gunnar Brådvik, Kristoffer Torbjørn Bæk, Dave Campbell, Bernard Casey, Kevin Dowd, Ulf Gerdtham, Nicholas Hanlon, Caleb Hofmann, Olga B. Jonas, Daniel B. Klein, Fredrik Charpentier Ljungqvist, Christian Heebøl-Nielsen, Martin Paldam, Jonas Ranstam, Spencer Ryan, John Strezewski, Roger Svensson, Ulf Persson, Anders Waldenström, and Joakim Westerlund. We also thank several proofreaders who reviewed the first version of this study and helped us find ways in which we could clarify our methodology and facilitate the understanding of our results. We thank Line Andersen, Troels Sabroe Ebbesen, and Anders Lund Mortensen for excellent research assistance. Needless to say, the usual disclaimer holds: All remaining errors are our own.

1 See https://dictionary.cambridge.org/dictionary/english/lockdown and https://ig.ft.com/coronavirus-lockdowns/ for two different examples of how the term “lockdown” is defined and used.

2 For example, we will say that the government of Country A introduced the *non-pharmaceutical interventions* of school closures and shelter-in-place-orders as part of the country’s *lockdown*.

3 The Oxford Covid-19 Government Response Tracker (OxCGRT) collects systematic information on policy measures that governments have taken to tackle COVID-19. The different policy responses are tracked since 1 January 2020, cover more than 180 countries, and are coded into 23 indicators, such as school closures, travel restrictions, vaccination policy. These policies are recorded on a scale to reflect the extent of government action, and scores are aggregated into a suite of policy indices.

4 With R_0_ = 2.0 and trigger on 60, the number of COVID-19-deaths in Great Britain could be reduced to 5,600 deaths from 410,000 deaths (−99%) with a policy consisting of case isolation + home quarantine + social distancing + school/university closure, see Table 4 in Ferguson et al. (2020). R_0_ (the basic reproduction rate) is the expected number of cases directly generated by one case in a population where all individuals are susceptible to infection. The lowest effect of lockdowns modelled by Ferguson et al. (2020) was with R_0_ = 2.6, trigger on 200-400, and case isolation + home quarantine + social distancing. In this case, deaths were predicted to be reduced from 550.000 to 120.000 (−78%).

5 This perception was supported by statements from the study’s authors and Imperial College London. March 17, 2020, Imperial College tweeted that “without more action, the virus would have overwhelmed intensive care units“. And, Nial Ferguson is cited for saying that “we’re going to have to suppress this virus — frankly, indefinitely — until we have a vaccine.”, see https://www.nytimes.com/2020/03/16/us/coronavirus-fatality-rate-white-house.html.

6 For example, see https://www.theguardian.com/world/2020/mar/16/new-data-new-policy-why-uks-coronavirus-strategy-has-changed.

7 Woolhouse (2022), who in January 2020 was appointed to a SAGE sub-committee called the Scientific Pandemic Influenza Group on Modelling (SPI-M) in the United Kingdom, describes how the reaction was due to a misinterpretation of the report from Ferguson et al. (2020): “The report should have been a wake-up call that we needed to invest quickly and heavily in other ways to control novel coronavirus or – according to the model – we’d end up in lockdown. This implication was barely mentioned – lockdown was accepted as a necessity the first time it was proposed. When Report 9 [i.e. Ferguson et al. (2020)]was published the details of the scenarios modelled were quickly forgotten, as were any mentions of the assumptions, caveats and uncertainties of the analysis. Report 9 was condensed to the simple but misleading message that, if the government didn’t impose full lockdown immediately, over half a million people would die.”

8 Given Ferguson and his Imperial College team’s track record, it’s surprising that questions were not raised before jumping on a lockdown bandwagon, see Hanke and Dowd (2022).

9 In economics, an externality is an indirect cost or benefit to a third party that arises as an effect of another party’s (or parties’) activity.

10 We use “mortality” and “mortality rates” interchangeably to mean COVID-19 deaths as a percentage of population.

11 The protocol was first published June 23, 2021, and updated the last time October 28, 2021.

12 The government response search string used was: “non-pharmaceutical”, “nonpharmaceutical”, “NPI”, “NPIs”, “lockdown”, “social distancing orders”, “statewide interventions”, “distancing interventions”, “circuit breaker”, “containment measures”, “contact restrictions”, “social distancing measures”, “public health policies”, “mobility restrictions”, “covid-19 policies”, “corona policies”, “policy measures”.

13 The methodology search string used was: “fixed effects”, “panel data”, “difference-in-difference”, “diff-in-diff”, “synthetic control”, “counterfactual”, “counter factual”, “cross country”, “cross state”, “cross county”, “cross region”, “cross regional”, “cross municipality”, “country level”, “state level”, “county level”, “region level”, “regional level”, “municipality level”, “event study”.

14 If a potentially relevant paper from one of the 13 reviews (see eligibility criteria) did not show up in our search, we added relevant words to our search strings and ran the search again. The 13 reviews were: Allen (2021); Brodeur et al. (2021); Gupta et al. (2020); Herby (2021a); Johanna et al. (2020); Nussbaumer-Streit et al. (2020); Patel et al. (2020); Perra (2020); Poeschl and Larsen (2021); Pozo-Martin et al. (2020); Rezapour et al. (2021); Robinson (2021); Zhang et al. (2021).

15 This included studies with titles such as “COVID-19 outbreak and air pollution in Iran: A panel VAR analysis” and “Dynamic Structural Impact of the COVID-19 Outbreak on the Stock Market and the Exchange Rate: A Cross-country Analysis Among BRICS Nations.”

16 Professor Christian Bjørnskov of the University of Aarhus was particularly helpful in this process.

17 Analyses based on cases pose major problems, as testing strategies for COVID-19 infections vary enormously across countries (and even over time within a given country). In consequence, cross-country comparisons of cases are, at best, problematic. Although these problems exist with death tolls as well, they are far more limited. Also, while cases and death tolls are correlated, there may be adverse effects of lockdowns that are not captured by the number of cases. For example, an infected person who is isolated at home with family under a SIPO may infect family members with a higher viral load causing more severe illness. So even if a SIPO reduces the number of cases, it may theoretically increase the number of COVID-19-deaths. Adverse effects like this may explain why studies like Chernozhukov et al. (2021) find that SIPO reduces the number of cases but have no significant effect on the number of COVID-19-deaths. Finally, mortality is hierarchically the most important outcome, see GRADEpro (2013).

18 Scopus returns 947 hits on mortality, etc. between January 1, 2020, and June 30, 2021. Searching for hospitalization etc. yields another 35 hits corresponding to 3.7% more studies.

19 E.g. Madsen et al. (2014) find that “high bed occupancy rates were associated with a significant 9 percent increase in rates of in-hospital mortality and thirty-day mortality, compared to low bed occupancy rates. Being admitted to a hospital outside of normal working hours or on a weekend or holiday was also significantly associated with increased mortality.”

20 These simulations are often made in variants of the susceptible-infected-recovered (SIR) model, which can simulate the progress of a pandemic in a population consisting of people in different states (Susceptible, Infectious, or Recovered) with equations describing the process of moving between these states.

21 See Mailman School of Public Health, Columbia (2007) which has also inspired Figure 5 and other parts of the description of the difference-in-difference method.

22 For a more in-depth discussion of the parallel assumption, we refer to McKenzie (2020a), McKenzie (2020b), and McKenzie (2021). Also see McKenzie (2022a) and McKenzie (2022b).

23 As we describe on page 81, Arnarson (2021) and Björk et al. (2021) show that areas in Europe where the winter holiday was relatively late (in week 9 or 10 rather than week 6, 7 or 8) were hit especially hard by COVID-19 during the first wave because the virus outbreak in the Alps could spread to those areas with ski tourists.

24 The effective reproductive number, denoted R_t_, is the expected number of new infections caused by an infectious individual.

25 For example, Dr. Seth Flaxman said “Lockdown averted millions of deaths”, see e.g. https://www.bbc.com/news/health-52968523, and Samir Bhatt said “Our estimates show that lockdowns had a really dramatic effect in reducing transmission,” and ““Without them [lockdowns] we believe the toll would have been huge,”, see e.g. https://news.wgcu.org/2020-06-09/modelers-suggest-pandemic-lockdowns-saved-millions-from-dying-of-covid-19.

26 For example, see https://www.reuters.com/article/us-health-coronavirus-lockdowns-idUSKBN23F1G3, https://www.bbc.com/news/health-52968523, https://www.imperial.ac.uk/news/198074/lockdown-school-closures-europe-have-prevented/, https://www.france24.com/en/20200609-covid-19-lockdowns-saved-millions-of-lives-and-easing-curbs-risky-studies-find, and https://www.washingtonpost.com/health/2020/06/08/shutdowns-prevented-60-million-coronavirus-infections-us-study-finds/.

27 Brauner et al. (2021) states that “if increasing availability of information reduces infection growth rates, it would cause us to overstate the effectiveness of anti-contagion policies” and Hsiang et al. (2020) write that “our approach cannot distinguish direct effects on transmission in schools and universities from indirect effects, such as the general population behaving more cautiously after school closures signaled the gravity of the pandemic”.

28 Several scholars have criticized Flaxman et al. (2020), e.g., see Homburg and Kuhbandner (2020), N. Lewis (2020), and Lemoine (2020).

29 A jurisdictional area can be countries, U.S. states, or counties. With “jurisdictional variance” we refer to variation in mandated lockdowns across jurisdictional areas.

30 One can discuss whether our criteria should in fact exclude Wu and Wu (2020), when their data include all U.S. states. However, by pooling the data, they also lose much of the variation in data which was our primary reason for requiring a minimum of jurisdictional variance. Excluding Wu and Wu (2020) is unlikely to change our results as they conclude that “there were no significant differences in total positive cases, average daily new cases and average daily fatality among the 3 groups [SIPO and mask mandates, SIPO but no mask mandate, and neither SIPO nor mask mandate] during the pandemic period. This study suggested that SIPO and mask mandate orders in the general public alone, probably have limited effects in lowering transmission and fatality.”

31 Also see Abadie (2021).

32 For more on this thesis see e.g. Abernethy and Glass (2022) and Oraby et al. (2021).

33 This exclusion criteria was mistakenly not made public in our protocol Herby et al. (2021), but “only looks at timing” was decided upon as an exclusion criteria mid-September 2021 (documentation can be provided on request). Also see Herby (2021d).

34 They estimate that 10-point higher stringency will reduce excess mortality by 20 “per week and million” in the 10 weeks from week 14 to week 23.

35 Wilder-Smith and Osman (2020) state that “six events were declared PHEIC between 2007 and 2020: the 2009 H1N1 influenza pandemic, Ebola (West African outbreak 2013-2015, outbreak in Democratic Republic of Congo 2018-2020), poliomyelitis (2014 to present), Zika (2016) and COVID-19 (2020 to present).”

36 There’s approximately a two-to-four-week lag between infection and deaths. See footnote 29.

37 In his book, “The Premonition: A Pandemic Story”, M. Lewis (2021) describes how White House experts wanted to close schools in United States in the beginning of the 2009 swine flu, but did not have the necessary support to implement what the group thought was the right decision. It turned out that closing schools would in fact have been a mistake. Later, in early 2020, the same group again wanted to close schools, etc. Again, they did not have the necessary political support, but this time it remains unknown if it could potentially have saved a significant number of lives.

38 We describe how we arrive at the 2.4% in Section 4.

39 See Jonung and Hanke (2020).

40 The following numbers are based on data presented in Table 4.

41 Yue Li et al. (2021) use “data on the stringency of state social distancing measures from wallethub.com”.

42 One reason that we cannot calculate a counterfactual based on the estimated effect on growth rates is that we do not know the effect on the distribution and vertex of the death curve. If lower growth rates simply flatten the curve, the effect on total mortality can be limited even if the effect on growth rates is substantial.

43 The total is larger than 22 because the 12 SIPO studies include six studies which look at multiple measures.

44 See Banholzer et al. (2022).

45 Vetted papers from CEPR Covid Economics are considered as working papers in this regard.

46 Leffler et al. (2020) write “On average, the time from infection with the coronavirus to onset of symptoms is 5.1 days, and the time from symptom onset to death is on average 17.8 days. Therefore, the time from infection to death is expected to be 23 days.” Meanwhile, Stokes et al. (2020) state that “evidence suggests a mean lag between virus transmission and symptom onset of 6 days, and a further mean lag of 18 days between onset of symptoms and death.”

47 Some of the authors are aware of this problem. E.g. Bjørnskov (2021a) writes “when the lag length extends to three or fourth weeks, that is, the length that is reasonable from the perspective of the virology of Sars-CoV-2, the estimates become very small and insignificant” and “these results confirm the overall pattern by being negative and significant when lagged one or two weeks (the period when they cannot have worked) but turning positive and insignificant when lagged four weeks.”

48 See Chernozhukov et al. (2022) for a discussion of why this may affect estimates.

49 Eight studies are based on stringency data from OxCGRT, two are based on specific NPI data from OxCGRT, three are based on data from New York Times, one on data from COVID-19 Tracking Project, and one on data from Response2covid19. These 15 studies are considered being based on verified data. Three studies collect their own data, two studies rely on data from other studies, one study uses purchased data from Burbio.com, one study use American Red Cross reporting on emergency regulation. These six data sources are considered as being unverified (the latter two because they require login credentials, and thus are not easily verified by external researchers).

50 Research fields classified as social sciences are economics, public health, management, political science, government, international development, and public policy, while research fields not classified as social sciences were ophthalmology, environment, medicine, evolutionary biology and environment, human toxicology, epidemiology, and anesthesiology.

51 Standard errors are converted such that the t-value, calculated based on standardized estimates and standard errors, is unchanged. When confidence intervals are reported rather than standard errors, we calculate standard errors using t-distribution with ∞ degrees of freedom (i.e., 1.96 for 95% confidence interval).

52 An earlier version of our meta-study also included Stockenhuber (2020). However, Stockenhuber (2020) does not use a difference-in-difference approach and is excluded in this version.

53 The nine parameters are “C1 School closing,” “C2 Workplace closing,” “C3 Cancel public events,” “C4 Restrictions on gatherings,” “C5 Close public transport,” “C6 Stay at home requirements,” “C7 Restrictions on internal movement,” “C8 International travel controls” and “H1 Public information campaigns.” The latter, “H1 Public information campaigns,” is not an intervention following our lockdown definition, as it is not a mandatory requirement. However, of 97 European countries and U.S. States in the OxCGRT database, only Andorra, Belarus, Bosnia and Herzegovina, Faeroe Islands, and Moldova – less than 1.6% of the population – did not get the maximum score by March 20, 2020, so the parameter simply shifts the index parallelly upward and should not have notable impact on our conclusions.

54 Unless otherwise noted, we use these values in our calculations. The average stringency index is relatively stable during the first wave until end of June 2020. For instance, the average stringencies are 73 and 72, respectively, between March 16^th^ and June 30^th^, 2020.

55 In reality, the most lenient lockdown varies from study to study depending on the group of countries and/or states included in the study. However, for practical purposes our definition is sufficient to calculate standardized estimates.

56 The estimate from Fuller et al. (2021), the only study finding a substantial effect of lockdowns (−35%), corresponds to 103,000 avoided deaths in Europe and 70,000 avoided deaths in United States.

57 The average estimated flu deaths in United States the five years prior to COVID-19 was 38,400 according to the CDC (2022), and the WHO (2022) states that there are 72,000 flu deaths in Europe each year.

58 As noted earlier, Sweden limited public gatherings to 500 people on March 12, 2020, and – at the same time as Denmark and Norway – eliminated influenza, see Figure 8. Based on this experience, limiting gatherings to 500 people could be sufficient, if some NPI is required to spur voluntary behavioral changes.

59 Morgenstern (1963) describes in detail how such indices can be “fuzzy” metrics containing lots of errors in mismeasurement.

60 An earlier version of our meta-analysis also included Aparicio and Grossbard (2021) and Chaudhry et al. (2020). However, these studies do not use a difference-in-difference approach and have been excluded in this version.

61 The average estimated flu deaths in United States the five years prior to COVID-19 was 38,400 according to CDC (2022), and WHO (2022) writes that there are 72,000 flu deaths in Europe each year.

62 An et al. (2021) define early mandate adoption as being taken within 14 days (the median in their dataset) after the first reported infection in each country.

63 See Guallar et al. (2020), who conclude, “Our data support that a greater viral inoculum at the time of SARS-CoV-2 exposure might determine a higher risk of severe COVID-19.”

64 One could imagine that SIPOs also affected other types of deaths, including deaths by despair, impacts of deferred diagnoses, etc. However, all studies in Table 7 examine the effect of SIPOs on COVID-19 deaths – not overall mortality – so these are not likely explanations.

65 Both Nuzzo et al. (2019) and World Health Organization Writing Group (2006) focus on quarantining infected persons. However, if quarantining infected persons is not effective, it should be no surprise that quarantining uninfected persons could be ineffective too.

66 Note that we – according to our search strategy – did not search on specific measures such as “school closures” but on words describing the overall political approach to the COVID-19 pandemic such as “non-pharmaceutical,” “NPIs”, “lockdown” etc.

67 Say two studies, A and B, examine the effect of lockdowns. Study A examines school closure and business closure, whereas study B examines business closure and SIPO. Then, the estimates from study A could capture the effect of the omitted variable SIPO, and the estimates from study B could capture the effect of school closures. Based on study A and B, we would report precision-weighted averages on three estimates, but since they all potentially capture the effect of omitted variables, our precision-weighted average would be biased towards larger effects.

68 An earlier version of our meta-analysis included Bongaerts et al. (2021), which looked at business closures in Italy and thus did not meet our eligibility criteria of sufficient jurisdictional variance.

69 An earlier version of our meta-analysis included Auger et al. (2020), which looked at school closures in the United States. However, Auger et al. (2020) represents an interrupted time series analysis and, thus, did not meet our eligibility criteria.

70 According to Auger et al. (2020), all 50 states closed schools between March 13, 2020, and March 23, 2020, which means that all difference-in-difference is based on maximum seven school days (44 states closed schools in just four school days (March 15, 2020 (Sunday) to March 19, 2020 (Friday)), see Table 1 in Auger et al. (2020)).

71 Isphording et al. (2021) apply a quasi-experimental approach where they use the staggered timing of summer breaks across federal states in Germany to estimate the causal impact of school re-openings after the summer break in 2020. They find no evidence of a positive effect of school re-openings on case numbers. Rather, they find that the end of summer breaks had a negative but insignificant effect on the number of new confirmed cases.

72 See Hale, Angrist, Goldszmidt, et al. (2021a).

73 An earlier version of our meta-analysis also included Toya and Skidmore (2021). However, Toya and Skidmore (2021) does not use a difference-in-difference approach and is excluded in this version.

74 Lipp and Edwards (2014) also find no evidence of an effect and – looking at disposable surgical face masks for preventing surgical wound infection in clean surgery – conclude “Three trials were included, involving a total of 2113 participants. There was no statistically significant difference in infection rates between the masked and unmasked group in any of the trials.” Meanwhile, Yanni Li et al. (2021) – based on six case-control studies – conclude, “In general, wearing a mask was associated with a significantly reduced risk of COVID-19 infection (OR = 0.38, 95% CI: 0.21-0.69, I^2^ = 54.1%).

75 Note that the stringency index does not include face mask mandates.

76 The average estimated flu deaths in the United States the five years prior to COVID-19 was 38,400 according to the CDC (2022), and the WHO (2022) writes that there are 72,000 flu deaths in Europe each year.

77 The average estimated flu deaths in the United States the five years prior to COVID-19 was 38,400 according to the CDC (2022), and the WHO (2022) writes that there are 72,000 flu deaths in Europe each year.

78 The five years prior to COVID-19, the flu caused 38,400 deaths annually on average according to CDC (2022).

79 For example, using survey data immediately (same day) before and after Boris Johnson announced the UK lockdown, Eggers and Harding (2021) find that “the lockdown announcement made people more supportive of the government’s response to the crisis but also (perhaps surprisingly) more concerned about the pandemic”. E.g., people responding after the announcement were more likely to respond “I fear for my future” and “I have started not going out at all”.

80 Some have argued that mask mandates may be efficient regulation during future influenza seasons. However, an effect of 18.7% will only reduce the number of deaths by approximately 25.000 in Europe and the United States combined during an average flu season. Although this *is* a large benefit, it should be compared to the economic cost of mandating more than 1 billion people to wear facemasks.

81 We are uncertain if these numbers are correct, as Poeschl and Larsen (2021) list only one study examining bar closures in their overview table, although they review two studies. Also, Poeschl and Larsen (2021) do not look at business closures, and at least one of their studies examine this. Weber (2020) writes that “In an estimation without the non-positivity constraints, the sum of all sector closure effects is insignificant at the one percent level”.

82 If the true estimate was far from zero, we would expect to see relatively few estimates which are zero or positive (more deaths). If, on the other hand, the true value is around 0, we would expect to see that approximately half of the “guesses” are greater than zero, while half are lower than zero. Such an outcome is illustrated in the figure below.

83 Talic et al. (2021) is another systematic review and meta-analysis which looks at the effectiveness of public health measures in reducing the incidence of COVID-19. However, their focus is on voluntary measures. They state that “the findings of this review suggest that personal and social measures, including handwashing, mask wearing, and physical distancing are effective at reducing the incidence of covid-19. More stringent measures, such as lockdowns and closures of borders, schools, and workplaces need to be carefully assessed by weighing the potential negative effects of these measures on general populations Further research is needed to assess the effectiveness of public health measures after adequate vaccination coverage”.

84 E.g., Bjørnskov (2021a).

85 E.g., Goldstein et al. (2021).

86 E.g., Dave et al. (2021) states that “estimated case reductions accelerate over time, becoming largest after 20 days following enactment of a SIPO. These findings are consistent with a causal interpretation.”

87 Sebhatu et al. (2020) estimate that the impact of “death rate (/100,000)” on stringency is 11.706 (Table S3) and highly significant. On average, the death rate (/100,000) was 0.18 (Table S4) meaning that the average explanatory power was 2.1 stringency points. According to OxCGRT, the average death rate (/100,000) in Europe was approximately 0.08 and 0.19 when reaching stringency 60 and 70, respectively. Only San Marino had a substantial death rate and – given the estimate from Sebhatu et al. (2020) – a substantial impact on stringency, when reaching stringency 60 (and Netherlands and Spain when reaching stringency 70). In San Marino, Netherlands, and Spain the total impact measured as death rate estimate was 34, 5, and 2 respectively when reaching stringency index 60 and 172, 15, and 13 respectively when reaching stringency index 60. No other country had an impact above 5 points.

88 Note that the “RCT like” studies, we have identified, support our conclusions based on observational studies. See Table 17.

89 See, for example, https://www.spectator.co.uk/article/how-did-sage-scenarios-compare-to-reality-an-update and https://www.svd.se/hundratusen-skulle-do--modellen-slog-fel.

90 In a randomized control trial, Helsingen, Løberg, et al. (2020) find that “provided good hygiene and social distancing measures, there was no increased COVID-19 spread at training facilities.” Their study shows that many activities can be safe with focus on hygiene and social distancing.

91 Andersen et al. (2020) analyze transaction data for Denmark and Sweden from a large bank in Scandinavia to reach this conclusion.

92 In fact, Ferguson et al. (2020) describe their results as “unlikely”, as they are based on the assumption of the “absence of any […] spontaneous changes in individual behavior”.

93 Munch et al. (2022) write “contact (OR 4.9, 95% CI 2.4–10) and close contact (OR 13, 95% CI 6.7–25) with a person with a known SARS-CoV-2 infection were main determinants. Contact most often took place in the household or workplace”.

94 23% were infected in the household, 27% were infected at work, and 19% were infected by close acquaintances.

95 This kind of behavior response may also explain why Subramanian and Kumar (2021) find that increases in COVID-19 cases are unrelated to levels of vaccination across 68 countries and 2947 counties in the United States. When people are vaccinated and protected against severe disease, they have less reason to be careful.

96 In economic terms, lockdowns are substitutes for – not complements to – voluntary behavioral changes.

97 For cities with a response time <4 days, the average excess morality (unweighted) is 458 compared to 500 (4-9 days) and 486 (>9 days) if we only include cities with a “mortality acceleration date” after September 24, 1918. Choosing later cut-off dates does not change the picture. For Hatchett et al. (2007), this comparison is more difficult, because there is little overlap between early and late intervention cities.

98 As an example of how difficult timing is, Herby (2021b) and Herby (2021c) show how the Danish authorities – responding to local outbreaks in the fall of 2021 – in several occasions closed schools *after* case numbers had regressed to the level before the outbreak. The figures below illustrate this fact. The black line is test-corrected case numbers, the red vertical line is the day the schools closed, and the pink vertical line is the earliest day the effect of the school closure can be seen in cases (in Denmark in the fall of 2021, people were advised to wait four days between close contact to an infected person and being tested). 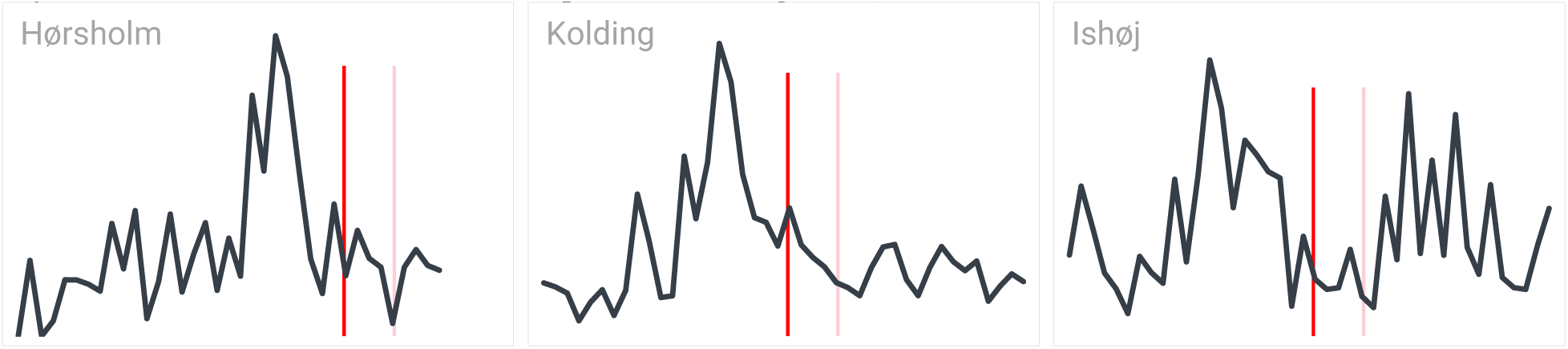

99 This is partly due to policy lags, i.e. the lag between the time an epidemic problem arises, and the effect of a policy intended to counteract it. In monetary and fiscal policies, the four lags are the *recognition lag*, the *decision lag*, the *implementation lag* and the *effectiveness lag*. These lags are likely to be relevant for pandemic policies too. As described on p. 19, the first time WHO characterized COVID-19 as a pandemic was by March 11, 2020, 2½ month after the first case in China.

100 In general, 3-4 clinical trials are required before a drug will be approved. Usually, a Phase 1 for safety, a Phase 2 to find the best dose (most effective while being safe), and 1-2 randomized Phase 3 trials to confirm the benefit seen in the Phase 2 trial.

101 The search was based on the same methodology as in our search strategy described in section 2.1, but we replace the methodology search string with “natural experiment” and “regression discontinuity”.

102 In their abstract they write 7.6%, but that result is from their difference-in-differences model. The 5.4% is from their regression discontinuity design (see Supporting Information Table A14).

103 By December 19, 2020, 321,035 had died with COVID-19 in the United States. The results from Hansen and Mano (2021) imply that 263,035 would have died with a nationwide mandate and 408,035 without any mandates. Hence, the effect of masks is 36%.

104 Hansen and Mano (2021) writes: “Specifically, mask mandates reduce COVID-19 cases and deaths by −78.03 and −1.45, respectively, in the median county more positively inclined to wearing masks in our sample. While the same numbers for the median county more negatively inclined towards to wearing masks are −44.54 and −0.37, also respectively”.

105 Source: https://crisis24.garda.com/alerts/2020/10/slovakia-authorities-to-introduce-partial-nationwide-lockdown-from-october-24-update-13.

106 See https://www.reuters.com/world/asia-pacific/melbourne-readies-exit-worlds-longest-covid-19-lockdowns-2021-10-20/.

107 Source: Hale, Angrist, Goldszmidt, et al. (2021a). Note that Hale, Angrist, Goldszmidt, et al. (2021a) only report one the extent of lockdowns in countries with several jurisdictional regions may be over.

108 The NUTS classification (Nomenclature of territorial units for statistics) is a hierarchical system for dividing up the economic territory of the EU and the UK. There are 1215 regions at the NUTS 3-level.

109 It is remarkable that the five countries above the confidence interval in Figure 18 – Belgium, Italy, the United Kingdom, Spain, and Sweden – all can be found among the countries hit early by the pandemic, see Figure 7, which may indicate that the mortality rates in these countries could have been much lower, if they had not been among the first countries to be hit by the COVID-19 pandemic. (There are no country labels in Figure 7, but the five countries all experienced more than 500 deaths per million during the first wave. The mortality rates by June 30, 2020, were 838 per million in Belgium, 576 per million in Italy, 593 per million in United Kingdom, 615 per million in Spain, and 525 per million in Sweden).

110 Another example is the Danish Prime Minister, Mette Frederiksen, who on several occasions said that every single death is a tragedy. A word choice that was very distinct from that of the Swedish state epidemiologist Anders Tegnell’s.

111 Another case of coincidence is illustrated by Shenoy et al. (2022), who find that areas that experienced rainfall early in the pandemic realized fewer deaths because the rainfall induced social distancing.

112 As noted by Thaler and Sunstein (2009), it might be fruitful to consider how nudging can influence citizens’ behavior without coercion.

113 In a review, Onyeaka et al. (2021) conclude that “the impact of the lockdown has had far-reaching effects in different strata of life, including; changes in the accessibility and structure of education delivery to students, food insecurity as a result of unavailability and fluctuation in prices, the depression of the global economy, increase in mental health challenges, wellbeing and quality of life amongst others.”

114 Engzell et al. (2021) find that children, on average, learn almost nothing in the weeks they received virtual education. The effect was particularly pronounced in less educated homes, where the negative effects for children with low-educated parents were 60 percent greater than those from well-educated homes. Rose et al. (2021) find that school closures in the UK in the spring of 2020 had put six- and seven-year-old students about two months behind in reading and seven months in math. Lindberg (2021) find that half of all students between 5th and 8th grade experienced got significantly less out of online teaching. Lindberg (2021) notes that school closures especially have caused academic and well-being problems among students from socially disadvantaged homes. Rajmil et al. (2021) indicate that children’s health has suffered in most of the world. Of particular concern is new research showing that even very young children’s development has suffered tremendously during the lockdowns. Some of the clearest evidence for the problems comes from Rhode Island, where Brown University’s Resonance project, Deoni et al. (2021), has followed children’s developments for more than ten years. Children’s development is measured with so-called Mullen scales for early learning, which measure their fine motor skills, language and visual reception, and which are considered a counterpart to intelligence goals for young children. Deoni et al. (2021) find that children’s scores on these goals have dropped markedly between 27 to 37 points compared to the normal of about 100 points before the shutdowns. The study also finds, like several other studies, that the losses in early development are significantly greater among poor and poorly educated families. Halloran et al. (2021) find that passing rates declined 10.1% in districts *without* in-person teaching relative to districts *with* in-person teaching and that the effect was larger in districts with larger populations of students who are Black, Hispanic or eligible for free and reduced price lunch. Goldhaber et al. (2022) conclude that “remote instruction was a primary driver of widening achievement gaps. Math gaps did not widen in areas that remained in-person (although there was some widening in reading gaps in those areas).”

115 See Wood et al. (2021).

116 Patrick et al. (2022) find that deviations from expected historical trends in alcohol use that occurred during the pandemic among U.S. young and middle adults included increases in alcohol use frequency and increases in the use of alcohol to relax/relieve tension and because of boredom. These shifts were likely due, in part, to drinking while alone and at home.

117 A crucial research object here is to determine not only how many deaths lockdowns avoid gross (e.g., avoided COVID-19 deaths), but also the net effect on excess mortality over a longer period. Also crucially, it matters which lives are saved and what lives are lost due to lockdowns. Lockdowns have an impact on the Health-Related Quality-of-Life (HRQoL) of the population, a key concept in health economics. Deaths results in a health loss. Loss of health can also be a consequence of a loss in the quality of life. Quality-adjusted life years (QALY) is a generally world-wide accepted measure of health loss which include both loss of health due to loss of life expectancy and loss of quality of life. In a recent study by Persson et al. (2021), the HRQoL was measured by a web based survey sent to randomized samples of the adult Swedish population before the outbreak of the pandemic in Sweden in February 2020 (n=1,016) and during the outbreak of the pandemic. The first wave pandemic data was collected in April 2020 (n=1,003), one-month after the outbreak and, the second wave data was collected in January 2021 (n=1,013), after 10-months living under the pandemic. The number of quality-adjusted life-years (QALYs) lost per month was calculated for both pandemic surveys. Loss of health for the Swedish population for one month in April 2020 was 29 000 QALYs and for the other month January 2021 was estimated to 44 000 QALYs. These monthly losses of health are in the same magnitude as the total loss of health due to excess mortality for the entire year 2020 which is 42 802 QALYs. Similar estimates of the NPIs and the related loss of quality of life are also available from the US, see Hay et al. (2021). It is fair to conclude that any analysis of the costs to society of NPIs should include measures of the HRQoL. However, such measures are so far only available for a few countries like Sweden and the United States.

118 For example, Dr. Seth Flaxman wrote that “smoking causes cancer, the earth is round, and ordering people to stay at home (the correct definition of lockdown) decreases disease transmission”. Needless to say, this is in no way a scientific comment. The effect of lockdowns is – unlike smoking and the shape of the Earth – still an open scientific question. And despite Flaxman’s attempt to monopolize the term “lockdown,” our definition is widely used (see footnote 1 on p. **Error! Bookmark not defined.** for an example).

119 For example, Dr. Seth Flaxman wrote that “[the only studies looked at] do not include key facts about disease transmission such as… lockdowns do not immediately save lives, because there’s a lag from infection to death, so to see the effect of lockdowns on Covid deaths we need to wait about two or three weeks.” We were very aware of this point and mentioned this lag several times in our meta-analysis. For example, we wrote “we distinguish between studies which find an effect on mortality sooner than 14 days after lockdown and those that do not.”

120 Prof. Samir Bhatt wrote that “two years in, it seems still to focus on the first wave of SARS-COV2 and in a very limited number of countries.” Dr. Bhatt made this statement even though we clearly described how we had gathered all available studies which covered Europe, the United States, and worldwide.

121 Referring to numbers in Figure 20: 1) The term “lockdown” has mainly been used to describe two different things. We define our use of lockdown on p. 2. 2) We examine the average lockdown – not the optimal lockdown. Hence, there are good reasons to exclude studies focusing on timing. We discuss timing on p. 16 and p. 71. 3) The quality of the included studies is handled using bias dimensions (see p. 33). 4) We clearly describe which studies are included and – to handle the “Other research shows lockdowns prevent deaths”-critique –we now explain in more detail why some of the prominent studies such as Flaxman et al. (2020) are both very problematic and ineligible, see section 2.2 on p. 11. We also relate our results to the conclusions in other reviews in section 5.2.1 on p. 61. 5) We use more bias-dimensions to handle the “Used incorrect statistical methods”-critique (see p. 33). 6) The “authors are economists” is an unscientific and irrelevant comment which we do not handle, but simply note that economists are skilled in handling meta-analysis in a wide range of contexts. 7) The “not endorsed by Johns Hopkins” critique is an unscientific and irrelevant comment. As a matter of fact, the Johns Hopkins University does not endorse specific research projects published by its faculty and staff. That said, our study was published by the Johns Hopkins Institute for Applied Economics, Global Health, and the Study of Business Enterprise, a research institute located at the Johns Hopkins University. 8) The “authors are biased against lockdowns” critique is an unscientific comment which was handled in the working paper where our search strategy and eligibility criteria were clearly described (see section 2 on p. 8 in this updated version). 9) We disagree with the conclusion in Chisadza et al. (2021). We explain why in section 3.1, p. 21. 10) We handle this critique on p. 8, where we write: “We believe that one major mistake in our first version was our failure to explain that the overall conclusions do not depend on whether the impact of lockdowns on COVID-19 mortality was 0.2%, 3.0% or 15%.”

